# Genomic insights into the 2022–2023 *Vibrio cholerae* outbreak in Malawi

**DOI:** 10.1101/2023.06.08.23291055

**Authors:** Chrispin Chaguza, Innocent Chibwe, David Chaima, Patrick Musicha, Latif Ndeketa, Watipaso Kasambara, Chimwemwe Mhango, Upendo L. Mseka, Joseph Bitilinyu-Bangoh, Bernard Mvula, Wakisa Kipandula, Patrick Bonongwe, Richard J. Munthali, Selemani Ngwira, Chikondi A. Mwendera, Akuzike Kalizang’oma, Kondwani C. Jambo, Dzinkambani Kambalame, Arox W. Kamng’ona, A Duncan Steele, Annie Chauma-Mwale, Daniel Hungerford, Matthew Kagoli, Martin M. Nyaga, Queen Dube, Neil French, Chisomo L. Msefula, Nigel A. Cunliffe, Khuzwayo C. Jere

## Abstract

Malawi is experiencing its deadliest *Vibrio cholerae* (*Vc*) outbreak following devastating cyclones, with >58,000 cases and >1,700 deaths reported between March 2022 and May 2023. Here, we use population genomics to investigate the attributes and origin of the Malawi 2022– 2023 *Vc* outbreak isolates. Our results demonstrate the predominance of ST69 seventh cholera pandemic El Tor (7PET) strains expressing O1 Ogawa (∼80%) serotype followed by Inaba (∼16%) and typical non-outbreak-associated non-O1/non-ST69 serotypes (∼4%). Phylogenetic reconstruction of the current and historical *Vc* isolates from Malawi, together with global *Vc* isolates, suggested the Malawi outbreak strains originated from Asia. The unique antimicrobial resistance and virulence profiles of the 2022–2023 isolates, notably the acquisition of ICE^GEN^/ICEVchHai1/ICEVchind5 SXT/R391-like integrative conjugative elements and a CTXφ prophage, which caused *ctxB3* to *ctxB7* genotype shift, support the importation hypothesis. These data suggest that the recent importation of *ctxB7* O1 strains, coupled with climatic changes, may explain the magnitude of the cholera outbreak in Malawi.

## Introduction

*Vibrio cholerae* (*Vc*) is a Gram-negative curved-rod-shaped bacterium that causes outbreaks and epidemics of life-threatening severe acute watery diarrhoeal illness, cholera, which is associated with high morbidity and mortality if untreated^1^. Cholera causes ∼3 million cases globally per year, leading to nearly 100,000 deaths, with a two-fold higher case fatality rate in Africa^2, 3^. The World Health Organisation (WHO) recommends the use of highly effective oral rehydration therapy using polymer-based or glucose-based rehydration solutions for mild or moderate infections and intravenous rehydration therapy complemented with antibiotics to treat severe cholera infection^4, 5^. *V. cholerae* is primarily transmitted from person to person through faecal contamination routes or poor food hygiene, and from the environment to person via *Vc-* contaminated water reservoirs^6^. Introductions of *Vc* strains from other countries^7, 8^, during humanitarian crises (war)^6, 9^ and natural disasters (earthquakes and cyclones)^10, 11^, which disrupt water and sanitation systems or displace populations towards inadequate and overcrowded living conditions, increase the risk of cholera transmission^1^.

More than 200 *Vc* serogroups have been characterised and are differentiated serologically based on the O-antigen of its cell surface lipopolysaccharide (LPS)^1^. However, only the O1 and O139 serogroups are typically associated with cholera outbreaks and epidemics, particularly in endemic settings^12^. The O1 serogroup is further divided into phenotypically distinct biotypes, namely, classical and El Tor, that evolved from independent lineages and the former is associated with earlier pandemics^12^. The O1 serogroup biotypes are further subdivided into the Ogawa and Inaba serotypes.

Although the modern history of cholera dates to 1817, several accounts of cholera-like illnesses were reported in the ancient times of Hippocrates circa, 300–500 BC^13^. Serogroup O1 was responsible for the early cholera outbreaks associated with the seven cholera pandemics starting from 1817^12^. However, O139 *Vc* strains resembling serogroup O1 strains emerged in the early 1990s, first reported in India^14^ and caused outbreaks in Bangladesh in the early 1990s^15, 16^. The current seventh cholera pandemic El Tor (7PET) lineage backbone, principally associated with the O1 serogroup and rarely with O139, dates back to 1961 in Sulawesi, Indonesia. E1 Tor subsequently spread globally^8^, including the first introductions into Africa in 1970, and it has persisted ever since^7^. Detailed phylogeographic analysis revealed twelve independent introductions of *Vc* into Africa from other continents (designated T1-T12) due to a single expanded multidrug-resistant (MDR) *Vc* lineage^7^. In addition to the serogroup and biotypes, the presence and absence of critical virulence factors are widely used to distinguish *Vc* strains. These *Vc* virulence factors include the cholera toxin (CT) carried on the filamentous lysogenic CTXφ prophage^17, 18^, encoded by *ctxA* and *ctxB* genes, which is responsible for the manifestation of severe watery “rice-water” diarrhoea with ongoing purging^6, 19^. The toxin-coregulated pilus (TCP), a receptor for the CTXφ phage^20, 21^, is the second most important *Vc* virulence factor encoded by the TCP operon in the Vibrio pathogenicity island (VPI-1)^21^ required for *Vc* colonisation of the small intestinal epithelium^22^. Both virulence factors are differentially expressed between classical and El Tor biotypes and other genes encoded on prophages and pathogenicity islands, including the Vibrio seventh pandemic island II (VSP-II) element commonly associated with 7PET *Vc* isolates^23^. In addition, the acquisition of mobile genetic elements (MGE), including plasmids, transposons, integrons and integrative conjugative elements (ICE)^24^, provides further context for characterising *Vc* isolates globally. These ICEs include the SXT/R391 family, which was first identified in an O139 isolate in 1993 in India and carried genes conferring antimicrobial resistance (AMR) to sulfamethoxazole/trimethoprim (SXT)^25^, and other variants of this ICE have been reported in recent years^26, 27^.

Cholera has been endemic in Malawi since 1998 and remains a major public health concern, with frequent seasonal outbreaks reported annually during the rainy season (November to May)^28^. The number of reported cholera-attributable deaths during seasonal outbreaks in Malawi rarely exceeds 100, with notable exceptions during the 1998–1999, 2001–2002 and 2008–2009 outbreaks, which resulted in 25,000, 33,546, and 5,751 cases with 968, 860 and 125 deaths, respectively^28^. However, since March 2022, Malawi has experienced its largest cholera outbreak, with >58,000 cases and >1,700 deaths reported countrywide across all 29 districts^11, 29^. The reasons for the high morbidity and mortality and persistence of the ongoing 2022–2023 cholera outbreak, beyond the devastation caused in Malawi by tropical cyclones Ana and Gombe in early 2022 and Freddy in 2023, remain unknown.

Here, we describe the epidemiology of cases and deaths attributed to cholera during the 2022– 2023 outbreak in Malawi and examine the origin and genomic attributes of *Vc* isolates collected as part of the national public health response. Unlike the traditional typing methods widely used to characterise *Vc*, such as serotyping, phage typing, and determination of antibiograms, whole-genome sequencing provides greater resolution, allowing for adequate characterisation of clones and the genetic repertoire of the isolates^12^. We specifically performed whole-genome sequencing of *Vc* isolates collected nationwide and compared them to the contextual historical *Vc* isolates from Malawi and globally. We determined the serogroups, serotypes and biotypes, and genetic similarities, together with the distribution of virulence, and AMR gene profiles of the *Vc* isolates, to understand potential pathogenic traits that may have contributed to the magnitude of the ongoing outbreak in Malawi. Our findings provide the first insights into the evolution and genomic diversity of the 2022–2023 cholera outbreak-associated *Vc* isolates in Malawi to inform public health strategies to prevent and control current and future cholera epidemics.

## Results

### The 2022–2023 *V. cholerae* outbreak is the deadliest recorded in Malawi

The WHO defines Malawi as a cholera-endemic country with annual outbreaks occurring during the rainy season from November to May. Based on data from the Malawi Ministry of Health (MoH) data (Accessed on May 20, 2023; https://cholera.health.gov.mw/surveillance), the 2022– 2023 cholera outbreak has to date resulted in 58,730 cases and 1,759 deaths in Malawi **(Fig. 1a,b)**. These case and death counts make the 2022–2023 cholera outbreak the largest and deadliest cholera outbreak recorded in Malawi^30^.

**Figure 1:**
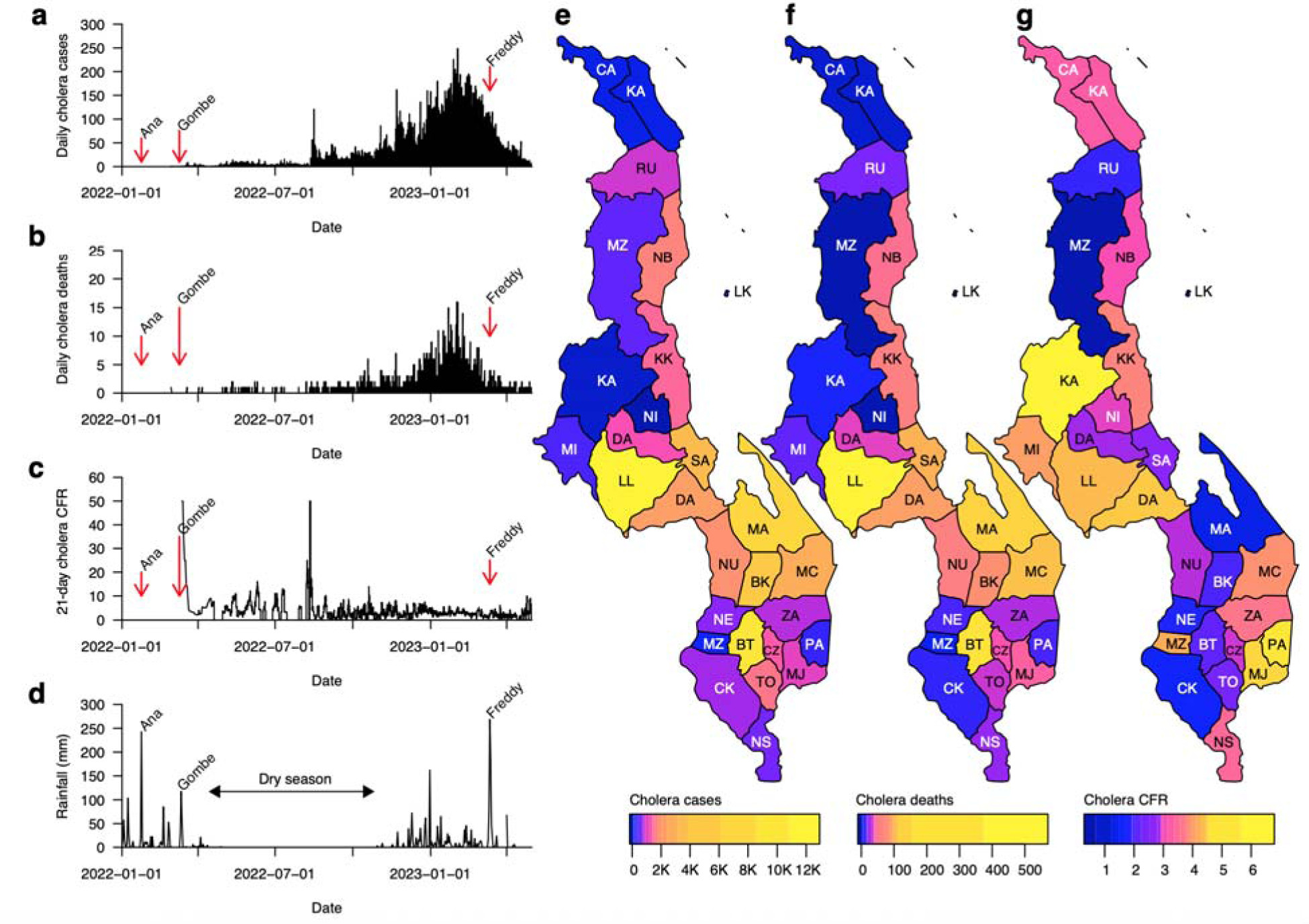
Cases, deaths, and case fatality ratio during the 2022–2023 cholera outbreak in Malawi (data from January, 2022 to May, 2023). **(a)** Total daily cholera cases. **(b)** Total daily cholera deaths. **(c)** Overall cholera case fatality ratio (CFR) based on a 21-day sliding window. The 21-day sliding window was chosen to obtain stable estimates of the CFR, especially during weeks and months with few reported cholera cases. **(d)** Daily rainfall in millimeter (mm) units obtained from Chichiri weather station in Blantyre, Malawi. **(e)** Map of Malawi showing the number of cholera cases per district. **(f)** Map of Malawi showing the number of cholera deaths per district. **(g)** Map of Malawi showing the CFR of cholera cases per district. Data were obtained from the Public Health Institute of Malawi, Malawi Ministry of Health (MoH) data on May 20, 2023 (https://cholera.health.gov.mw/surveillance).

Based on these estimates, the 2022–2023 cholera outbreak in Malawi has a case fatality ratio (CFR) of 3.0% (1,759 of 58,730, cases; 95% confidence interval [CI]: 2.88 to 6%) **(Fig. 1c)**. This CFR is similar to that reported during the 1998–1999 outbreak in Malawi (3.4%; 860 of 25,000 cases) but slightly higher than those observed during 2001–2002 (2.3%;968 of 33,546) and 2008–2009 (2.17%; 125 of 5,751) outbreaks^28^. Similarly, the CFR for the current Malawi outbreak is higher than those reported from Malawi’s neighbouring countries; for instance, the CFR was 1.8% (98 of 5,414 cases) for the 2017–2018 outbreaks in Zambia and 0.7% (37, 5 of 237 cases) during the 2022–2023 outbreak in Mozambique^31^. On a global scale, the overall CFR for the 2022–2023 outbreak in Malawi is higher compared to the cholera outbreaks that occurred in 2016–2017 in Yemen (0.22%; 2,385 of 1,103,683 cases)^9^ and the 2010–2011 epidemic in Haiti (2.3%; 654 of 29,295 cases)^32^ cholera epidemics, but similar to that reported during the 2022–2023 outbreak in Port-au-Prince, Haiti (3.0%; 144 of >20,000 cases)^33^. The CFR estimates for the current outbreak in Malawi varied greatly by district, ranging from 0.62% (3 of 486 cases) to 6.67% (6 of 90 cases) in Mzimba and Kasungu districts, respectively. Similarly, the number of cases ranged from 68 in Ntchisi to 12,683 in Lilongwe **(Fig. 1e,f, Table 1)**. To account for the differences in the population size in each district, we calculated the overall cumulative incidence of cases per district per 100,000 people. The mean incidence was 352.91 (range: 10.68 to 1,404) cases per 100,000 people while the incidence of deaths was 8.66 (range: 0.32 to 21.24) deaths per 100,000 people **(Table 1 and Supplementary Fig. 1)**. These data highlight areas to be given the highest priority for public health outbreak prevention and control measures, including oral cholera vaccination^34, 35^.

**Table 1:** Summary of cholera cases, deaths, incidences, and case fatality ratio (CFR) per district across Malawi.

| District | Number of cases | Number of deaths | Population size | Overall incidence per 100,000 people |  | CFR* |
| --- | --- | --- | --- | --- | --- | --- |
|  |  |  |  | Cases | Deaths |  |
| Mzimba** | 486 | 3 | 940,184 | 51.69 | 0.3191 | 0.6173 |
| Ntchisi | 68 | 2 | 317,069 | 21.45 | 0.6308 | 2.941 |
| Kasungu | 90 | 6 | 842,953 | 10.68 | 0.7118 | 6.667 |
| Chitipa | 91 | 3 | 234,927 | 38.74 | 1.277 | 3.297 |
| Chikwawa | 748 | 11 | 564,684 | 132.5 | 1.948 | 1.471 |
| Mchinji | 394 | 16 | 602,305 | 65.42 | 2.656 | 4.061 |
| Phalombe | 302 | 14 | 429,450 | 70.32 | 3.26 | 4.636 |
| Zomba | 759 | 28 | 851,737 | 89.11 | 3.287 | 3.689 |
| Mwanza | 120 | 5 | 130,949 | 91.64 | 3.818 | 4.167 |
| Thyolo | 1,520 | 36 | 721,456 | 210.7 | 4.99 | 2.368 |
| Dowa | 1,349 | 39 | 772,569 | 174.6 | 5.048 | 2.891 |
| Mulanje | 936 | 42 | 684,107 | 136.8 | 6.139 | 4.487 |
| Karonga | 1,403 | 25 | 365,028 | 384.4 | 6.849 | 1.782 |
| Nsanje | 607 | 21 | 299,168 | 202.9 | 7.019 | 3.46 |
| Rumphi | 932 | 18 | 229,161 | 406.7 | 7.855 | 1.931 |
| Ntcheu | 1,926 | 56 | 659,608 | 292 | 8.49 | 2.908 |
| Neno | 733 | 14 | 138,291 | 530 | 10.12 | 1.91 |
| Mangochi | 8,182 | 120 | 1,148,611 | 712.3 | 10.45 | 1.467 |
| Dedza | 2,124 | 95 | 830,512 | 255.7 | 11.44 | 4.473 |
| Chiradzulu | 1,426 | 41 | 356,875 | 399.6 | 11.49 | 2.875 |
| Likoma | 204 | 2 | 14,527 | 1404 | 13.77 | 0.9804 |
| Machinga | 2,708 | 104 | 735,438 | 368.2 | 14.14 | 3.84 |
| Nkhotakota | 1,461 | 56 | 393,077 | 371.7 | 14.25 | 3.833 |
| Blantyre | 8,975 | 190 | 1,251,484 | 717.1 | 15.18 | 2.117 |
| Nkhata Bay | 1,628 | 49 | 284,681 | 571.9 | 17.21 | 3.01 |
| Balaka | 4,111 | 81 | 438,379 | 937.8 | 18.48 | 1.97 |
| Salima | 3,591 | 97 | 478,346 | 750.7 | 20.28 | 2.701 |
| Lilongwe | 12,683 | 558 | 2,626,901 | 482.8 | 21.24 | 4.4 |

\*CFR: Case fatality ratio
\*\*Mzimba shows combined cases from Mzimba South and Mzimba North districts
The population sizes per district is based on data reported by the National Statistics Office of Malawi for the 2018 population census in Malawi (http://www.nsomalawi.mw/)

**Table 2:** Summary of the major attributes of the 2022–2023 and historical *V. cholerae* isolates in Malawi.

| Attribute | 2022–2023 outbreak isolates | Historical isolates (80s and 90s) |
| --- | --- | --- |
| Species | *V. cholerae* | *V. cholerae* |
| Serogroup, serotype, and biotype | Mostly O1 Ogawa and a minority of O1 Inaba El Tor and sporadic non-O1/O139 | All O1 Ogawa El Tor |
| Pandemic strain | Pandemic | Pandemic |
| ST or clone | Mostly ST69 (outbreak); also, ST40 and ST635 (sporadic) | All ST69 |
| Genomic wave | Late wave 3 | Early wave 3 |
| Genetic markers | *ctxB7* | *ctxB3* |
| AMR profile | DOX<sup>S</sup>/TET<sup>S</sup>, CIP<sup>I</sup>, SXT<sup>R</sup>/SXZ<sup>R</sup>, STR<sup>R</sup>, CRO<sup>R</sup>, AMP<sup>R</sup>, CAZ<sup>R</sup>, CAR<sup>R</sup>, CEP<sup>R</sup>, CPM<sup>R</sup>, MER<sup>S</sup>, NA<sup>R</sup>, NIT<sup>R</sup>, STR<sup>R</sup>, TMP<sup>R</sup>, FZD<sup>R</sup>, and CHL<sup>R</sup> | DOX<sup>S</sup>/TET<sup>S</sup>, CIP<sup>S</sup>, SXT<sup>S</sup>/SXZ<sup>S</sup>, STR<sup>R</sup>, CRO<sup>R</sup>, AMP<sup>R</sup>, CAZ<sup>R</sup>, CAR<sup>R</sup>, CEP<sup>R</sup>, CPM<sup>R</sup>, MER<sup>S</sup>, NA<sup>R</sup>, NIT<sup>R</sup>, STR<sup>R</sup>, TMP<sup>S</sup>, FZD<sup>S</sup>, and CHL<sup>S</sup> |
| Horizontally acquired AMR-conferring ICEs | ICE<sup>GEN</sup>/ICEVchHai1/ICEVchind<br>5 SXT-like ICEs | None |
| Bacteriophages | CTXφ | CTXφ |
| Pathogenicity islands | VPI-1 and VSP-II<sup>**</sup> | VPI-1 and VSP-II<sup>***</sup> |
| <sup>**</sup> VSP-II pathogenicity island according to GenBank accession no. KM660639<br><sup>***</sup> VSP-II pathogenicity island according to GenBank accession no. KU601747<br>ST: Sequence type<br>AMR: Antimicrobial resistance<br>ICE: Integrative conjugative elements |  |  |

Uniquely, the 2022–2023 cholera outbreak in Malawi started towards the end of a typical seasonal cholera outbreak in Malawi, with the first *Vc* cases observed in March 2022, which persisted throughout the dry season until the 2022–2023 rainy season **(Fig. 1a)**. The number of cholera cases and deaths started to increase immediately following tropical cyclone Gombe, one of the largest cyclones recorded in Malawi, which occurred between March 8–14, 2022 **(Fig. 1b, Supplementary Fig. 2).** The number of cases increased steadily every month from March 2022 as new districts registered cases, leading to a substantial increase of cases along the lakeshore districts before a larger outbreak towards the start of the 2022–2023 rainy season. The larger outbreak reached a peak between in February 2023, before the occurrence of cyclone Freddy in March 2023 **(Fig. 1a,c)**. Whereas past seasonal cholera outbreaks in Malawi mostly occurred in a few districts, especially Machinga, Zomba, Thyolo, Nsanje, Chikwawa, and Phalombe, which are typically at the highest risk during the rainy season^36^, the 2022–2023 outbreak spread to all twenty-nine districts **(Fig. 1e,f)**. The cholera outbreaks that occurred in several districts did not entirely overlap with each other; instead, there was a sequential occurrence of outbreaks, starting in the Nsanje district, immediately after cyclone Gombe **(Supplementary Fig. 2)**. Following this initial rise of cases in this flood-prone district, an increase of cholera cases occurred a few weeks later in nearby districts in the lower Shire region in southern Malawi, including Chikwawa and Neno, which spilled over to Blantyre possibly igniting the spread of cases throughout the country. Cholera cases were reported in Nsanje, Chikwawa, Neno, and Blantyre throughout 2022, including the dry season, leading to the larger outbreaks observed in all districts after the start of the 2022–2023 rainy season **(Supplementary Fig. 2)**. Together, these findings demonstrate the countrywide spread of *Vc,* with variable incidence and temporal spread across districts in Malawi, which spanned the dry season during which when no cases are typically reported, possibly reflecting ongoing human-to-human transmission.

### Potential to incorrectly attribute cholera-like cases to *V. cholerae* in Malawi

Studies in Africa and elsewhere have suggested that non-*Vc* bacterial species may cause simultaneous cholera-like infections during cholera outbreaks^37–41^. However, it is currently unknown what proportion of cholera-like cases may be attributed to non-*Vc* bacterial pathogens. To begin to understand this in Malawi, we assessed the proportion of confirmed *Vc* and other bacteria based on whole-genome sequencing (WGS) data generated from presumptive *Vc* isolates from patients presenting with cholera or cholera-like symptoms. All the cultures that were subjected to WGS had yellow colonies on thiosulphate citrate bile salt sucrose (TCBS) agar, suggestive of *Vc* (see methods). We recovered sufficient genomic data from 68 out of 75 suspected *Vc* isolates, of which 49 of 68 cases (∼72%) of the sequenced genomes were confirmed to contain *Vc* based on a comparison of the genome assemblies to reference sequences of bacterial species in the National Center for Biotechnology Information (NCBI) RefSeq database^42^ and the presence of the *Vc*-specific *ompW* outer membrane protein-encoding gene^43^ **(Supplementary Data 2)**. Among the genomes containing the *ompW* and showing similarity to reference *Vc* sequences, 45 *Vc* genomes had no detectable contamination, i.e., genetic similarity to other pathogens beyond shared accessory genome content, including plasmid sequences, while the other four were mixed with other species. In a total of 19 of 68 cases (∼28%), the recovered genomes were associated with non-*Vc* species. In almost 6% (4 of 68 cases), the non-*Vc* isolates were associated with *Aeromonas caviae,* whereas the remainder of the genomes were associated with *Enterobacter cloacae* (∼4.4%, 3 of 68 cases)*, Providencia alcalifaciens* (∼1.5%, 1 of 68 cases), and *Escherichia coli* (∼1.5%, 1 of 68 cases), and a mixture of these and/or other bacterial pathogens (∼21%, 14 of 68 cases). These observations are consistent with reports elsewhere that other bacterial diarrhoea-associated gastrointestinal pathogens, especially those mimicking *Vc* enteropathy or those poorly investigated via routine laboratory diagnosis, may lead to misdiagnosis of *Vc* in patients with suspected cholera infection^37–41, 44–47^. These findings emphasise the need for further studies to assess the contribution of other non-*Vc* bacteria in cholera-like diarrhoeal diseases during seasonal cholera outbreaks.

### The 2022-2023 outbreak in Malawi was primarily driven by Ogawa and Inaba O1 serotypes from the ST69 clone although diverse lineages of *V.cholerae* circulated

concurrently We mapped genomic sequence *k-*mers of the 2022–2023 outbreak and historical *Vc* isolates from Malawi against all known reference LPS O-antigen biosynthesis gene cluster sequences to determine the specific serogroups, biotypes, and serotypes of the isolates^48^. We inferred the serotype as the reference LPS O-antigen biosynthesis gene cluster with the highest sequence coverage compared to the rest of the LPS O-antigen sequences. The percentage genome coverage for the highest matching LPS O-antigen biosynthesis gene cluster based on this *in silico* genomic-based serotyping approach ranged from ∼63% to 100% with a mean of ∼94% **(Supplementary Data 3)**. We found that ∼95% of 2022–2023 (42 of 44 clinical cases) and 100% of the historical (6 of 6 cases) *Vc* isolates from Malawi belonged to the O1 serogroup, El Tor biotype, confirming its primary role in the cholera outbreaks in Malawi, consistent with reports from other countries in Africa^7, 49–53^ and elsewhere^54–58^ **(Fig. 2a)**. We found the Ogawa serotype in ∼80% (35 of 44 cases) of the 2022–2023 clinical *Vc* O1 isolates, while ∼16% (7 of 44 cases) were of Inaba serotype. Although serotype Inaba has been associated with previous outbreaks in Malawi^59^, our current observations showed the predominance of the serotype Ogawa and the absence of serogroup O139 in the country^60^. The dominance of serotype Ogawa is consistent with recent studies conducted by others^61–64^. The presence of both Ogawa and Inaba O1 serotypes reflects the occurrence of capsule-switching events in the ST69 *Vc* isolates consistent with reports elsewhere, including in Haiti and Bangladesh^56, 65^. In contrast, the two non-O1 *Vc* isolates were assigned serogroups O7 and O19. The low detection rates of non-O1 *Vc* serogroups among the 2022–2023 isolates are consistent with reports from elsewhere that show that cholera outbreaks and epidemics are typically caused by serotypes O1 and O139^12, 66^. Therefore, we concluded that the *Vc* serogroup O1, serotype Ogawa strains are predominantly driving the ongoing 2022–2023 cholera outbreak in Malawi, and the non-O1 isolates were likely associated with sporadic seasonal cases.

**Figure 2:**
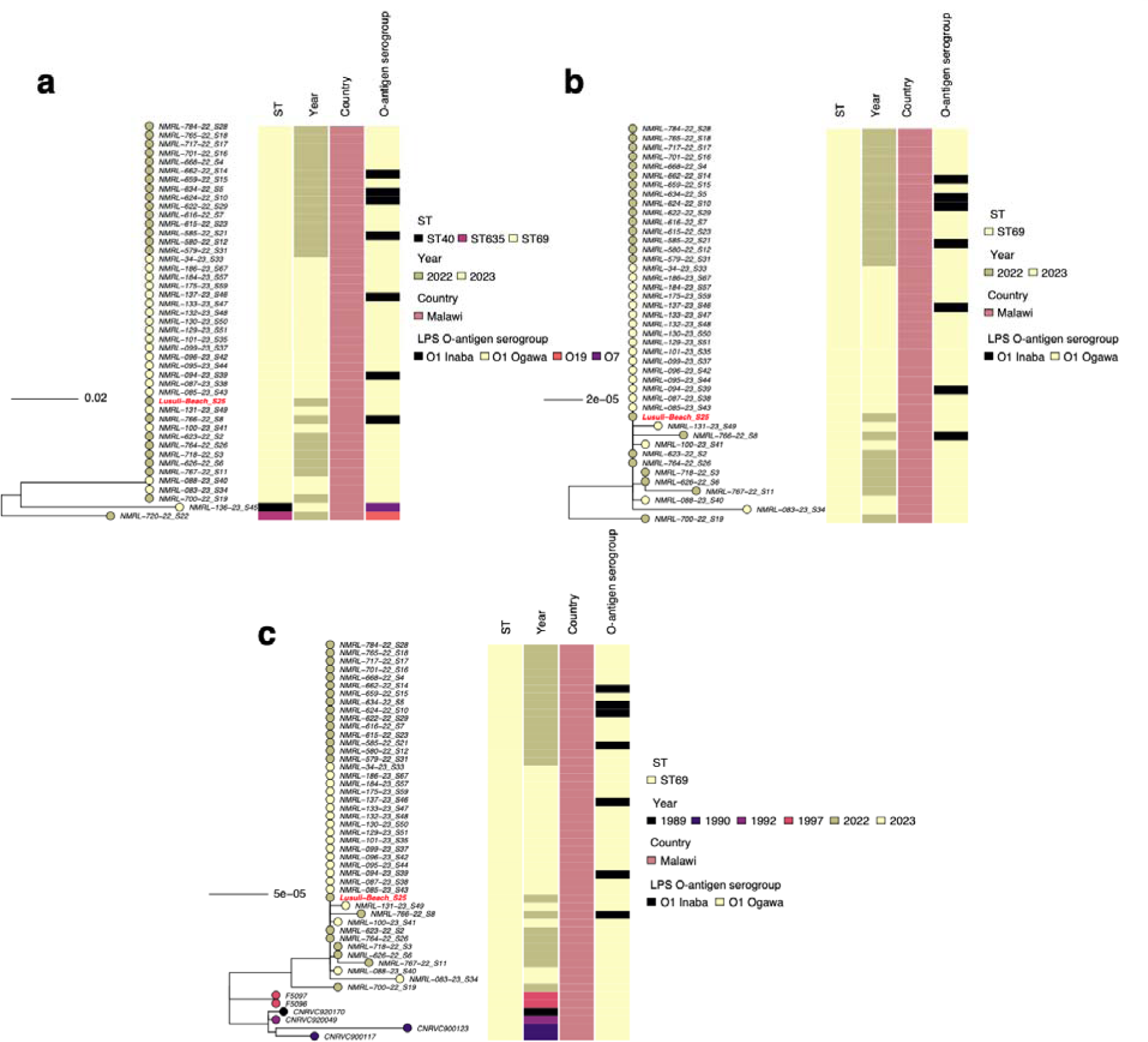
Genetic relatedness of the *V. cholerae* isolates from the 2022–2023 outbreak and the historical isolates from the late 1980s and 1990s in Malawi. **(a)** Maximum likelihood phylogenetic tree showing the genetic relatedness of all the *Vc* isolates collected between 2022– 2023. **(b)** Maximum likelihood phylogenetic tree showing the genetic relatedness of the ST69 *Vc* isolates collected between 2022–2023. **(c)** Maximum likelihood phylogenetic tree showing the genetic relatedness of the ST69 *Vc* isolates collected between 2022–2023 in the context of the historical ST69 *Vc* isolates from the late 1980s and 1990s in Malawi. The circles with different colours at the tip of the phylogeny represent the year of isolation. All the isolates were sampled from human clinical cases except one (coloured in red text), which was obtained from a water sample at a beach in the southern region of Malawi. The phylogeny is annotated by four column colour strips at the tips of each tree representing the sequence type (ST), year of isolation, country, and LPS O-antigen serogroup and/or serotype. The phylogeny was constructed based on the core-genome SNPs identified from the merged alignments of chromosomes 1 and 2, and rooted based on an outgroup *Vibrio mimicus* species, not shown in the tree (see methods).

As part of the present study, we sequenced a single environmental *Vc* isolate collected from a water sample at Lisulu Beach in the lower Shire region in Southern Malawi. Like most of the clinical *Vc* isolates, this isolate also belonged to ST69 lineage, and it was assigned serogroup O1, serotype Ogawa **(Fig. 1a)**. We then compared our 2022–2023 clinical and environmental *Vc* isolates to gain insights into their genetic diversity using phylogenetic analysis. The maximum likelihood phylogeny of these whole-genome sequenced isolates generated, based on the core genome multiple nucleotide sequence alignment of 93,704 single nucleotide polymorphisms (SNPs), revealed three genetically distinct clusters corresponding to the three ST clones. Ninety-five per cent of the isolates (43 of 45 cases), including the environmental isolate, belonged to the ST69 clone, a 7PET lineage widely responsible for seasonal cholera outbreaks and epidemics globally^49, 67–70^. All the ST69 isolates were assigned either serotype O1 Ogawa or Inaba, while the two non-O1 isolates, O7 and O19, belonged to ST40 and ST635 lineages, respectively **(Fig. 2a)**.

Our next step was to focus on the 2022–2023 ST69 isolates to gain further insights into these outbreak-associated *Vc* isolates. The maximum likelihood phylogeny placed some isolates, which differed from the rest of the isolates by up to 20 SNPs, at the base of the phylogeny **(Fig. 2b)**. The SNP differences amongst the outbreak-associated ST69 isolates were lower than the SNP thresholds previously used to distinguish outbreak-associated and sporadic isolates for several bacterial pathogens^71, 72^. These observations confirmed that the majority of the ST69 isolates were part of a single outbreak responsible for most of the reported cholera cases in Malawi. Considering the zero core-genome SNPs distinguishing the rest of the 2022–2023 ST69 isolates, we propose that a single strain, likely imported from a single geographic source, predominantly caused the 2022–2023 cholera outbreak in Malawi.

We then compared the 2022–2023 and historical *Vc* isolates from Malawi to understand whether the current outbreak strains evolved from previously locally circulating strains. Due to the lack of routine genomic surveillance of *Vc*, there is scant WGS data on isolates collected during the previous seasonal cholera outbreaks in Malawi. We searched publicly available WGS databases and found six whole genome sequences of *Vc* isolates from Malawi collected in 1989 (1 of 6 cases), 1990 (2 of 6 cases), 1992 (1 of 6 cases), and 1997 (2 of 6 cases), sequenced by the US Centers for Disease Control and Prevention (CDC) and Institut Pasteur in France^7^. A phylogenetic tree of the 2022–2023 *Vc* isolates in the context of these historical isolates from Malawi placed the historical isolates at the base of the tree and was distinguished by ∼19-45 SNPs from the current outbreak isolates **(Fig. 2c)**. This finding raised a possibility that the 2022– 2023 outbreak isolates were derived through stepwise evolution from the historical strains circulating in the 1980s and 1990s in Malawi. However, a comparison of the accessory genome and the phylogenetic context in the global context (described in the latter sections) ruled out this hypothesis, in favour of recent importation from outside the country. Together, these findings suggest that the *Vc* O1 Ogawa isolates belonging to the global 7PET, ST69 clone, are primarily driving the ongoing 2022–2023 cholera outbreak in Malawi.

### Genetic analysis suggests the 2022–2023 outbreak-associated *V. cholera* isolates were recently imported from Asia

To investigate the potential origin of the Malawi 2022–2023 outbreak-associated *Vc* isolates, we contextualised them in the global phylogeny. We obtained a large collection of 4,613 globally diverse *Vc* sequences from 110 countries/territories worldwide (see methods). Our collection included *Vc* sequences from notable recent and past cholera outbreaks and epidemics globally, including other African settings, which captured the global *Vc* genetic diversity **(Fig. 3)**. The non-ST69 Malawian isolates deemed to cause sporadic cases during the ongoing outbreak, belonging to ST40 and ST635 clones, clustered closest in the global phylogeny with a single isolate from India and three isolates from Austria of identical STs, respectively **(Fig. 3)**. The single ST40 isolate from India was collected in 1962 and differed from the 2023 Malawi isolate by 81 core-genome SNPs, which suggested a possible historical importation from Asia possibly in the 1960s or 1970s, consistent with early *Vc* introductions from Asia into Africa^7^. On the other hand, ∼627-689 core-genome SNPs separated the 2022 Malawi ST635 isolate from the three recent Austrian ST635 isolates sampled in 2012, implying a possible historical introduction from a shared source.

**Figure 3:**
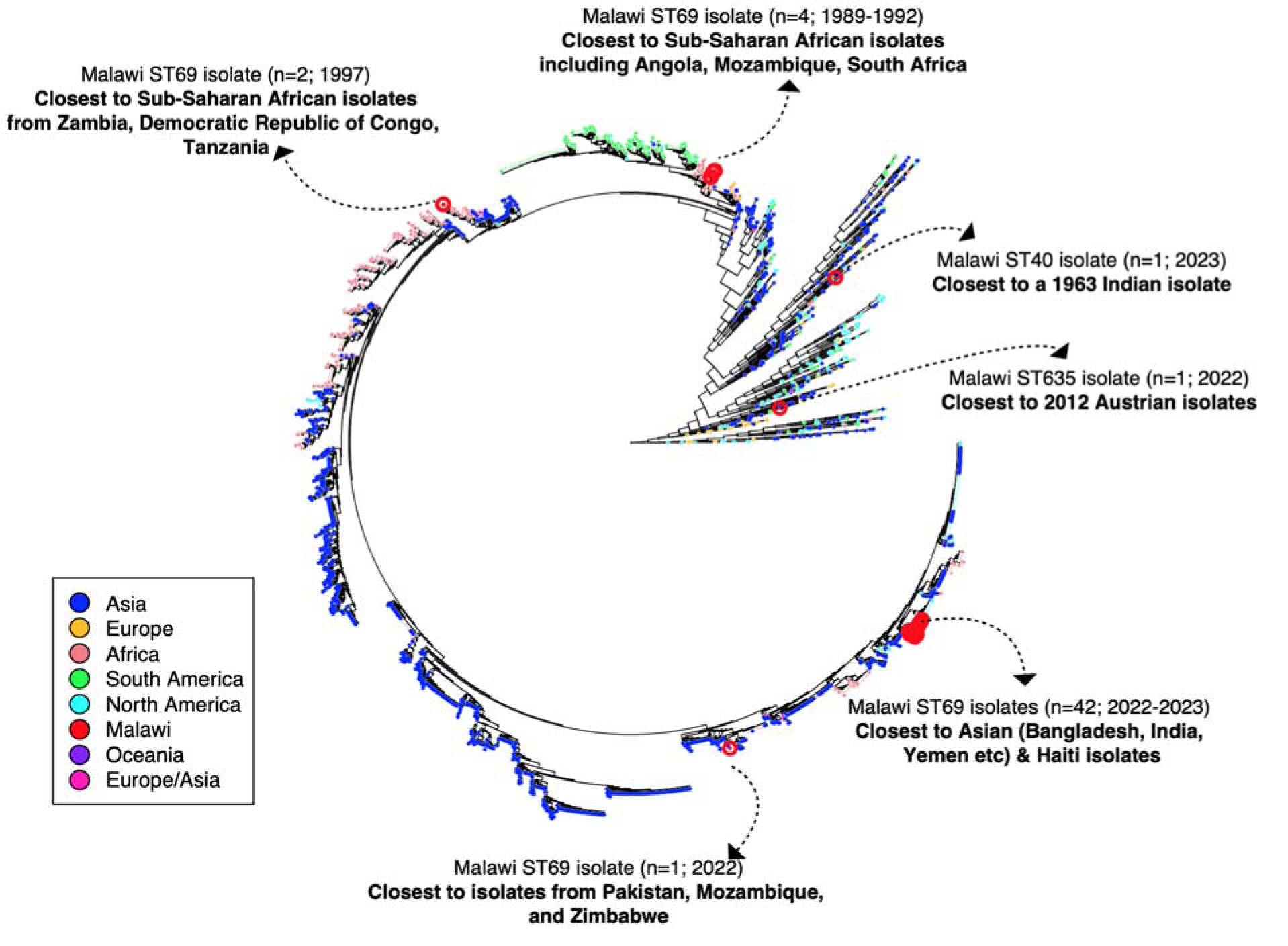
Maximum likelihood phylogenetic tree showing genetic relatedness of the 2022– 2023 and historical Malawi *Vc* isolates in the context of global *Vc* isolates. The branches of the phylogeny were transformed, to zoom in the shorter branches, especially at the tips of the tree, for clarity to broadly highlight the phylogenetic and global context of the *Vc* isolates from Malawi. The tips of the phylogeny are coloured by the geographical origin of the isolates with the isolates from Malawi highlighted by larger red circles. Examples of the *Vc* isolates from other geographical regions showing the closest genetic similarity to the 2022–2023 and historical isolates from Malawi are highlighted by arrows and text annotations. The phylogeny was constructed based on the core-genome SNPs identified from the merged alignments of chromosomes 1 and 2, and rooted based on an outgroup *Vibrio mimicus* species, not shown in the tree (see methods). An interactive phylogenetic tree of the current outbreak and historical *Vc* isolates from Malawi in the context of the global sequences is available on Microreact (https://microreact.org/project/malawi-vibrio-cholerae-2022-2023).

Among the ST69 isolates, one of the three slightly genetically divergent isolates was placed at the base of the phylogeny of Malawi isolates belonging to this clone and showed the closest genetic similarity with isolates sampled from Zimbabwe, Mozambique, and Pakistan **(Fig. 3)**. These isolates from Malawi and Zimbabwe appeared to be descendants of *Vc* strains collected from 2009 and 2010. In contrast, the predominant 2022–2023 outbreak-associated isolates from Malawi formed a single cluster in the global phylogeny **(Fig. 3)**. These 2022–2023 Malawi isolates showed the closest genetic similarity (∼3 core-genome SNPs) to isolates from multiple countries, including India (2011–2017), Nepal (2010), Mexico (2013), Russia (year unknown), Bangladesh (2018), Dominican Republic (2011), and Haiti (2010–2013) belonging to the T12 transmission lineage into Africa from Asia^7^. The most recent common ancestor of the current outbreak strain appears to be of Asian origin due to their near sequence identity to the majority of Asian and global *Vc* isolates previously reported to have descended from Asia^7, 8^, including those associated with the 2010 Haiti outbreak^54^. Such broader geographical dissemination poses a challenge in determining the precise country of origin for potential recent importations into Malawi, although the most recent ancestor of the introduced strains originated from Asia^7, 8^. Together, these findings suggest the predominant outbreak-associated strain in Malawi is a highly successful clone, which has been disseminated globally in the past two decades following its emergence in Asia^7, 8^.

We further investigated whether the Malawi 2022–2023 *Vc* outbreak strains were derived through stepwise evolution from the sequenced ancestral ST69 strains from the late 1980s and 1990s. A maximum-likelihood phylogenetic tree of the current outbreak and historical Malawi *Vc* isolates in the context of the global *Vc* sequences can be explored interactively at the Microreact webtool for open data visualisation and sharing for genomic epidemiology^73^ (https://microreact.org/project/malawi-vibrio-cholerae-2022-2023). The current 2022–2023 outbreak-associated strains differed from historical isolates from Malawi collected in 1989, 1990, 1992, and 1997 by ∼19-45 core-genome SNPs **(Fig. 3)**. Although based on these SNP differences, it seems plausible that the current outbreak may have been derived from the historical Malawi *Vc* strains through stepwise evolution, the historical isolates from Malawi form a distinct phylogenetic cluster from the 2022–2023 and other global phylogeny isolates. These historical isolates collected in 1997 showed a high phylogenetic affinity with sequenced isolates from Zambia, Tanzania, and Democratic Republic of Congo (∼1 core-genome SNP), while those collected in 1989, 1990, and 1992 were more similar to the isolates from Mozambique (∼19 core-genome SNPs), South Africa (∼20 core-genome SNPs), and Angola (∼20 core-genome SNPs). Collectively, unlike the 2022–2023 outbreak isolates, which were genetically closest to the T12 transmission lineage, the historical Malawi *Vc* isolates clustered with isolates belonging to the T1 transmission lineage^7^.

These findings provide compelling evidence for the recent emergence of the 2022–2023 Malawi *Vc* isolates through international importations rather than derivation from the locally circulating strains through stepwise evolution.

### The 2022–2023 outbreak-associated *V. cholerae* isolates from Malawi harbour a diverse set of virulence genes

One of the strongest prevailing hypotheses for the occurrence of the largest cholera outbreak in Malawi from 2022–2023 primarily pertains to tropical cyclones Ana and Gombe that occurred in early 2022. However, although similar cyclones occurred in some years prior to 2022, the reported cholera cases and deaths during each corresponding seasonal outbreak were significantly lower than seen during the 2022–2023 outbreak^28^. Understanding the environmental and pathogen-specific factors which may contribute to the high incidence of cholera cases and deaths during the ongoing 2022–2023 cholera outbreak in Malawi is critical to minimise the impact of future outbreaks. We hypothesised that the presence of a diverse set of virulence genes in the 2022–2023 outbreak-associated *Vc* isolates, and the absence in the historical isolates from Malawi might promote their pathogenicity. Our hypothesis seemed credible as the 2022–2023 *Vc* isolates from Malawi were inferred to be of Asian origin based on recent studies of African *Vc* isolates^7^ and the high genetic similarity of the Malawi isolates to those linked with large outbreaks and epidemics elsewhere, including Haiti in 2010^74^ and Yemen in 2016–2017^75^. To gain insights into the virulence and pathogenicity profiles of the 2022–2023 Malawi cholera outbreak *Vc* isolates, we investigated the genome-wide presence and absence patterns of genes and MGEs known to be associated with the pathogenicity and virulence of *Vc* strains.

We first assessed the distribution of the VC2346 gene, widely considered to be a specific non-transferrable marker in the *Vc* genetic backbone for identifying circulating 7PET wave *Vc* strains^55^ **(Fig. 4a)**. We found this 7PET genetic marker in all 2022–2023 outbreak-associated and historical ST69 *Vc* isolates from the late 80s and 90s in Malawi. This observation confirmed that the 7PET clone has circulated in Malawi for over three decades, consistent with the phylogenetic analysis **(Supplementary Data 4,5)**. In contrast, both non-outbreak-associated *Vc* isolates sampled during the 2022–2023 outbreak (ST40 and ST635) did not harbour the 7PET marker gene.

**Figure 4:**
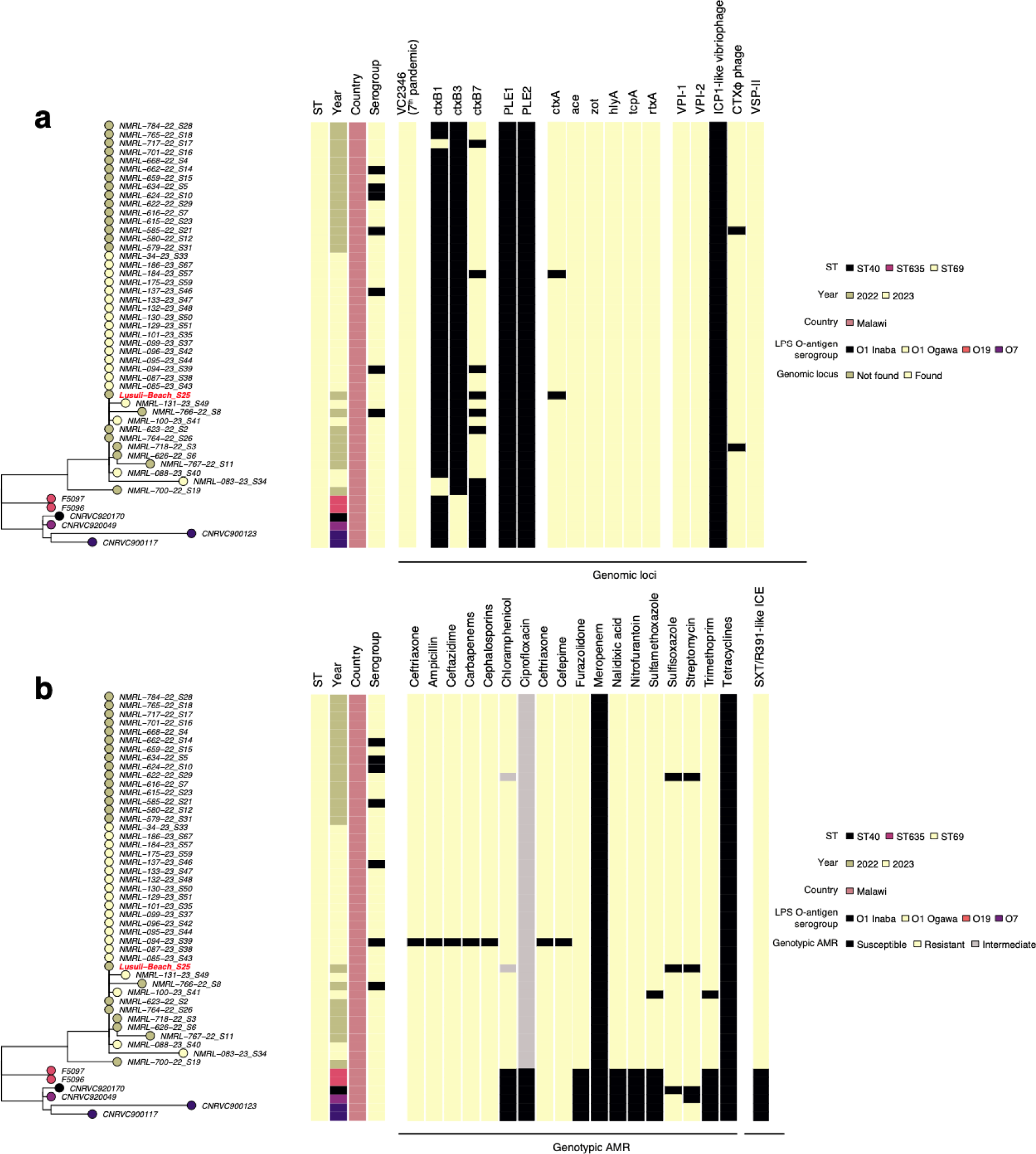
Phylogenetic distribution of the virulence factors and genotypic antimicrobial resistance. **(a)** Distribution of virulence genes, pathogenicity islands, and bacteriophages in the 2022–2023 and historical *Vc* isolates from Malawi. **(b)** Distribution of genotypic AMR profiles and presence and absence of the SXT/R391-like ICE in the 2022–2023 and historical *Vc* isolates from Malawi. The circles with different colours at the tip of the phylogeny represent the year of isolation. All the isolates were sampled from human clinical cases except one (coloured in red text), which was obtained from a water sample at a beach in the southern region of Malawi. The phylogeny is annotated by four column colour strips at the tips of each tree representing the sequence type (ST), year of isolation, country, and LPS O-antigen serogroup and/or serotype. The phylogeny was constructed based on the core-genome SNPs identified from the merged alignments of chromosomes 1 and 2, and rooted based on an outgroup *Vibrio mimicus* species, not shown in the tree (see methods).

Next, we analysed the distribution of the biotype-specific genes *ctxB* and *rstR,* which are disseminated via the filamentous lysogenic CTXφ prophage^17, 18^. The *ctxB* gene encodes a cholera toxin (CT) subunit B, which is considered to be the major virulence factor of *Vc,* while the *rstR* gene encodes a repeat sequence transcriptional regulator required for replication and regulation of the phage^17, 76^. All the 2022–2023 outbreak-associated and historical ST69 isolates from Malawi contained the *rstR* gene, while ∼90% (38 of 42 cases) of the clinical isolates and 100% (6 of 6 cases) of the historical isolates harboured *ctxB* gene. Since the *ctxB* gene is co-carried with *ctxA* on the CTXφ prophage, we also assessed the distribution of *ctxA* in the four clinical *Vc* isolates in which we did not find the *ctxB* gene and found that three harboured a *ctxA* gene, while a single isolate did not contain *ctxA* or *ctxB* genes. This observation implied that the correct prevalence of the *ctxB* gene in the 2022–2023 clinical *Vc* isolates may be ∼97% (41 of 42 cases). Interestingly, a single environmental ST69 *Vc* isolate sequenced in the present study showed the absence of both *ctxA* and *ctxB* although it clustered with the rest of the outbreak-associated ST69 *Vc* isolates **(Fig. 4a)**.

Unlike the historical Malawi *Vc* isolates collected in the late 80s and 90s, which contained the typical El Tor genotype *ctxB3* allele, the current outbreak isolates predominantly contained Haitian *ctxB7* variant **(Fig. 4a, Supplementary Data 5)**. The *ctxB7* variant is characteristic of the circulating wave 3 7PET *Vc* strains reported globally, including during the well-known *Vc* epidemics in Haiti^55^ and Yemen^75^. The observed *ctxB3* to *ctxB7* genotype shift, which probably occurred during the past two decades in Malawi, provided further evidence supporting the importation of the 2022–2023 outbreak-associated ST69 isolates rather than via stepwise evolution of the sequenced historical locally circulating ST69 strains from the 1980s and 1990s. We also found a similar distribution of additional CTXφ prophage core genes (*ace* and *zot*) considered to play a role in *Vc* virulence and pathogenicity in the outbreak-associated and historical ST69 isolates, but not in the non-ST69 isolates widely associated with sporadic cholera cases.

The outbreak-associated ST69 isolates also showed the presence of other virulence genes, including the *Vc* heptameric pore-forming hemolysin protein-encoding gene (*hlyA*), ToxR-regulated membrane-associated pore-forming proteins (*ompT* and *ompU*)^77, 78^, flagella-associated cytotoxin (*makA*), mannose-sensitive hemagglutinin adhesin (*mshA*), and a multifunctional RTX toxin family protein (*rtxA*), which is closely linked with the CTX prophage^79^. Besides the CTXφ prophage, all the 2022–2023 outbreak-associated and historical ST69 isolates harboured the *Vc* pathogenicity island 1 (VPI-1), which harbours the second most important *Vc* virulence factor^21^ **(Fig. 4a, Supplementary Data 5,6,7)**. The VPI-1 element encodes *tcpA,* a toxin-coregulated pilus (TCP), a receptor for the CTXφ phage^20, 21^, critical for intestinal colonisation^22^. Unsurprisingly, VPI-1 island was absent in non-ST69 isolates, further highlighting its role in regulating the virulence of outbreak-associated *Vc* strains. Similarly, all the current outbreak and historical ST69 and non-ST69 isolates also harboured the *Vc* pathogenicity island 2 (VPI-2)^80^ and Vibrio seventh pandemic island II (VSP-II) element commonly associated with 7PET *Vc* isolates^23^. The VSP-II of historical *Vc* isolates from Malawi closely resembled the VSP-II sequence of *Vc* isolates isolated in the 1990s (GenBank accession: KU601747), while the 2022– 2023 outbreak isolates harboured a VSP-II similar to that found in recent O1 El Tor *Vc* isolates (GenBank accession: KM660639) **(Supplementary Data 7)**. Furthermore, both the historical and 2022–2023 *Vc* isolates harboured PICI-like elements (PLE), which inhibit host immunity and promote lytic phage infection^81^ **(Fig. 4a)**. These findings suggest that the 2022–2023 *Vc* isolates from Malawi harboured a diverse set of genetic factors known to promote the virulence of *Vc* isolates typically associated with global outbreaks.

### The 2022–2023 outbreak-associated *V. cholerae* isolates show higher AMR and harbour SXT/R391-like ICE than the historical isolates from Malawi

We screened for the presence of genotypic antibiotic resistance for seventeen antibiotics, namely, ceftriaxone (CRO), ampicillin (AMP), ceftazidime (CAZ), carbapenems (CAR), cephalosporins (CEP), chloramphenicol (CHL), ciprofloxacin (CIP), cefepime (CPM), furazolidone, meropenem (MER), nalidixic acid (NA), nitrofurantoin (NIT), sulfonamides (sulfamethoxazole [SXT] and, sulfisoxazole [SXZ]), furazolidone (FZD), streptomycin (STR), trimethoprim (TMP), and tetracyclines (tetracycline [TET] and doxycycline [DOX]) **(Fig. 4b)**. We found higher genotypic resistance in ST69 than in non-ST69 isolates, associated with nine to fifteen antibiotics. The ST69 isolates showed TET/DOX susceptibility; this antibiotic is a recommended treatment option for severe cholera infection to complement the highly effective rehydration therapy using polymer-based or glucose-based rehydration solutions^4, 5^. We also observed intermediate resistance against CIP, which is an alternative treatment option in children in Malawi. Interestingly, the historical ST69 isolates from Malawi showed resistance to fewer antibiotics than the 2022–2023 outbreak isolates, which consistently showed genotypic resistance to the antibiotics mentioned above, except for CIP, TET, and MER **(Fig. 4b, Supplementary Data 8)**.

We first assessed the presence and absence of AMR-conferring genes typically carried on the 100Kb SXT/R391-like ICE originally isolated from a 1993 *Vc* O139 clinical isolate from India^25^ **(Fig. 4b, Supplementary Data 7)**, which was the first MGE with ICE-like properties described in the Gammaproteobacteria^26^. Due to horizontal gene transfer, the SXT/R391 ICE is genetically related to ICE-like MGEs found in other bacterial species, including *Salmonella*^82^ and *Providencia rettgeri*^82^, and harbours a diverse set of genes which confer resistance to STR (*strAB, aph(3’’)-Ib, aph(3’)-IIa,* and *aph(6)-Id*), sulfonamides SXT and SXZ (*sul2*), TMP (*dfrA*), and tetracyclines (*tet(A)* and *tetA(D)*). Nearly all the Malawi 2022–2023 outbreak-associated *Vc* ST69 isolates (∼97%, 42 of 43 cases) harboured *aph(3’’)*-Ib and *aph(6)*-Id, and the dual presence of *strA* and *strB*, which contributed to the STR resistance **(Supplementary Data 8–10)**. We also found these genes in ∼67% (4 of 6 cases) of the historical Malawi ST69 isolates. We noted universal TPM resistance conferred by the *dfrA1* gene in the current outbreak-associated ST69 *Vc* isolates, but both non-ST69 isolates from the current outbreak did not harbour this gene. In contrast, none of the historical ST69 *Vc* isolates from Malawi harboured the *dfrA1* gene; instead, ∼83% (5 of 6 cases) of these isolates showed the presence of the *dfrA15* gene, which was suggested to be a general feature of the African serotype O1 *Vc* isolates^83^. This suggested that the *dfrA1* gene may have been acquired independently elsewhere before the 2010s when identical strains were reported in several countries globally and before the subsequent introduction of the *dfrA1-*positive strains in Malawi.

We also detected sulfonamide resistance genes *sul1* and *sul2* in the historical ST69 isolates from Malawi **(Supplementary Data 8–10)**. However, the Malawi 2022–2023 outbreak ST69 isolates harboured only the *sul2* gene. Similarly, we identified the SXT-like ICE-borne CHL resistance gene *floR* in the current outbreak-associated *Vc* ST69 isolates but not the historical isolates. Additionally, we found additional CHL resistance genes, *catB9* genes in the current outbreak and historical ST69 isolates, with the latter also containing the *catA1* gene. Interestingly, we did not find *tet(A)* and *tetA(D)* resistance genes carried by the SXT-like ICEs in all the isolates from Malawi, which suggested universal susceptibility of the *Vc* isolates to TET/DOX, consistent with AMR findings in outbreak-associated serotype O1 isolates from other African countries, including from Kenya^84^, Central African Republic^50^, and Algeria^51^. We observed universal CAR genotypic resistance in the current outbreak and historical ST69 isolates from Malawi mediated by the *varG* gene^85^. Apart from *blaCARB4* CAR resistance-conferring gene detected in ∼67% (4 of 6 cases) of the historical isolates from Malawi, there was no presence of other known CAR resistance genes, including *blaCARB*, *blaCMY*, *blaCTX-M, blaDHA*, *blaOXA*, *blaPER*, *blaSCO*, *blaSHV*, and *blaTEM*.

We then performed an in-depth analysis of the 2022–2023 outbreak and historical *Vc* isolates from Malawi to identify the SXT/R391-like ICEs characteristic of global 7PET wave 3 *Vc* isolates^8^ **(Fig. 4b, Supplementary Data 8–9)**. We mapped *k-*mers of the *Vc* isolates against reference SXT/R391-like ICE sequences, namely, the original SXT/R391^25^, ICEVchban5^27^, ICEVchind4^27^, ICEVchind5^27^, ICEVchmex1^27^, ICEVflInd1^27^, ICEVchHai1^86^, ICE^TET^ ^87^ and ICE^GEN^ ^87^. These SXT-like ICEs exhibit some homology with each other, with the ICE^GEN^, ICEVchHai1, ICEVchind5, and ICEVchban5 showing the highest homology, suggesting they are nearly identical SXT/R391 elements **(Supplementary Data 7)**. The rest of the SXT/R391-like elements showed considerable genetic divergence, which suggested they represent distinct allelic versions of the original SXT/R391 element.

The *k-*mer-based mapping analysis showed the absence of all the versions of the SXT/R391-like ICEs in the historical and non-ST69 *Vc* isolates collected during the 2022–2023 outbreak in Malawi consistent with the distribution of the ICE-borne AMR genes. In contrast, all the outbreak-associated ST69 isolates contained an SXT/R391-like element genetically closest to the ICE^GEN^, ICEVchHai1, ICEVchind5, and ICEVchban5 ICEs **(Fig. 4b, Supplementary Data 11)**. All the outbreak-associated ST69 *Vc* isolates showed the highest mapping coverage against the ICEVchban5, ICEVchHai1, and ICE^GEN^ reference sequences **(Supplementary Data 11)**. These ICEs are commonly found in outbreak-associated *Vc* isolates of Asian origin, including those reported in Africa, Bangladesh, India, Nepal, Yemen, and Haiti^7, 8, 54, 56, 70^.

Together, these findings suggest that the *Vc* strains responsible for the 2022–2023 cholera outbreak in Malawi are genotypically resistant to more antibiotics than the historical *Vc* isolates from Malawi due to the presence of the ICE^GEN^/ICEVchHai1/ICEVchind5 SXT/R391-like ICEs. However, the absence of TET/DOX resistance genes supports the continued use of tetracyclines in Malawi as a first-line antibiotic for treating cholera-affected patients.

## Discussion

Cholera outbreaks and epidemics associated with the seventh pandemic, exemplified by those in 2010–2011 Haiti^88^ and 2016–2017 Yemen^9^, continue to cause a significant diarrhoea-associated disease burden globally, especially in endemic settings in the low-and-middle-income countries (LMIC)^36, 89, 90^. Here, we describe that the deadliest, and ongoing cholera outbreak in Malawi’s history, which started in March 2022, is caused by the O1 serogroup El Tor biovar. We identified co-circulation of the Ogawa and Inaba serotypes, highlighting potential serotype switching as seen elsewhere^56, 65^, with a predominance of the Ogawa serotype, which appears the dominant serotype in the African region^7, 49–53^ and other continents^54–58^. In contrast to the previous seasonal cholera outbreaks in Malawi^28, 36^, the 2022–2023 outbreak caused cholera cases in all the districts, with the persistence of cases even during the dry season throughout 2022 in some districts, including Blantyre, Chikwawa, and Neno. We speculate that the persistence of cases in these districts, particularly Blantyre, which is a major city and transportation hub linking the southern region to other districts in Malawi, may have contributed to the human-to-human *Vc* transmission to other districts following the onset of the outbreak in Nsanje district.

Despite identifying diverse *Vc* lineages, associated with three STs, the 2022–2023 outbreak was primarily driven by the ST69 lineage, which showed the highest similarity to Asian and Haiti isolates, suggesting a potential Asian origin, consistent with previous phylogeography work^7, 8, 54^. Unlike the historical isolates that circulated in Malawi during the late 1980s and 1990s, the 2022–2023 isolates demonstrate higher AMR, which is partly driven by the acquisition of the ICE^GEN^/ICEVchHai1/ICEVchind5 SXT/R391-like ICEs^8^. However, the acquired SXT/R391-like ICEs did not harbour tetracycline-resistance-conferring *tetA*, which supports the continued use of tetracyclines as the currently recommended antibiotic for treating cholera in Malawi. These findings demonstrate that the 2022–2023 cholera outbreak in Malawi is caused by recently imported multi-drug resistant ST69 O1 *Vc* strains.

Previous studies in Africa and elsewhere have suggested that some enteric bacteria may cause cholera-like diarrhoea infections, which may be incorrectly attributed to *Vc* during cholera outbreaks^37–41^. Although our study was not designed to test the proportion of cholera-like cases attributed to non-*Vc* enteric pathogens, our genomic analysis identified these bacterial pathogens in ∼28% of the suspected *Vc* isolates collected from patients presenting with cholera-like symptoms. These findings might suggest that a considerable number of profuse diarrhoea cases may have been incorrectly attributed to *Vc*. This is especially true in sub-Saharan African settings, such as Malawi, where a full battery of readily available tests to accurately identify a range of non-*Vc* enteropathogens may not be routinely available. These findings have repercussions towards patient management and could explain, in part, why the 2022–2023 outbreak persisted. Considering that all patients presenting with cholera-like symptoms at the hospitals are managed in the cholera treatment units (CTUs), there is an opportunity for *Vc* transmission between the clinically isolated definitive *Vc*-positive and *Vc-*negative patients presenting with cholera-like symptoms. Since up to 25% of those infected with *Vc* shed the bacteria asymptomatically^91^, those that contracted *Vc* in CTUs likely facilitated repeated reseeding of *Vc* into the community following an incubation period^92^. Although we recovered several enteric bacteria in *Vc*-negative samples, *Aeromonas caviae,* which mimics *Vc* enteropathy^37–41, 44–47^, may be the most common non-*Vc* bacterial pathogen in these incorrectly diagnosed cholera-like diarrhoea cases. Our literature search revealed no epidemiological studies on the fraction of cholera-like cases attributable to *Aeromonas spp.* in sub-Saharan Africa. Thus, to inform public health responses, further studies are required to understand the proportion of cholera-like cases attributable to *Vc* and other enteropathogens during cholera outbreaks.

One of the plausible hypotheses that could explain the increased risk of cholera transmission are humanitarian crises that disrupt water and sanitation systems displace populations towards inadequate and overcrowded camps, and climatic conditions such as flooding due to cyclones. Malawi has experienced three tropical cyclones since January 2022 (Ana in January 2022, Gombe in March 2022 and Freddy in March 2023). The first cholera wave was reported in the Nsanje district in March 2022, which coincided with the occurrence of tropical cyclone Gombe in Malawi, which caused floods in the lower Shire area in the southern region of Malawi. It is likely that the disruption to water supplies and sanitation facilities, as well as overcrowding in camps caused by tropical cyclone Ana, which occurred in the southern region of Malawi approximately two months before cyclone Gombe, created a conducive environment to kickstart the cholera outbreak. However, human-to-human transmission events likely contributed to the spread of cholera across the country throughout the dry season in 2022, which is strikingly different from the past years, where cholera outbreaks have occurred only during the rainy seasons. Notably, cholera cases in all districts of the northern region peaked during the hot months in 2022, contrary to the central region and most districts in the southern region (except Nsanje, Neno, Chikwawa, and Blantyre), where most of the cases were reported after November 2022 subsequently peaking in early 2023. Surprisingly, the occurrence of cyclone Freddy in early 2023 coincided with a reduction in cholera cases in all districts except the two lower shire river districts, Chikwawa and Nsanje, which are typically affected by floods and tend to kickstart cholera outbreaks in Malawi. Our findings suggest that the occurrence of the tropical cyclones in Malawi may have precipitated the deadliest cholera outbreak in Malawi, highlighting the impact of climatic changes on the risk of cholera and possibly other infectious diseases.

Despite the occurrence of previous cyclones in Malawi, the number of cholera cases has never reached the magnitude seen during the 2022–2023 outbreak. Before 2022, the two largest cholera outbreaks in Malawi occurred between 1998–1999 (CFR=3.4%; 860 of 25,000 cases) and 2001– 2002 (CFR=2.3%;968/33,546)^28^. Therefore, we hypothesised that in addition to the climatic conditions conducive to *Vc* transmission and infection, additional *Vc*-specific factors might explain the scale of the 2022–2023 cholera outbreak in Malawi. Our comparative genomic analysis of the 2022–2023 and historical *Vc* isolates from Malawi in the context of global *Vc* sequences revealed two potential independent introductions of the 2022–2023 strains into Malawi. The close genetic similarity of the Malawi isolates to those from Asian countries, including Bangladesh, India, Yemen, and also Haiti, which were originally imported from Asia^54^, suggested that the 2022–2023 outbreak ST69 clone, serogroup O1 strains may have also originated from Asia^7, 8^. Uniquely, the 2022–2023 outbreak ST69 *Vc* isolates appear to be associated with higher AMR than the historical isolates from Malawi, partly due to the acquisition of the ICE^GEN^/ICEVchHai1/ICEVchind5 SXT/R391-like ICEs^8^, which highlights the growing public health concern in this cholera endemic setting. For life-threatening acute infections such as cholera, AMR-associated *Vc* strains could lead to worse clinical outcomes and higher healthcare costs due to longer hospital stays. Although MDR *Vc* strains resistant to tetracyclines due to the presence of an IncA/C2 plasmid have been recently identified in Zimbabwe^93^, a nearby country, and other African settings^61, 94^ as well as globally^95–98^, our genomic analysis suggests that these strains have not yet established themselves in Malawi. In terms of the virulence profiles, the 2022–2023 outbreak-associated O1 *Vc* strains in Malawi mostly exhibit similar characteristics to the historical Malawian *Vc* isolates. The major distinguishing genetic characteristic of the 2022–2023 isolates from the historical isolates is the presence of a different version of the VSP-II pathogenicity island and *ctxB7* allele, which may partly explain the observed transmission and virulence of the 2022–2023 O1 strains in Malawi. These findings show the importance of elucidating the genetic makeup of *Vc* strains to understand the evolution and spread of the *Vc* strains and suggest that genomic surveillance of *Vc* strains in endemic settings could be an important tool, especially for monitoring the emergence and spread of antimicrobial resistance. Such surveillance may be particularly useful to understand the *Vc* strain dynamics following the rollout of oral cholera vaccines (OCV) in Malawi^99^ and other settings^100, 101^. Collectively, these findings highlight the need for robust real-time genomic surveillance to monitor the spread of AMR and virulent strains, and the impact of vaccination on *Vc* strains to inform strategies for controlling and preventing cholera outbreaks.

We acknowledge some limitations. First, the burden of cholera during the 2022–2023 outbreak might have been underestimated due to underreporting of cases because of the unavailability of diagnostic tests, especially in rural settings, and negative healthcare-seeking behaviour, mostly among those presenting with mild disease. Second, we did not employ a full battery of assays required to accurately identify non-*Vc* enteropathogens, which may cause cholera-like symptoms. However, the identification of non-*Vc* bacterial isolates associated with cholera-like diarrhoea has been reported even in high-income settings, suggesting that the isolation of these bacteria, especially *Aeromonas spp.,* may carry clinical relevance. Therefore, we recommend further studies to investigate the contribution of non-*Vc* bacteria in cholera-like diarrhoea during seasonal cholera outbreaks in sub-Saharan Africa. Third, there are limited genotypic characteristics of *Vc* strains in Malawi, other African, and other LMIC countries beyond the country-specific case counts tracked by the WHO. In this study, we had access to only a few contextual *Vc* genomes from Malawi collected in the late 1980s and 1990s, which limited our ability to determine temporal changes in the distribution of *Vc* strains leading to the 2022–2023 outbreak. Consequently, whole genome sequences generated in our study will start to close this knowledge gap and will be critical in providing the much-needed context to understand the origin of *Vc* strains associated with future outbreaks in Malawi. Fourth, due to the small number of sequenced genomes, we could not compare the characteristics of *Vc* strains collected from different districts or regions and temporal scales in Malawi and assess their association with the clinical characteristics, including disease incidence and CFR. Improved disease surveillance systems, particularly sample collection, preservation, and tracking, are critical to generating robust genomic and epidemiological insights in Malawi. Furthermore, we did not perform phylogeography and phylodynamic analyses in this study. Follow-up work should conduct these more elaborate phylogenomic analyses using BEAST^102^ and other tools^103–105^, to estimate the frequency and timing of *Vc* O1 7PET lineage into Malawi. The use of long-read sequencing in future studies would be useful to gain better insights into the structural variation in the *Vc* pathogenicity islands, ICEs, and prophage sequences, beyond variation at the SNP level.

Our study has provided an early snapshot of the genomic characteristics associated with the 2022–2023 *Vc* outbreak in Malawi, the deadliest cholera outbreak ever recorded in the country. Our whole-genome sequencing of *Vc* isolates collected across Malawi shows that the ongoing cholera outbreak is driven by imported late wave 3 global 7PET ST69 lineage strains, rather than strains derived through stepwise evolution from the historical local serogroup O1 *Vc* strains, predominantly expressing serotype Ogawa and, to a smaller extent, the Inaba serotype. This work highlights a concerted locally-driven genomic surveillance effort, with support from international partners, to understand the genomic epidemiology of *Vc* strains linked with the 2022–2023 outbreak. Continued molecular and genomic surveillance in Malawi and the region will be crucial to understand long-term strain dynamics, including the impact of oral cholera vaccines, AMR, and the geographical spread of *Vc*.

## Methods

### Ethical approval

This work was conducted according to the guidelines of the Declaration of Helsinki and was approved by the National Health Sciences Research Committee, Lilongwe, Malawi (Protocol #867) and the Research Ethics Committee of the University of Liverpool, Liverpool, UK (000490) under the Diarrhoea Surveillance study, and the College of Medicine Ethics Committee (COMREC, Protocol #P.10/22/3790) under the NIHR Global Health Research Group on Gastrointestinal Infections: Facilitating the Introduction and Evaluation of Vaccines for Enteric Diseases in Children in Eastern and Southern sub-Saharan Africa study.

### Analysis of Malawian cholera cases and deaths

We analysed the case and deaths data obtained from the Public Health Institute of Malawi, Malawi Ministry of Health (MoH) data on May 20, 2023 (https://cholera.health.gov.mw/surveillance) using the dplyr (version 1.0.8) package for data wrangling (https://github.com/tidyverse/dplyr) in R (version 4.0.3) (https://www.R-project.org/). We calculated the incidence of cases and deaths by dividing the total number of cases reported in Malawi or per district by the population size multiplied by 100,000. We used the population sizes per district based on data reported by the National Statistics Office of Malawi for the 2018 population census (http://www.nsomalawi.mw/). To calculate the CFR during the course of the 2022–2023 cholera outbreak, we first obtained the number of cases and deaths using a sliding window of 21 days using the runner (version 0.3.7) package (https://CRAN.R-project.org/package=runner) and then calculated the percentage of deaths during each window. We plotted the number and incidence of cases and deaths per district using sf (version 1.0.7), rnaturalearth (version 0.3.2), rnaturalearthdata (version 0.1.0), and rnaturalearthhires (version 0.2.1) packages in R (version 4.0.3) (https://www.R-project.org/).

### Specimen collection and preparation

Since surveillance of *Vc*, especially the preservation of clinical isolates is not routinely undertaken in all districts in Malawi, we could not systematically select representative samples for microbiological examination. Therefore, we used a convenience sampling method. Faecal specimens (liquid stools) were collected in clean unchlorinated disposable containers from patients presenting with cholera-like symptoms, including profuse watery diarrhoea and vomiting, at Cholera Treatment Units (CTUs) in Malawi from March 2022 to February 2023. A stool-soaked rectal swab was placed into Cary-Blair transport medium (Oxoid, Thermo Fisher Scientific, USA) and transported to the hospital laboratories within two hours of collection. Once the stool samples arrived in the laboratory, a cholera Rapid Diagnostic Test (RDT) and culture were performed. In addition, a stool-soaked swab from each sample was then inoculated in Alkaline Peptone Water (APW) (Becton Dickinson, UK) for enrichment, which was kept between 4–6 hours at ambient temperature prior to RDT and culture. We did not use clinical metadata related to the patients, and all isolate identifiers were de-identified; therefore, additional institutional review board approval was not required.

### A rapid diagnostic test procedure for *V. cholerae*

Testing was performed by qualified and well-trained laboratory technologists from the District Hospital Laboratories and the Malawi National Microbiology Reference Laboratory. In brief, Crystal VC Ag O1/O139 rapid diagnostic testing kits were used for testing samples and interpreting results by following the manufacturer’s instructions (Arkray, Japan). About four drops of watery stool were transferred into a sample processing vial (pre-filled with 1 ml of sample diluent buffer), and the Crystal VC strip was dipped into it for at least 15 minutes; the test line and/or control line appeared as a red colour. The appearance of both lines indicated that the sample was positive for *Vc* serogroup O1; the appearance of only the control line but not the test line indicated a negative result for the test.

### Identification of *V. cholerae*

Sample collection was restricted to patients with known antibiotic consumption history. Strict laboratory safety precautions were followed at all times when working with suspected cholera specimens. Appropriate personal protective equipment (PPEs) was always worn, and standard precautions were followed for handling and disposing of biological materials. The stool samples were streaked (cultured) on Thiosulphate Citrate Bile Sucrose Agar (TCBS) (Becton Dickinson, UK) media after four hours of incubation in APW, and then incubated at 37°C for 18–24 hours. After 18–24 hours of incubation on TCBS, large (2–4 mm in diameter) slightly flattened, yellow colonies with opaque centres and translucent peripheries were examined, suggestive of *Vc*. Well-isolated (pure) single yellow colonies were picked and streaked on Nutrient Agar (Becton Dickinson, UK) and incubated at 37°C for 24 hours. Presumptive identification of *V. cholerae* was made with a positive oxidase biochemical test.

### Bacterial DNA extraction

Genomic DNA of the suspected *Vc* colonies was extracted using QIAamp DNA Mini Kit (Qiagen, Germany), at the National Microbiology Reference Laboratory within the Public Health Institute of Malawi (PHIM). The extracted nucleic acid material was shipped on dry ice to the University of the Free State-Next Generation Sequencing (UFS-NGS) Unit, Bloemfontein, South Africa, for library preparations and whole genome sequencing.

### Library preparation

The bacterial DNA samples were quantified on a Qubit fluorometer using a High Sensitivity dsDNA Assay kit (Thermo Fisher Scientific, USA). The obtained DNA concentrations were normalised to 0.2–0.3ng/µl by diluting with an elution buffer (Qiagen, Germany). Genomic libraries were prepared with the Nextera XT DNA Library preparations kit (Illumina, USA). Normalized DNA was enzymatically fragmented and simultaneously tagged with Illumina sequencing adapters, and each sample of the fragmented DNA was uniquely indexed using Nextera DNA CD Indexes (Illumina, USA). This was followed by library size selection and purification using Ampure XP magnetic beads (Beckman Coulter, USA) and freshly prepared 80% ethanol.

### Library validation and sequencing

The quality of the libraries and fragment size distribution was assessed on Agilent 2100 Bioanalyzer using the dsDNA High Sensitivity Assay kit (Agilent Technologies, USA), and the average fragment size obtained was 600 bp. The purified libraries were fluorometrically quantified on Qubit 3.0 fluorometer, followed by normalisation to equimolar concentrations of 4 nM. Normalized libraries were pooled into a clean 1.5 ml tube. The library pool was denatured with a freshly prepared 0.2 N sodium hydroxide (NaOH), followed by dilution with a hybridisation buffer (HT1) to a final concentration of 8 pM. Lastly, the library was spiked with 0.5% PhiX sequencing control (20 pM), and DNA sequencing was performed on a MiSeq platform (Illumina) for 600 cycles, using a V3 reagent kit (Illumina, USA), to generate 2×301 pb paired-end reads. After sequencing, the libraries were demultiplexed on an instrument based on the unique index sequences and separate fastq files that were generated for each library. In summary, nearly 80% of the data achieved a Phred score of at least Q30 and a cluster density of 926 K/mm^2^ was achieved, with 88.7% of the clusters having passed the filter. We used cutadapt (version v4.4) to trim adapters from the raw sequence reads^106^.

### Comparative genomic analysis

To generate the whole-genome phylogeny of the *Vc* isolates, we first mapped the assembled contigs of each *Vc* isolate and an outgroup *Vibrio mimicus* genome Y4 strain (GenBank accessions: CP077425 and CP077426) against a merged reference sequence of *Vc* O1 biovar El or strain N16961 chromosome 1 (GenBank accession: AE003852) and 2 (GenBank accession: AE003853) separated by ambiguous bases (Ns) using Snippy (version 4.6.0) (https://github.com/tseemann/snippy). We used the “–ctgs” option to determine SNPs between the assembled contigs and the merged reference genome. We then compared the merged reference sequence against nucleotide sequences of known *Vc* pathogenicity islands, prophages, and ICEs to identify their genomic coordinates using BLASTN (version 2.12.0+)^107^. We then masked the genomic regions containing pathogenicity islands, prophages, and ICEs using a custom Python script developed at the Wellcome Sanger Institute (https://github.com/sanger-pathogens/remove_blocks_from_aln). We identified variable sites in the whole-genome alignment containing SNPs using snp-sites (version 2.5.1)^108^. Next, we converted all ambiguous bases (N) to gaps (–) in the generated alignment of variable sites to exclude positions with >0.05% gaps using trimAl (version 1.4.rev15)^109^, which resulted in the final core-genome SNP alignment used for phylogenetic analysis. We selected this threshold of 0.05% to include as many variable genomic sites in the *Vc* genome which may have been absent in the *V. mimicus* outgroup species.

We constructed a maximum likelihood core-genome phylogeny of the Malawi *Vc* isolates using FastTree (version 2.1.11) with 50 threads using the generalised time reversible model. To root the generated phylogeny on the branch separating the *Vc* isolates and the outgroup *V. mimicus* genome, we used the “root” function in the APE package (version 5.6.2)^110^. Next, we ladderised the outgroup-rooted phylogeny using “ladderize” function in APE (version 5.6.2)^110^, and then dropped the outgroup taxon from the phylogeny, for clarity, using the “drop.tip” function in APE (version 5.6.2)^110^. We exported a Newick file of the resulting phylogenetic tree and visually explored it using Taxonium, a web-based phylogenetic visualisation tool (https://taxonium.org/)^111^. To show the genetic relatedness of specific *Vc* isolates, we pruned the global phylogeny and performed additional visualisation using the APE package (version 5.6.2)^110^ and RCandy (version 1.0.0)^112^. We determined the number of core-genome SNPs distinguishing specific *Vc* isolates using snp-dists (version 0.7.0) (https://github.com/tseemann/snp-dists). To place the Malawian *Vc* genomes in the global context, we obtained a large collection of globally diverse *Vc* isolates worldwide available in the VibrioWatch implemented in the PathogenWatch web tool (https://pathogen.watch/).

To assess the presence and absence of genes used for species identification and biotyping, and those encoding virulence factors and AMR, we used ABRicate (version 1.0.1) (https://github.com/tseemann/abricate). We compared each genome to a reference database of genes obtained from the NCBI AMRFinderPlus database and the virulence factor database (VFDB)^113^. We used a custom database of the genes used for biotyping, AMR, and pandemic lineage and species identification in the CholeraeFinder tool (https://cge.cbs.dtu.dk/services/CholeraeFinder). We obtained these genes from the tool’s repository (https://bitbucket.org/genomicepidemiology/choleraefinder_db/src/master/). The presence of the VC2346 gene (GenBank accession: AE003852) was used to determine pandemic *Vc* lineages while *rstR* classical (GenBank accession: KJ023707), *rstR* El Tor (GenBank accession: AE003852), *tcpA* classical (GenBank accession: M33514), *tcpA* classical (GenBank accession: CP001235), *tcpA* El Tor wave 3 (GenBank accession: AF325734). We identified specific *ctxB* alleles of the *Vc* isolates by comparing the assemblies to *ctxB* alleles, namely, *ctxB1* (GenBank accession: CP001235), *ctxB3* (GenBank accession: AE003852), and *ctxB7* (GenBank accession: JN806157). Additionally, we also inferred the genotypic AMR of the *Vc* isolates using VibrioWatch implemented in the PathogenWatch web tool (https://pathogen.watch/).

We inferred serogroups and serotypes of the *Vc* isolates using an *in silico* genomic-based approach based on mapping nucleotide sequence *k-*mers of each isolate against all known reference LPS O-antigen biosynthesis gene cluster sequences to determine the specific serogroups and serotypes of our isolates^48^. We inferred the serotype as the reference LPS O-antigen biosynthesis gene cluster with the highest sequence coverage than the rest of the LPS O-antigen sequences.

Lastly, we checked the presence and absence of MGEs, including pathogenicity islands, prophages, and ICEs. Due to the genomic variability of these sequences, driven by horizontal gene transfer, and their variably larger sizes compared to individual genes, we inferred their presence in the genomes based on *k-*mer sequences using KMA (version 1.4.12a)^114^. We specified the following flags when running KMA “-ef -dense -ex_mode -mct 1.0 -1t1 -mrs 0.1” to improve the query coverage so that the results were consistent with those based on visual inspection of the comparison of each genome and the MGEs using BLASTN (version 2.12.0+)^107^ and ACT (version 18.1.0)^115^. We mapped *k-*mers of each *Vc* isolate against reference SXT-like ICE sequences, namely, the original SXT (GenBank accession: KJ817376), ICEVchban5 (GenBank accession: GQ463140), ICEVchind4 (GenBank accession: GQ463141), ICEVchind5 (GenBank accession: GQ463142), ICEVchmex1 (GenBank accession: GQ463143), ICEVflInd1 (GenBank accession: GQ463144), ICEVchHai1 (JN648379), ICE^TET^ (GenBank accession: MK165649), and ICE^GEN^ (GenBank accession: MK165650). We used minimum cut-offs of 40% and 80% for the query sequence coverage and identity, respectively.

## Data Availability

All data produced in the present work are contained in the manuscript and upon reasonable request to the authors

## Code availability

We have described all the tools and methods used for the analysis in the Material and Methods sections.

## Data availability

We have deposited the whole-genome sequence data for the study isolates in the National Center for Biotechnology Information (NCBI) Sequence Read Archive (BioProject number: PRJNA974496). The accession numbers and other metadata for our isolates are available in the **Supplementary Data 1** file. All the other data supporting the findings of this study are described in this paper or are available as part of the supplementary material. Any additional data not included in the manuscript is available upon reasonable request to the authors.

## Author contributions

C.C., I.C., W.K., N.A.C., and K.C.J., conceived and designed the study. I.C., W.K., and K.C.J. performed sample selection. I.C. and W.K. performed microbiological work. M.M.N. carried out whole-genome sequencing of the samples. K.C.J., I.C., B.M., C.M., Q.D., and U.L.M. assisted with compiling cholera case and death data. C.C., I.C., and K.C.J. analysed and interpreted the data. S.N. and D.K. assisted with data management and analysis. C.C. and K.C.J. wrote the initial draft of the paper. C.C., I.C., P.M., W.K., C.M., U.L.M., J.B-B, K.C.Jambo, B.M., W.Kapindula, P.B., R.J.M, S.N., D.K., A.K., L.N., A.D.S., A.W.K., M.M.N., D.H., N.F., M.K., Q.D., C.L.M., N.A.C., and K.C.J. reviewed and approved the manuscript.

## Acknowledgements

We acknowledge the support of the Ministry of Health and clinical staff in Malawi, who cared for the patients presenting with cholera-like diarrhoea and collected the samples used in this study. We thank the National Microbiology Reference Laboratory (NMRL) at the Public Health Institute of Malawi (PHIM) for coordinating sample collection and compiling data on the number of cholera cases and deaths in Malawi. We also thank the Next Generation Sequencing Unit and Division of Virology at the University of the Free State for timely genome sequencing services and technical support, especially Milton Mogotsi and the wet-lab team. We are also grateful to Dr Florent Lassalle at the Wellcome Sanger Institute for providing advice regarding genomic-based serotyping of *Vc* and kindly sharing a reference dataset of LPS O-antigen loci for all known *Vc* serogroups. Finally, we thank the Yale Center for Research Computing at Yale University and Wellcome Sanger Institute for providing computational resources used for the bioinformatic analysis.

## Funding

This research was funded by the UK National Institute for Health and Care Research (NIHR) Global Health Research Group on Gastrointestinal Infections (grant number: NIHR133066) using UK Aid from the UK Government to support global health research; and the Bill and Melinda Gates Foundation (Grant number: Investment number: INV-046917). N.A.C. is an NIHR Senior Investigator (NIHR203756). N.A.C., D.H., and K.C.J. are affiliated with the NIHR Health Protection Research Unit in Gastrointestinal Infections at the University of Liverpool, a partnership with the UK Health Security Agency (UKHSA), in collaboration with the University of Warwick. The funders had no role in the study design, data collection and interpretation, or the decision to submit the work for publication. The authors did not receive any financial support or other forms of reward related to the development of the manuscript. Therefore, the findings and conclusions in this report are those of the authors and do not necessarily represent the formal position of the funders. The views expressed are those of the author(s) and not necessarily those of the NIHR, the Department of Health and Social Care or the UK Health Security Agency or the UK government.

## Competing interests

The authors declare no competing financial or non-financial interests.

## Notes

### Competing Interest Statement

The authors have declared no competing interest.

