## Supplementary material for "Genomic insights into the 2022–2023 *Vibrio cholerae* outbreak in Malawi"

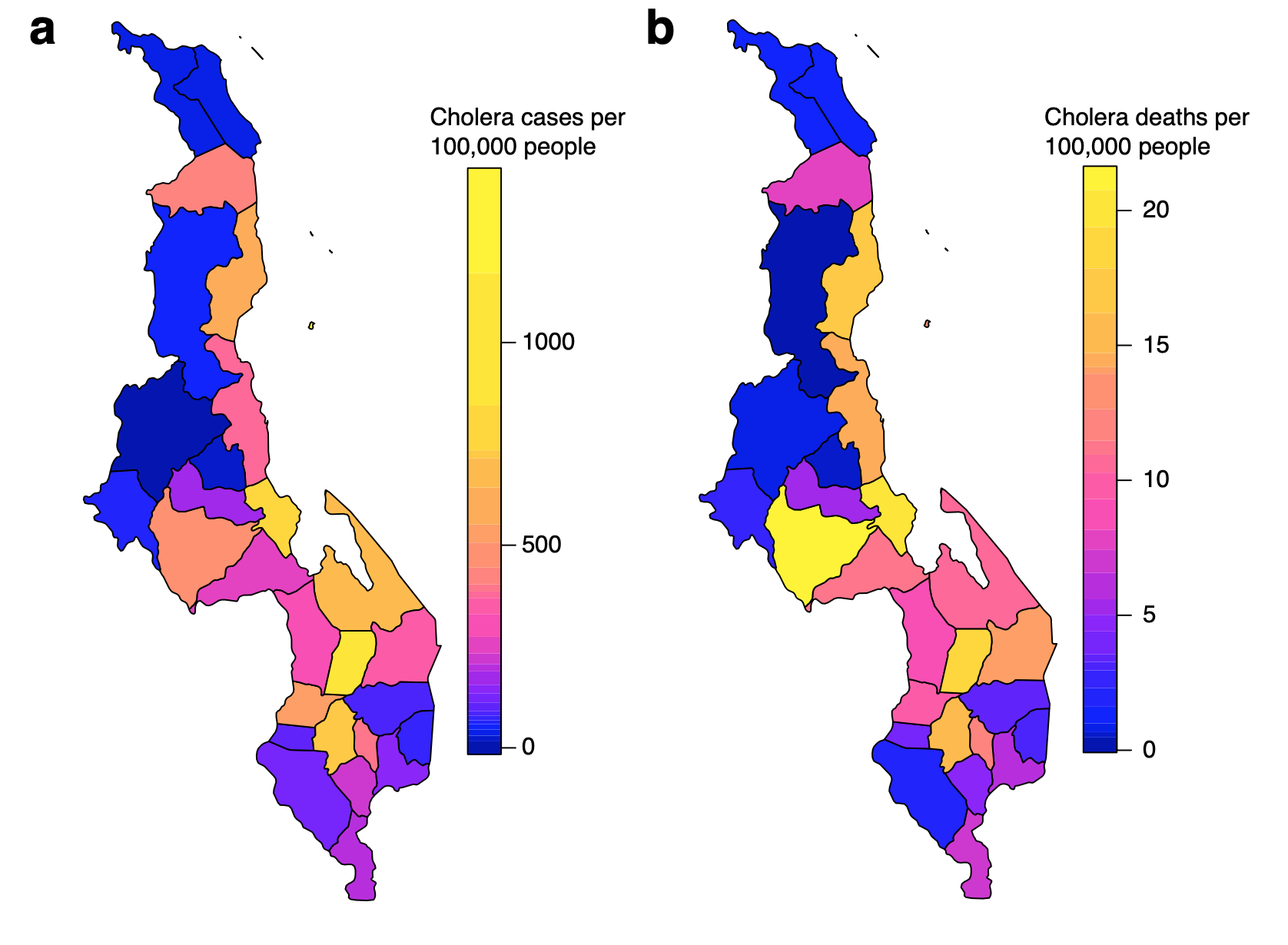

**Supplementary Fig. 1: Incidence of cases and deaths during the 2022–2023 cholera outbreak in Malawi (data from January, 2022 to May, 2023). (a)** The overall incidence of cholera cases per 100,000 people. **(b)** The overall incidence of cholera deaths per 100,000 people.

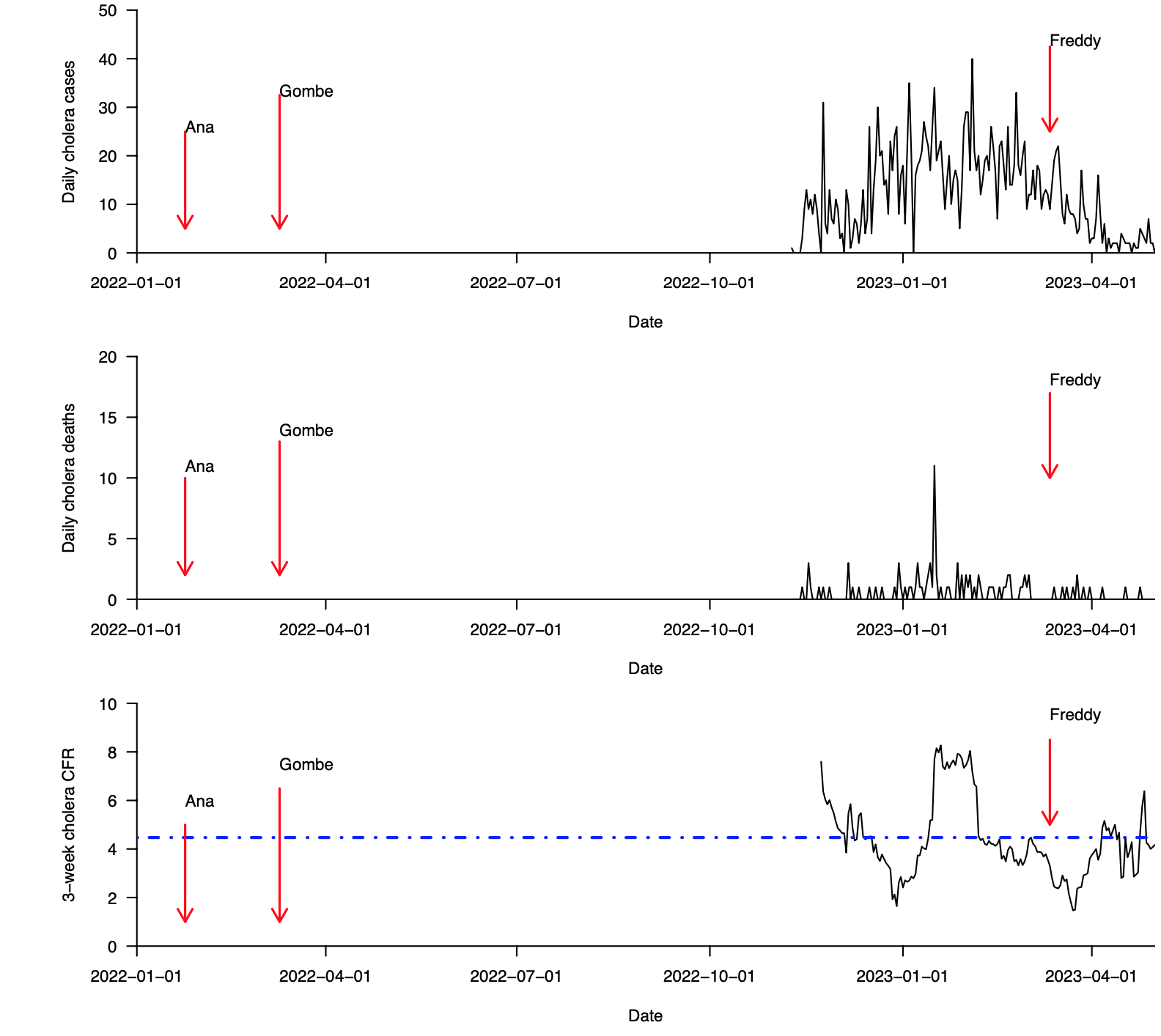

**Supplementary Fig. 2: Cases, deaths, and case fatality ratio during the 2022–2023 cholera outbreak in Dedza district in Malawi (data from January, 2022 to May, 2023). (a)** Total daily cholera cases. **(b)** Total daily cholera deaths. **(c)** Overall cholera case fatality ratio (CFR) based on a 21-day sliding window. The 21-day sliding window was chosen to obtain stable estimates of the CFR, especially during weeks and months with few reported cholera cases. Data were obtained from the Public Health Institute of Malawi, Malawi Ministry of Health (MoH) data on May 20, 2023 [(https://cholera.health.gov.mw/surveillance](https://cholera.health.gov.mw/surveillance)).

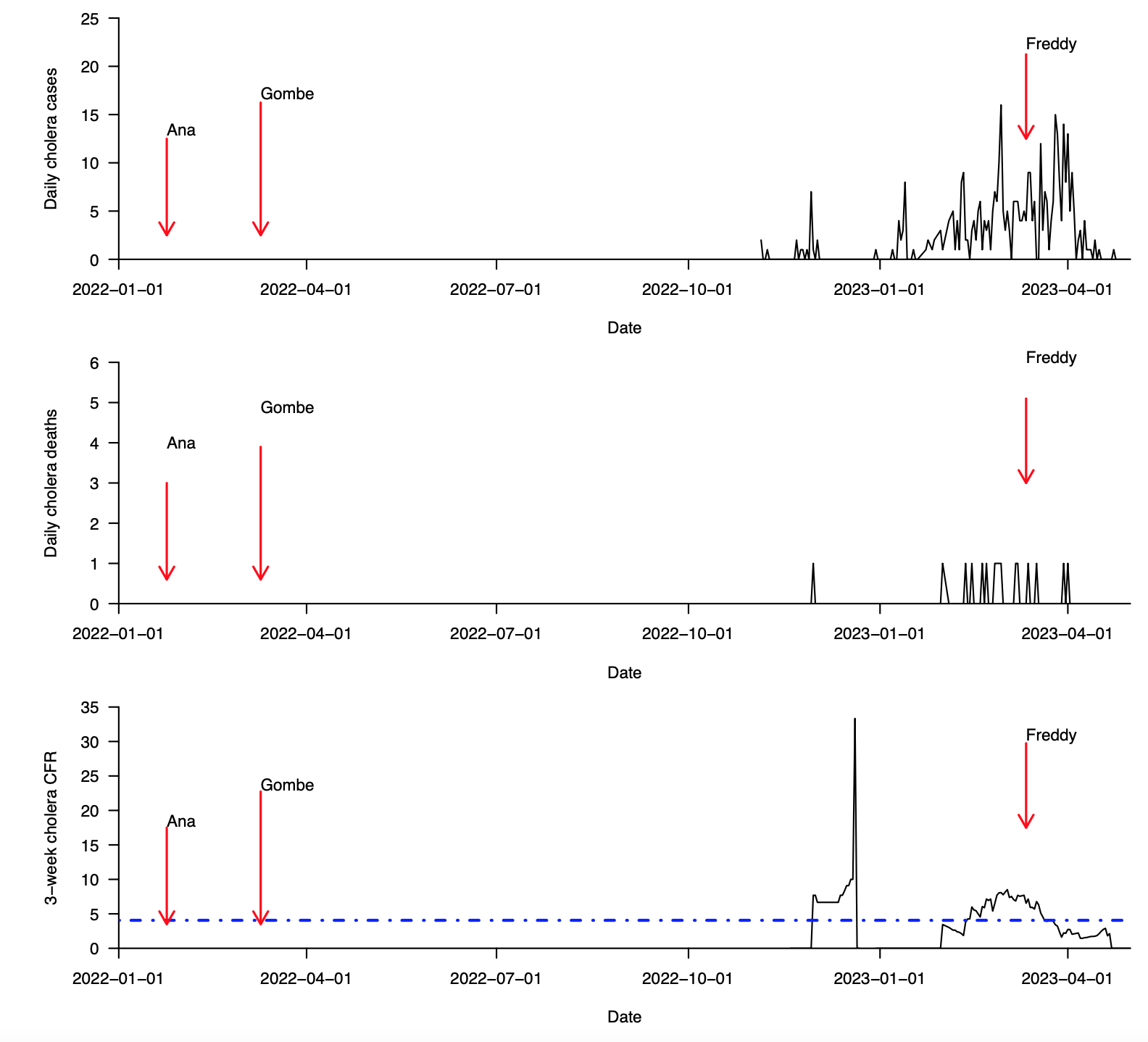

**Supplementary Fig. 2 (continued): Cases, deaths, and case fatality ratio during the 2022–2023 cholera outbreak in Mchinji district in Malawi (data from January, 2022 to May, 2023). (a)** Total daily cholera cases. **(b)** Total daily cholera deaths. **(c)** Overall cholera case fatality ratio (CFR) based on a 21-day sliding window. The 21-day sliding window was chosen to obtain stable estimates of the CFR, especially during weeks and months with few reported cholera cases. Data were obtained from the Public Health Institute of Malawi, Malawi Ministry of Health (MoH) data on May 20, 2023 [(https://cholera.health.gov.mw/surveillance](https://cholera.health.gov.mw/surveillance)).

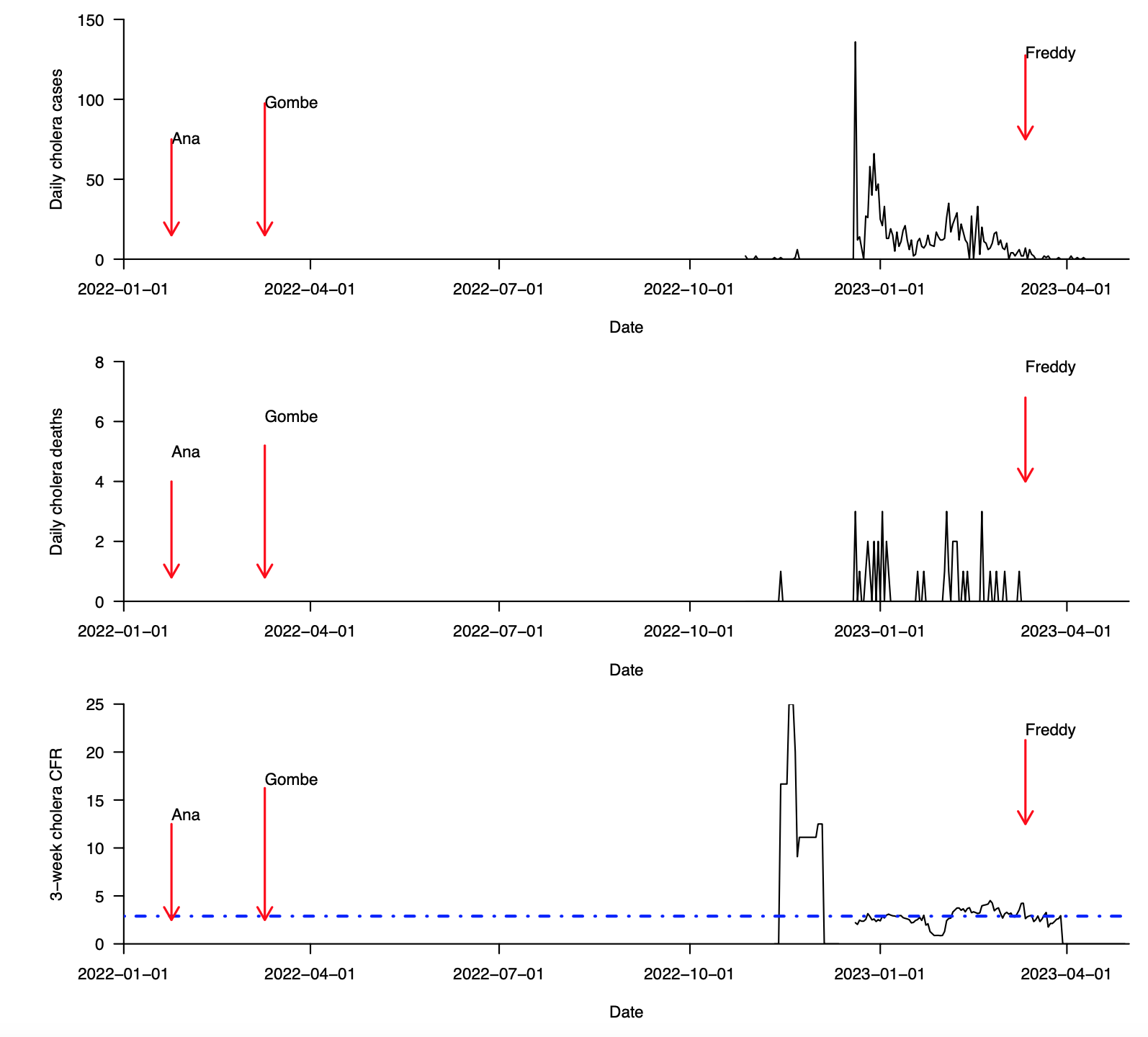

**Supplementary Fig. 2 (continued): Cases, deaths, and case fatality ratio during the 2022–2023 cholera outbreak in Dowa district in Malawi (data from January, 2022 to May, 2023). (a)** Total daily cholera cases. **(b)** Total daily cholera deaths. **(c)** Overall cholera case fatality ratio (CFR) based on a 21-day sliding window. The 21-day sliding window was chosen to obtain stable estimates of the CFR, especially during weeks and months with few reported cholera cases. Data were obtained from the Public Health Institute of Malawi, Malawi Ministry of Health (MoH) data on May 20, 2023 [(https://cholera.health.gov.mw/surveillance](https://cholera.health.gov.mw/surveillance)).

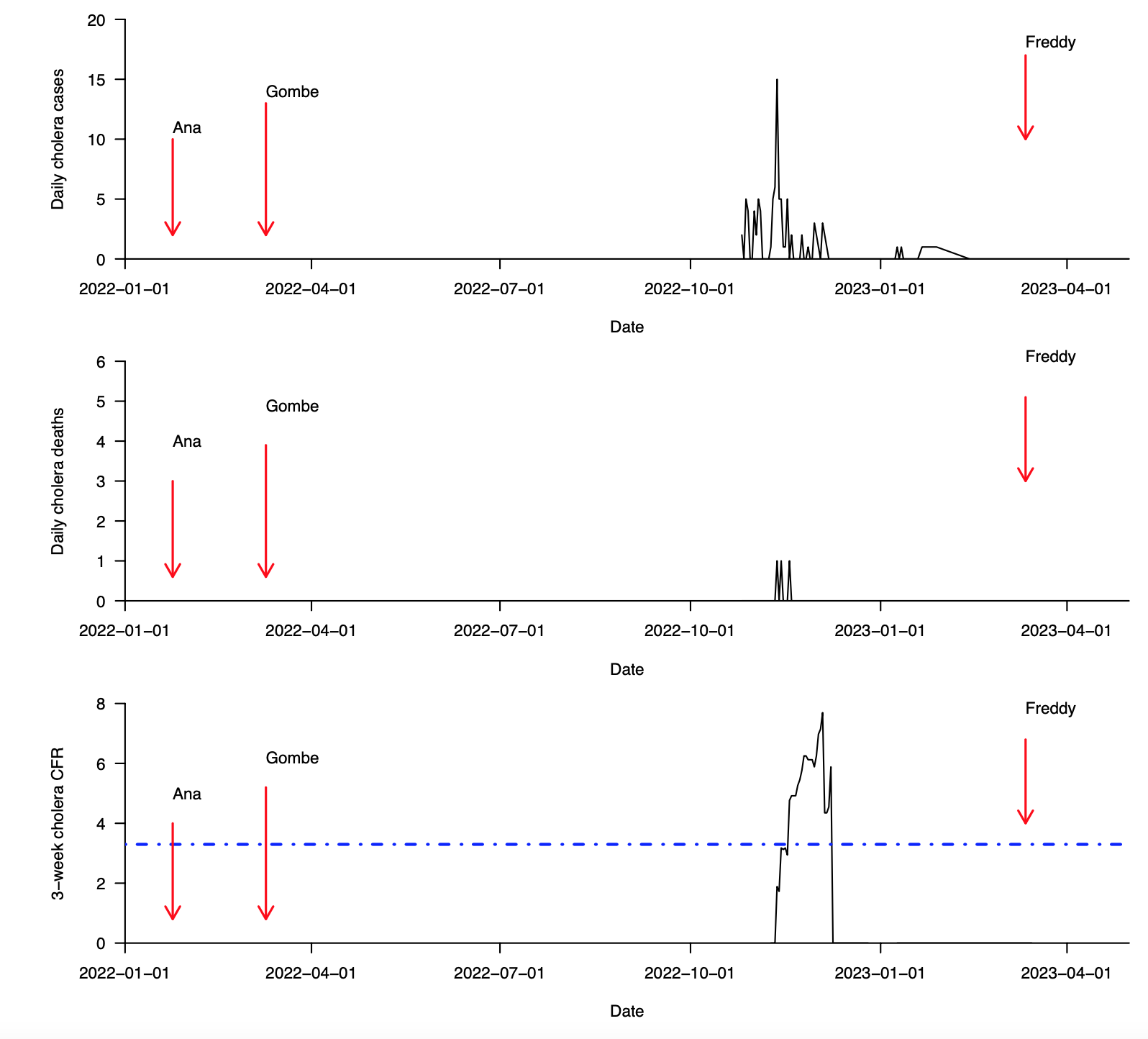

**Supplementary Fig. 2 (continued): Cases, deaths, and case fatality ratio during the 2022–2023 cholera outbreak in Chitipa district in Malawi (data from January, 2022 to May, 2023). (a)** Total daily cholera cases. **(b)** Total daily cholera deaths. **(c)** Overall cholera case fatality ratio (CFR) based on a 21-day sliding window. The 21-day sliding window was chosen to obtain stable estimates of the CFR, especially during weeks and months with few reported cholera cases. Data were obtained from the Public Health Institute of Malawi, Malawi Ministry of Health (MoH) data on May 20, 2023 [(https://cholera.health.gov.mw/surveillance](https://cholera.health.gov.mw/surveillance)).

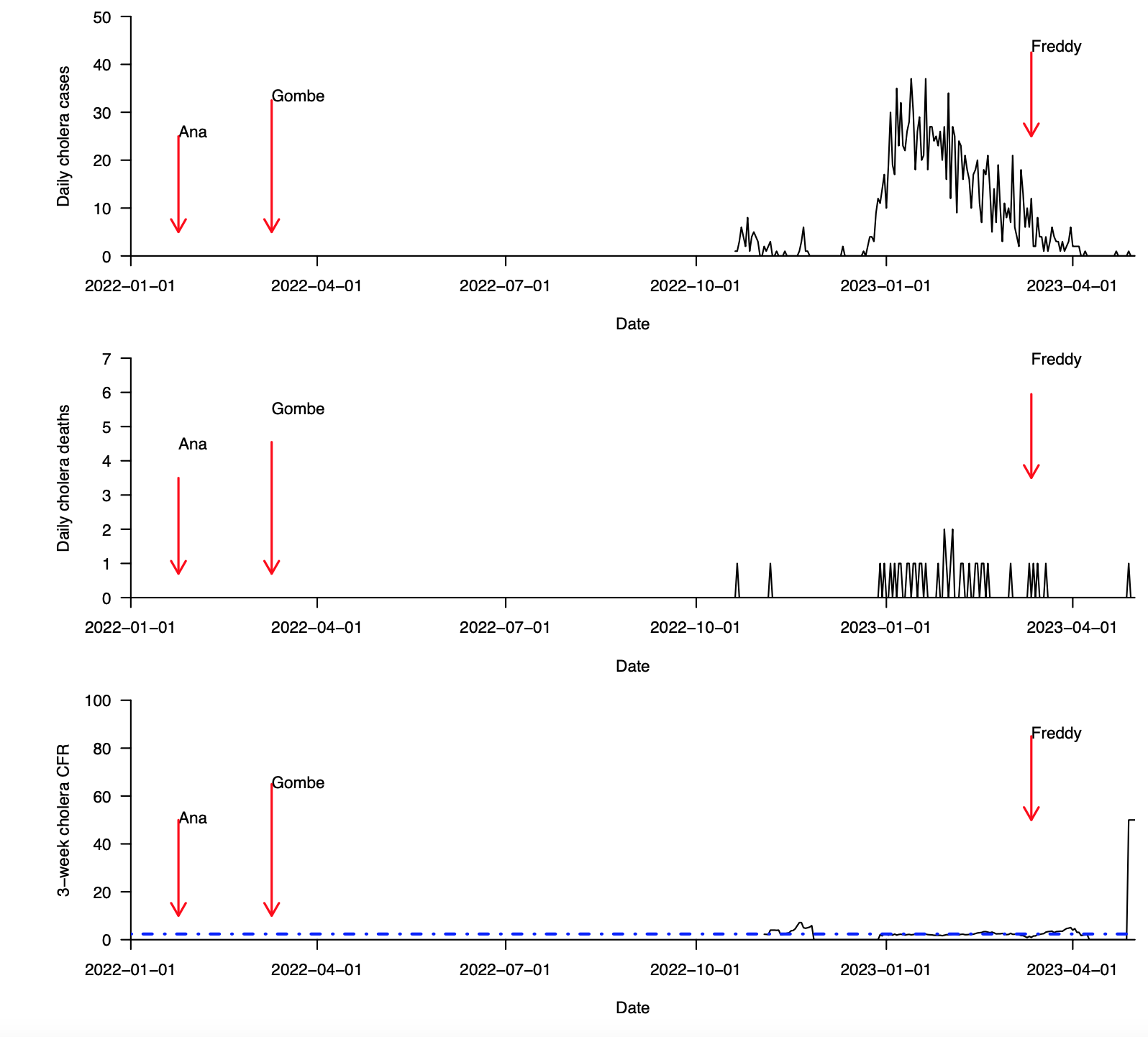

**Supplementary Fig. 2 (continued): Cases, deaths, and case fatality ratio during the 2022–2023 cholera outbreak in Thyolo district in Malawi (data from January, 2022 to May, 2023). (a)** Total daily cholera cases. **(b)** Total daily cholera deaths. **(c)** Overall cholera case fatality ratio (CFR) based on a 21-day sliding window. The 21-day sliding window was chosen to obtain stable estimates of the CFR, especially during weeks and months with few reported cholera cases. Data were obtained from the Public Health Institute of Malawi, Malawi Ministry of Health (MoH) data on May 20, 2023 [(https://cholera.health.gov.mw/surveillance](https://cholera.health.gov.mw/surveillance)).

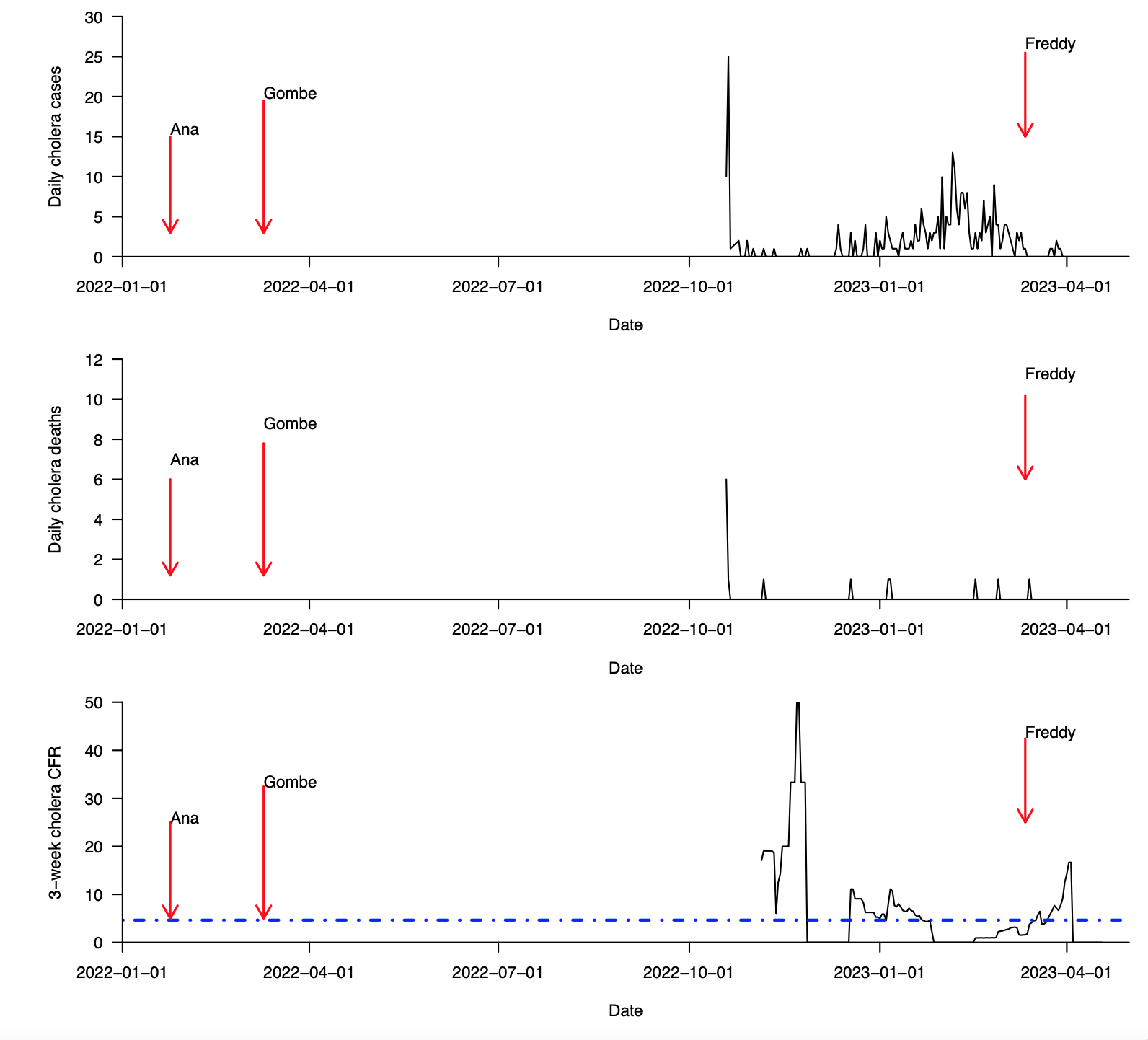

**Supplementary Fig. 2 (continued): Cases, deaths, and case fatality ratio during the 2022–2023 cholera outbreak in Phalombe district in Malawi (data from January, 2022 to May, 2023). (a)** Total daily cholera cases. **(b)** Total daily cholera deaths. **(c)** Overall cholera case fatality ratio (CFR) based on a 21-day sliding window. The 21-day sliding window was chosen to obtain stable estimates of the CFR, especially during weeks and months with few reported cholera cases. Data were obtained from the Public Health Institute of Malawi, Malawi Ministry of Health (MoH) data on May 20, 2023 [(https://cholera.health.gov.mw/surveillance](https://cholera.health.gov.mw/surveillance)).

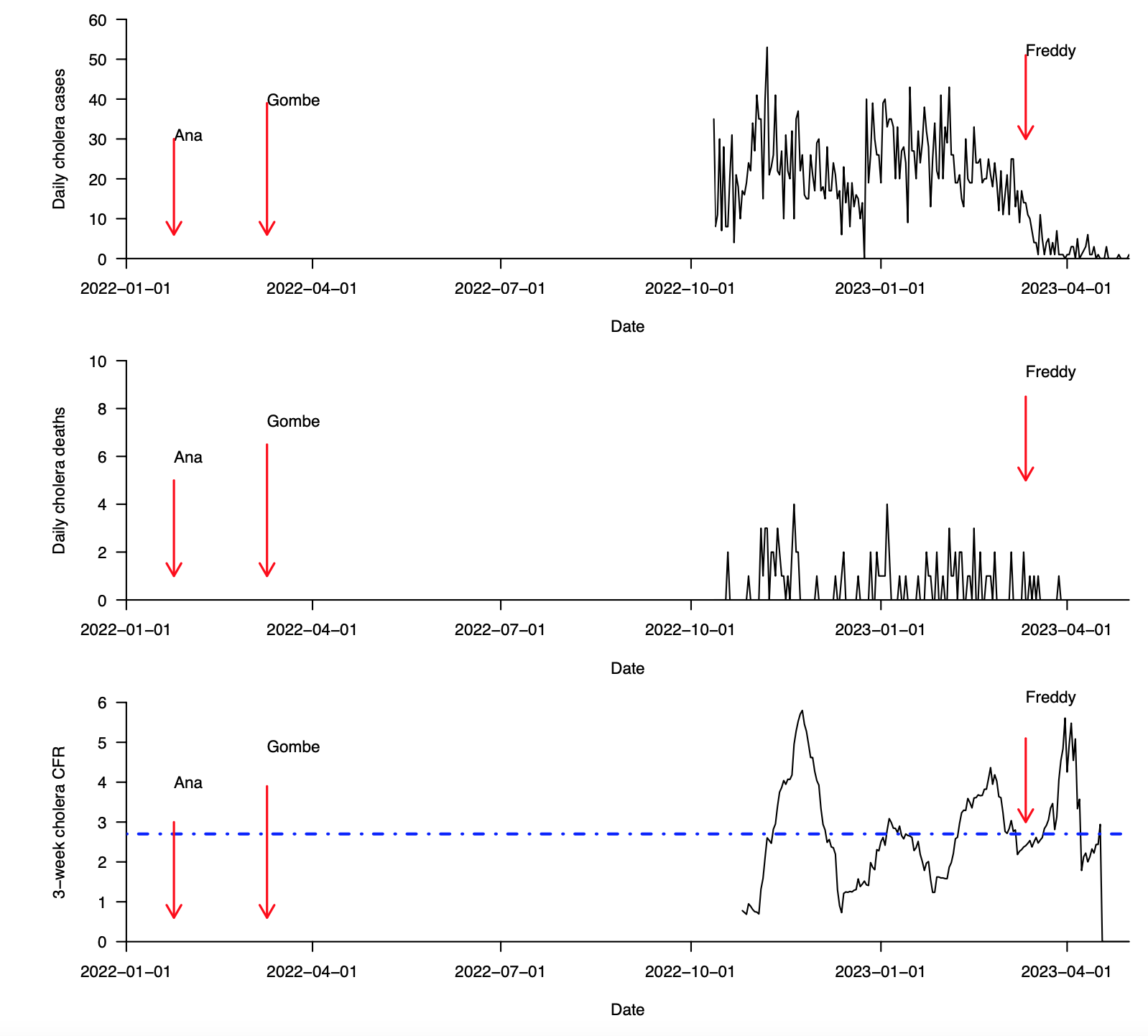

**Supplementary Fig. 2 (continued): Cases, deaths, and case fatality ratio during the 2022–2023 cholera outbreak in Salima district in Malawi (data from January, 2022 to May, 2023). (a)** Total daily cholera cases. **(b)** Total daily cholera deaths. **(c)** Overall cholera case fatality ratio (CFR) based on a 21-day sliding window. The 21-day sliding window was chosen to obtain stable estimates of the CFR, especially during weeks and months with few reported cholera cases. Data were obtained from the Public Health Institute of Malawi, Malawi Ministry of Health (MoH) data on May 20, 2023 [(https://cholera.health.gov.mw/surveillance](https://cholera.health.gov.mw/surveillance)).

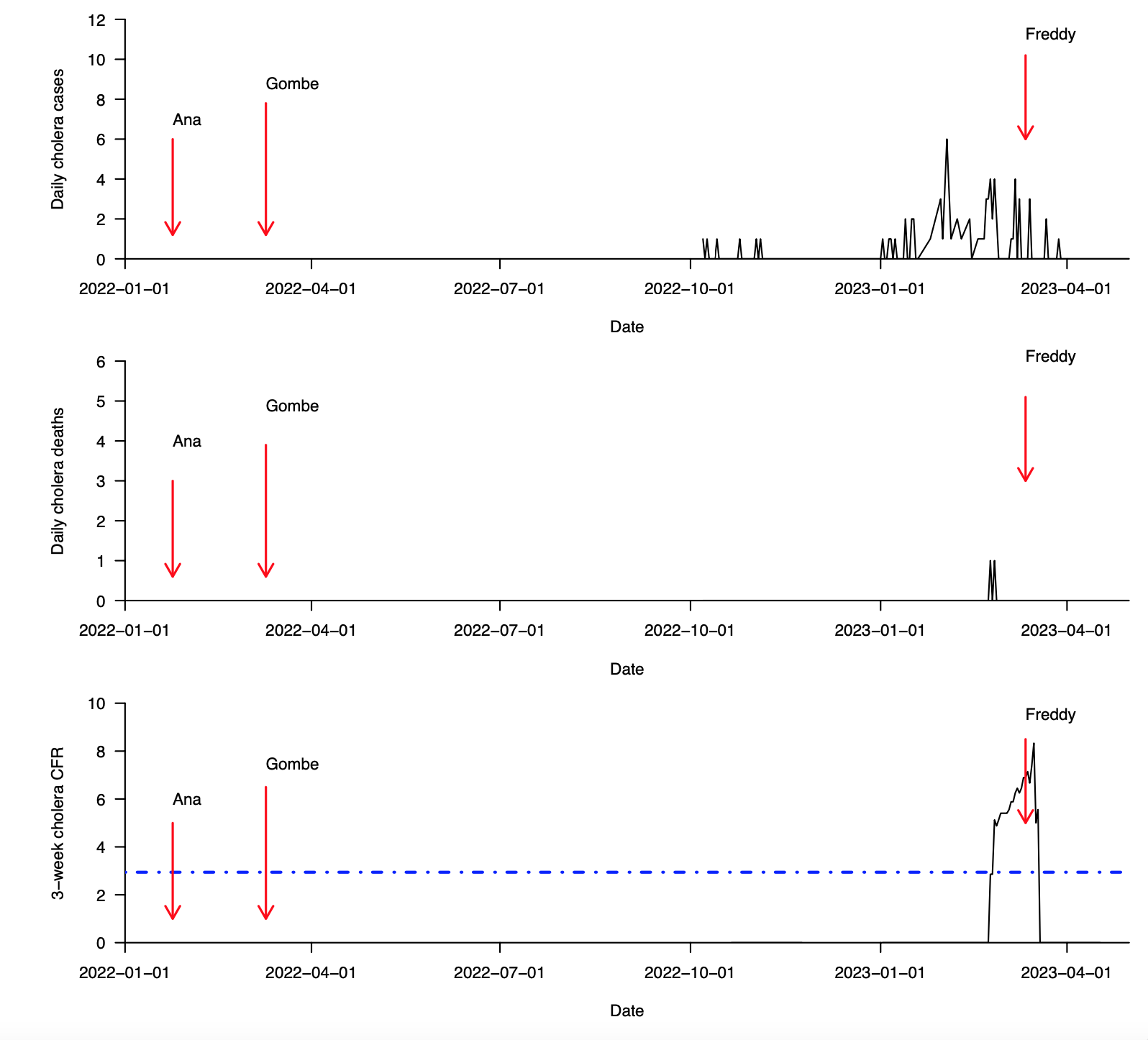

**Supplementary Fig. 2 (continued): Cases, deaths, and case fatality ratio during the 2022–2023 cholera outbreak in Ntchisi district in Malawi (data from January, 2022 to May, 2023). (a)** Total daily cholera cases. **(b)** Total daily cholera deaths. **(c)** Overall cholera case fatality ratio (CFR) based on a 21-day sliding window. The 21-day sliding window was chosen to obtain stable estimates of the CFR, especially during weeks and months with few reported cholera cases. Data were obtained from the Public Health Institute of Malawi, Malawi Ministry of Health (MoH) data on May 20, 2023 [(https://cholera.health.gov.mw/surveillance](https://cholera.health.gov.mw/surveillance)).

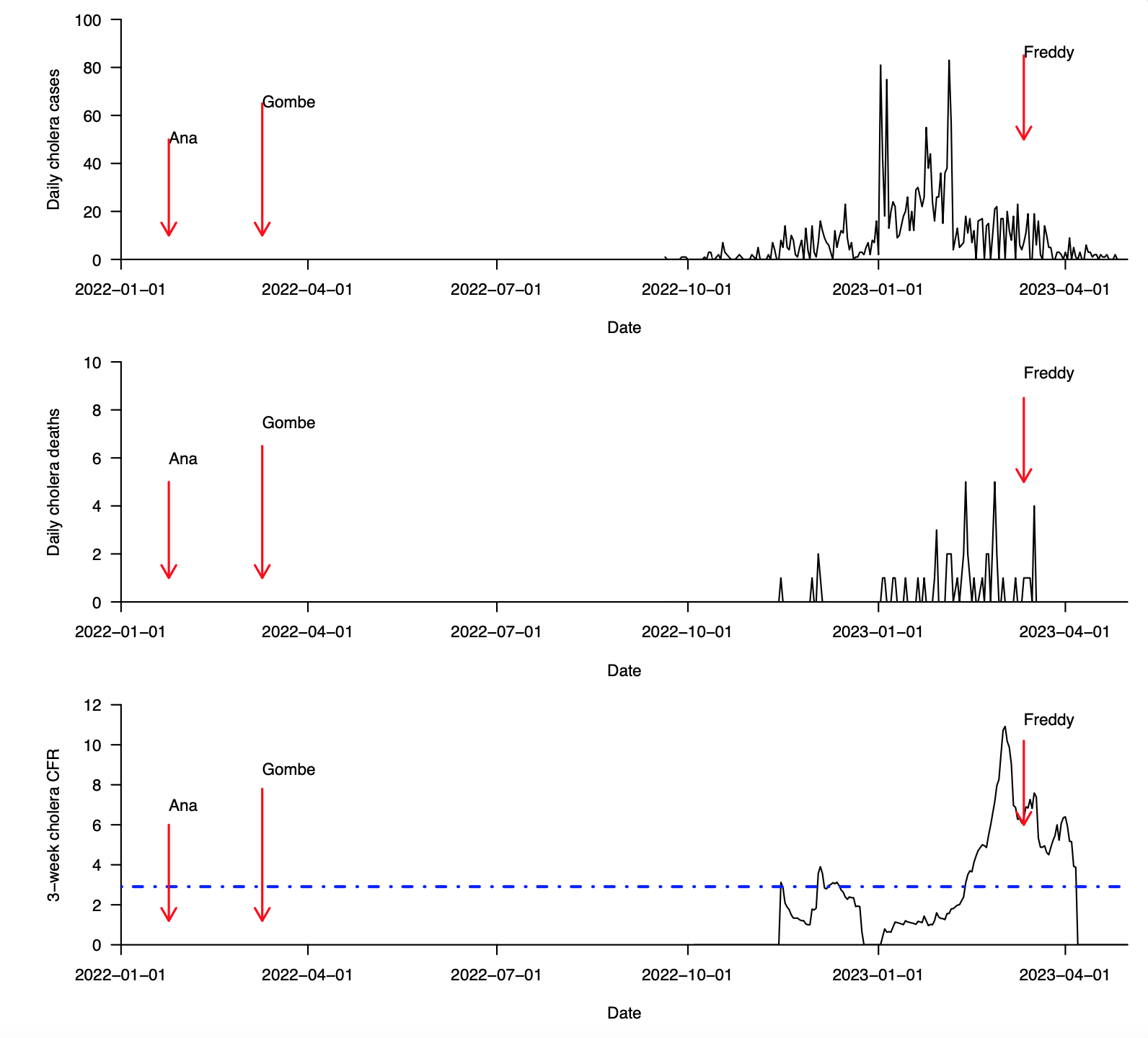

**Supplementary Fig. 2 (continued): Cases, deaths, and case fatality ratio during the 2022–2023 cholera outbreak in Ntcheu district in Malawi (data from January, 2022 to May, 2023). (a)** Total daily cholera cases. **(b)** Total daily cholera deaths. **(c)** Overall cholera case fatality ratio (CFR) based on a 21-day sliding window. The 21-day sliding window was chosen to obtain stable estimates of the CFR, especially during weeks and months with few reported cholera cases. Data were obtained from the Public Health Institute of Malawi, Malawi Ministry of Health (MoH) data on May 20, 2023 [(https://cholera.health.gov.mw/surveillance](https://cholera.health.gov.mw/surveillance)).

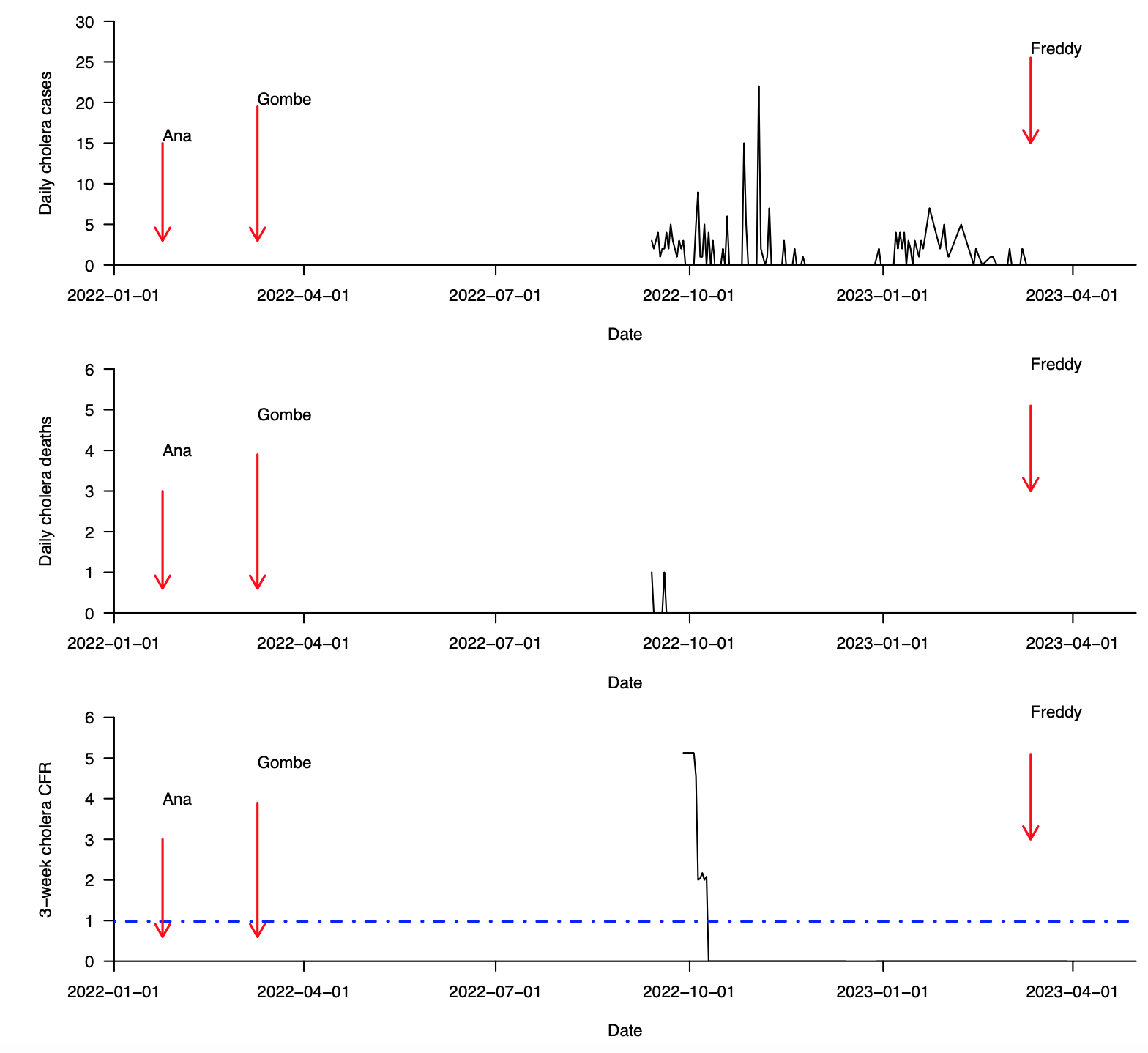

**Supplementary Fig. 2 (continued): Cases, deaths, and case fatality ratio during the 2022–2023 cholera outbreak in Likoma district in Malawi (data from January, 2022 to May, 2023). (a)** Total daily cholera cases. **(b)** Total daily cholera deaths. **(c)** Overall cholera case fatality ratio (CFR) based on a 21-day sliding window. The 21-day sliding window was chosen to obtain stable estimates of the CFR, especially during weeks and months with few reported cholera cases. Data were obtained from the Public Health Institute of Malawi, Malawi Ministry of Health (MoH) data on May 20, 2023 [(https://cholera.health.gov.mw/surveillance](https://cholera.health.gov.mw/surveillance)).

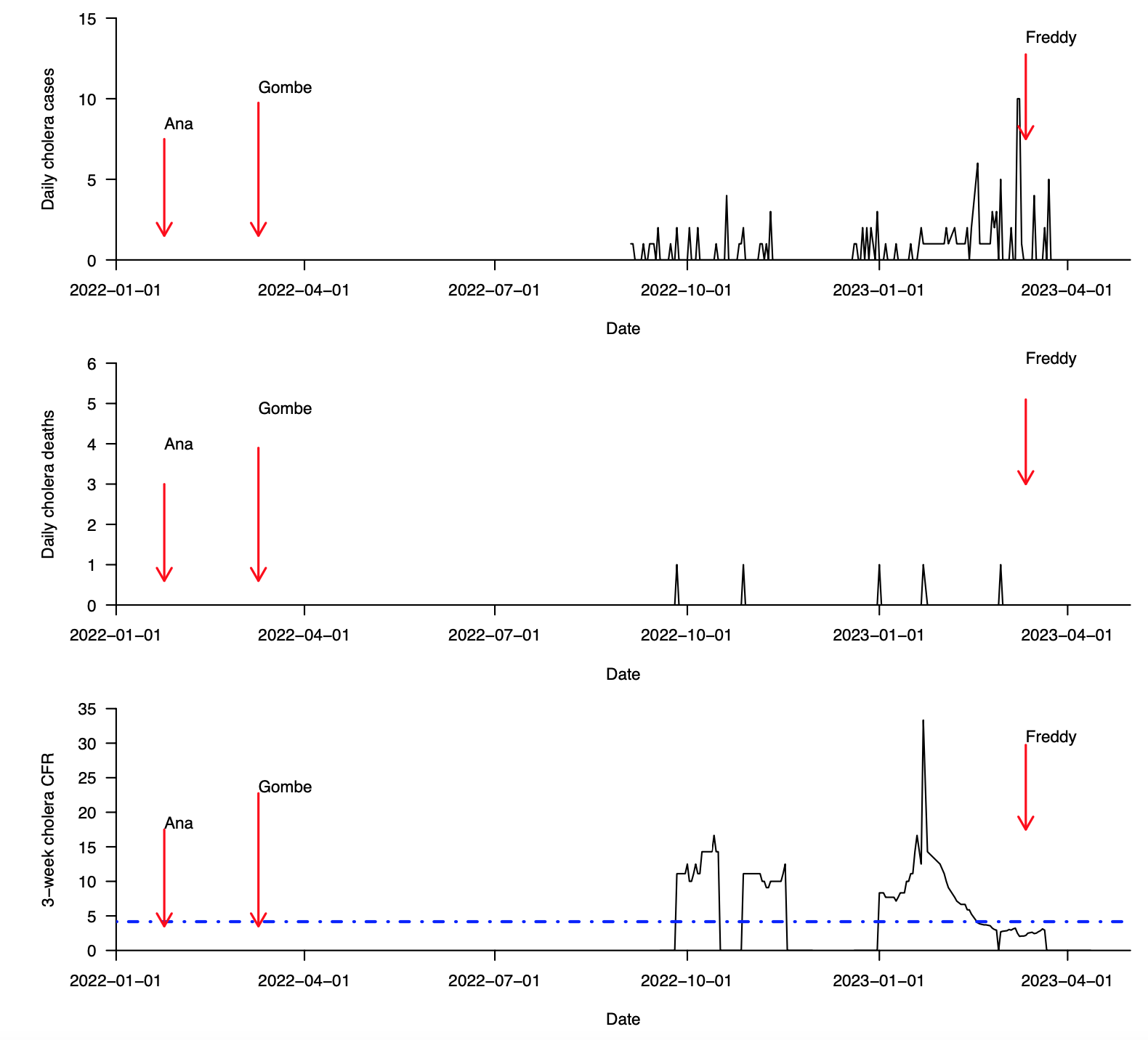

**Supplementary Fig. 2 (continued): Cases, deaths, and case fatality ratio during the 2022–2023 cholera outbreak in Mwanza district in Malawi (data from January, 2022 to May, 2023). (a)** Total daily cholera cases. **(b)** Total daily cholera deaths. **(c)** Overall cholera case fatality ratio (CFR) based on a 21-day sliding window. The 21-day sliding window was chosen to obtain stable estimates of the CFR, especially during weeks and months with few reported cholera cases. Data were obtained from the Public Health Institute of Malawi, Malawi Ministry of Health (MoH) data on May 20, 2023 [(https://cholera.health.gov.mw/surveillance](https://cholera.health.gov.mw/surveillance)).

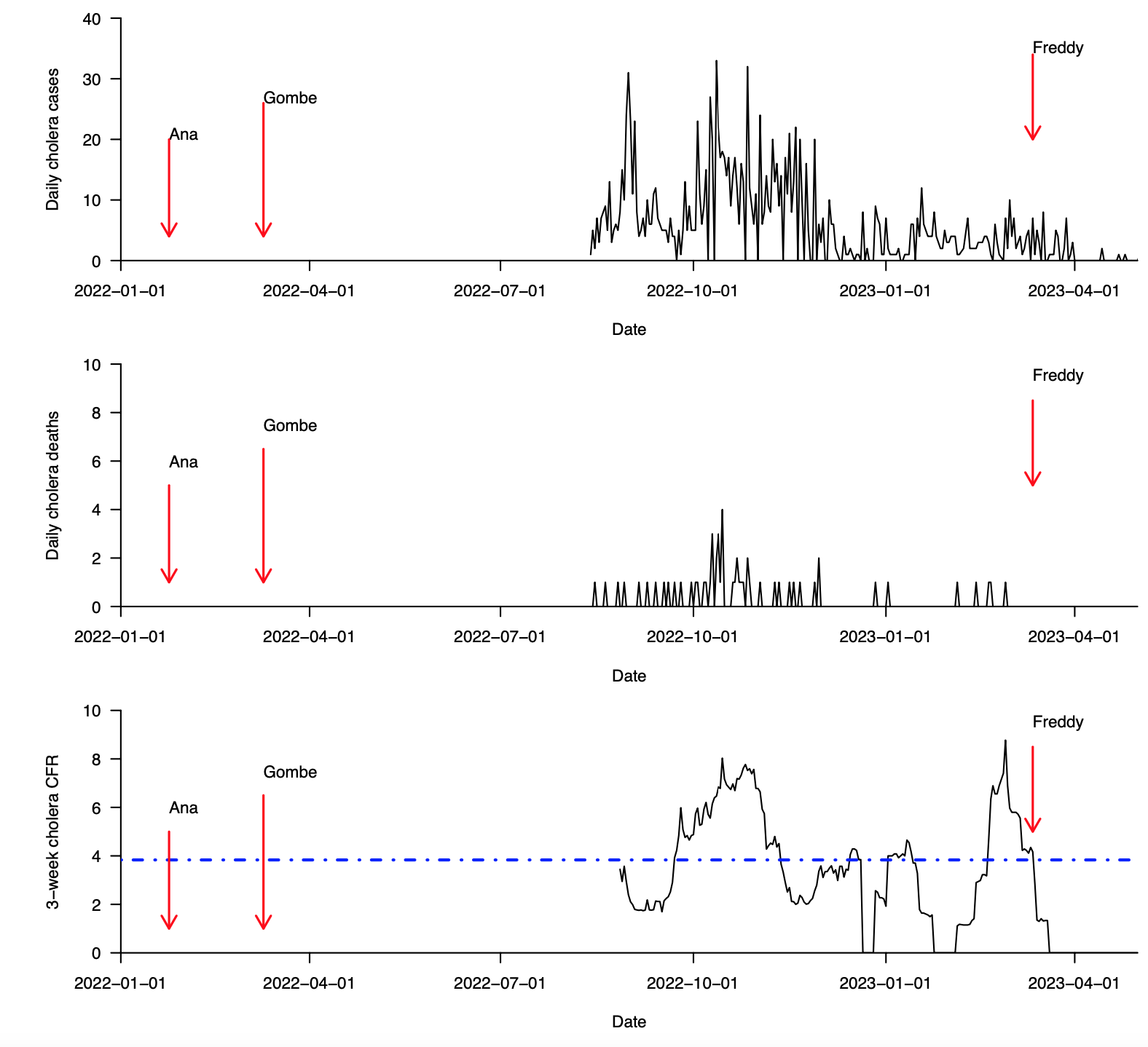

**Supplementary Fig. 2 (continued): Cases, deaths, and case fatality ratio during the 2022–2023 cholera outbreak in Kasungu district in Malawi (data from January, 2022 to May, 2023). (a)** Total daily cholera cases. **(b)** Total daily cholera deaths. **(c)** Overall cholera case fatality ratio (CFR) based on a 21-day sliding window. The 21-day sliding window was chosen to obtain stable estimates of the CFR, especially during weeks and months with few reported cholera cases. Data were obtained from the Public Health Institute of Malawi, Malawi Ministry of Health (MoH) data on May 20, 2023 [(https://cholera.health.gov.mw/surveillance](https://cholera.health.gov.mw/surveillance)).

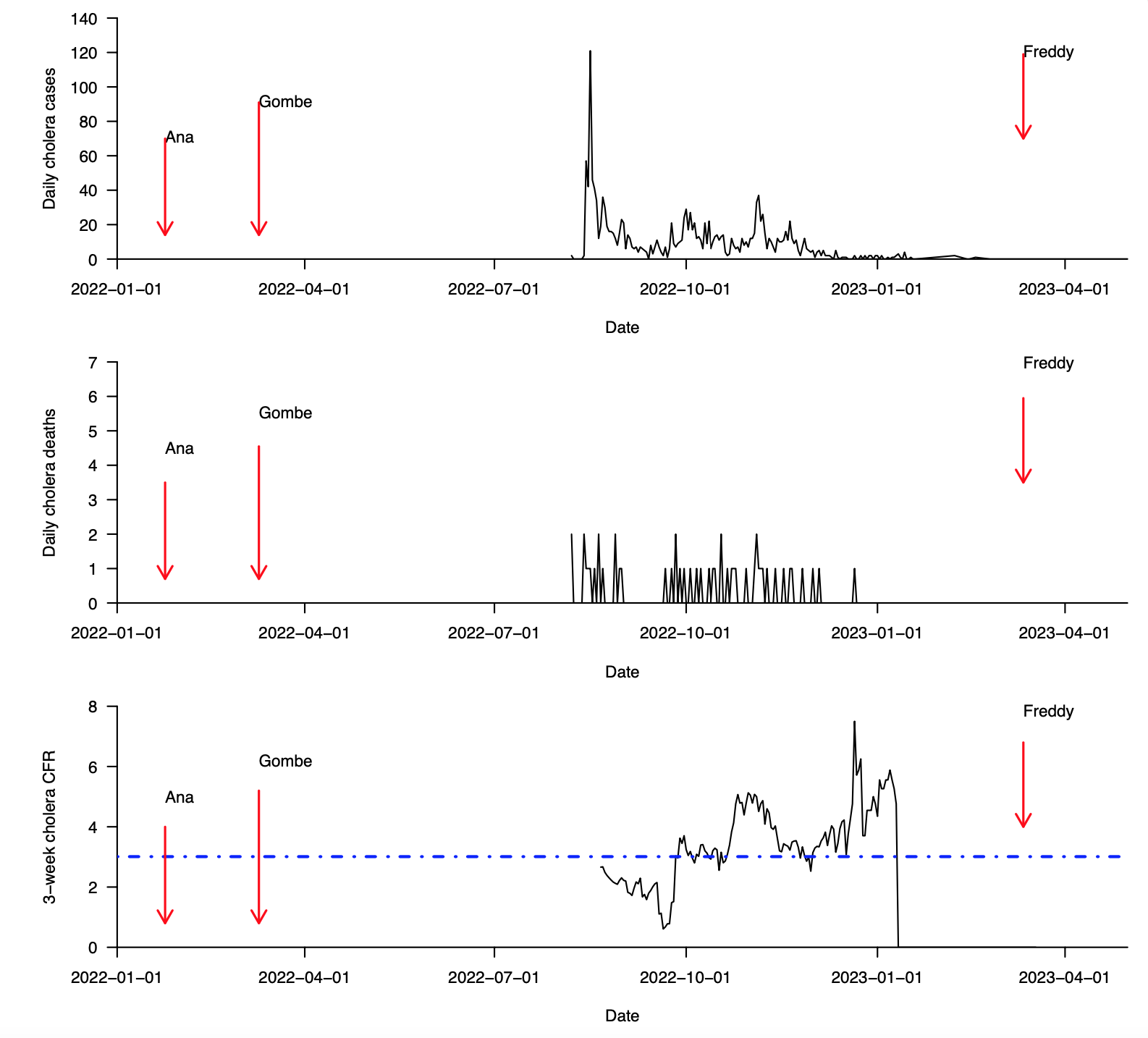

**Supplementary Fig. 2 (continued): Cases, deaths, and case fatality ratio during the 2022–2023 cholera outbreak in Karonga district in Malawi (data from January, 2022 to May, 2023). (a)** Total daily cholera cases. **(b)** Total daily cholera deaths. **(c)** Overall cholera case fatality ratio (CFR) based on a 21-day sliding window. The 21-day sliding window was chosen to obtain stable estimates of the CFR, especially during weeks and months with few reported cholera cases. Data were obtained from the Public Health Institute of Malawi, Malawi Ministry of Health (MoH) data on May 20, 2023 [(https://cholera.health.gov.mw/surveillance](https://cholera.health.gov.mw/surveillance)).

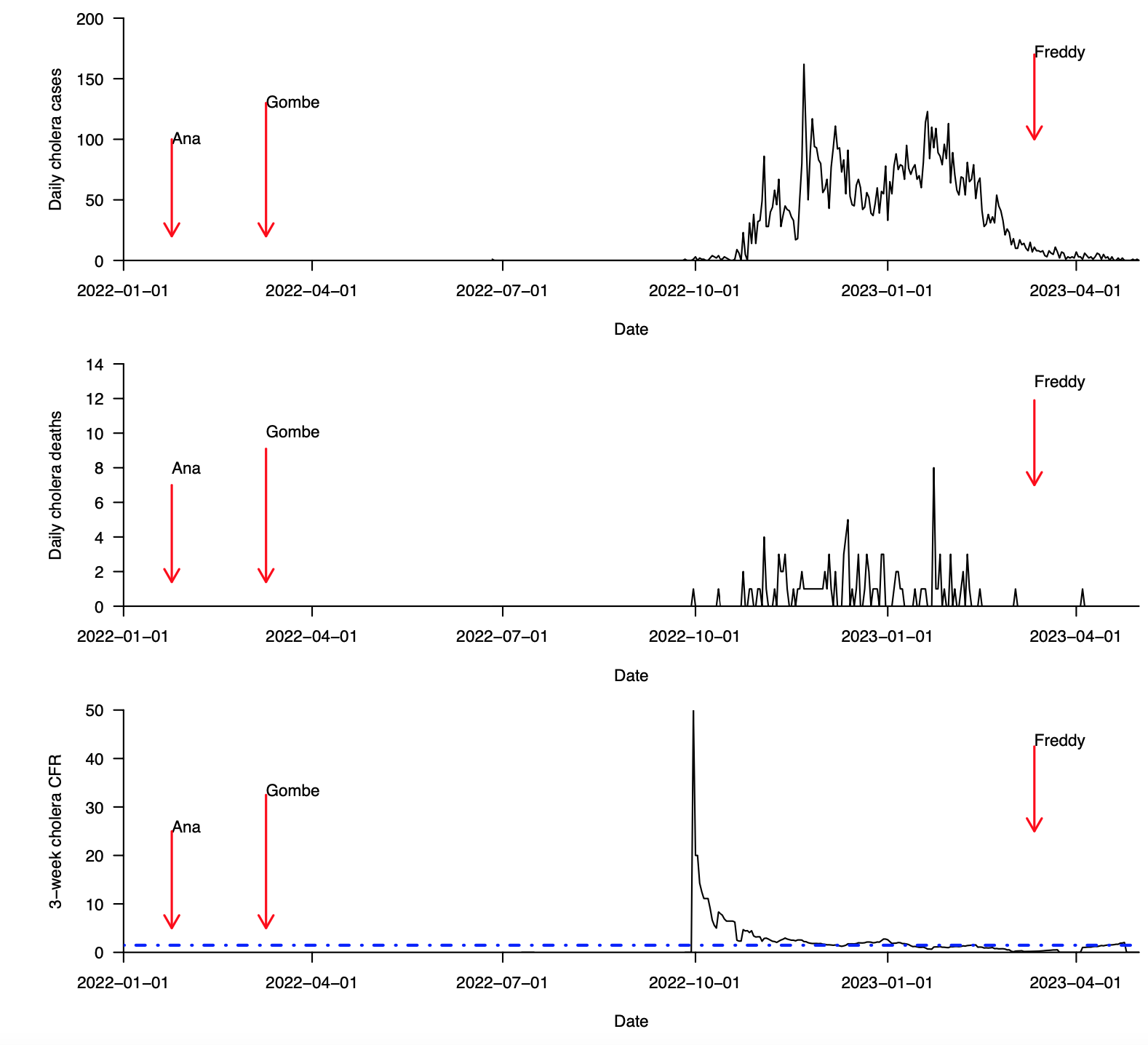

**Supplementary Fig. 2 (continued): Cases, deaths, and case fatality ratio during the 2022–2023 cholera outbreak in Zomba district in Malawi (data from January, 2022 to May, 2023). (a)** Total daily cholera cases. **(b)** Total daily cholera deaths. **(c)** Overall cholera case fatality ratio (CFR) based on a 21-day sliding window. The 21-day sliding window was chosen to obtain stable estimates of the CFR, especially during weeks and months with few reported cholera cases. Data were obtained from the Public Health Institute of Malawi, Malawi Ministry of Health (MoH) data on May 20, 2023 [(https://cholera.health.gov.mw/surveillance](https://cholera.health.gov.mw/surveillance)).

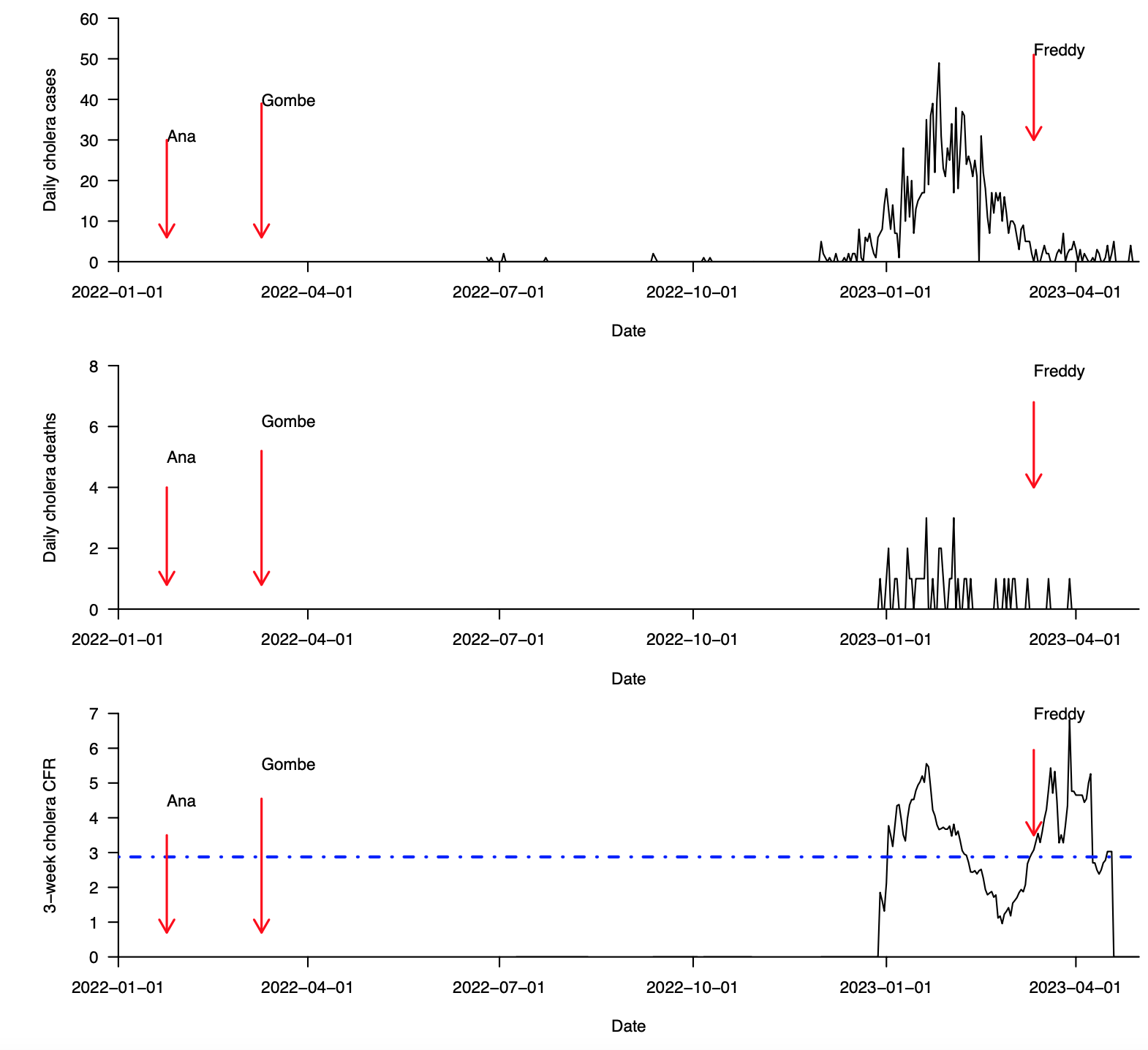

**Supplementary Fig. 2 (continued): Cases, deaths, and case fatality ratio during the 2022–2023 cholera outbreak in Rumphi district in Malawi (data from January, 2022 to May, 2023). (a)** Total daily cholera cases. **(b)** Total daily cholera deaths. **(c)** Overall cholera case fatality ratio (CFR) based on a 21-day sliding window. The 21-day sliding window was chosen to obtain stable estimates of the CFR, especially during weeks and months with few reported cholera cases. Data were obtained from the Public Health Institute of Malawi, Malawi Ministry of Health (MoH) data on May 20, 2023 [(https://cholera.health.gov.mw/surveillance](https://cholera.health.gov.mw/surveillance)).

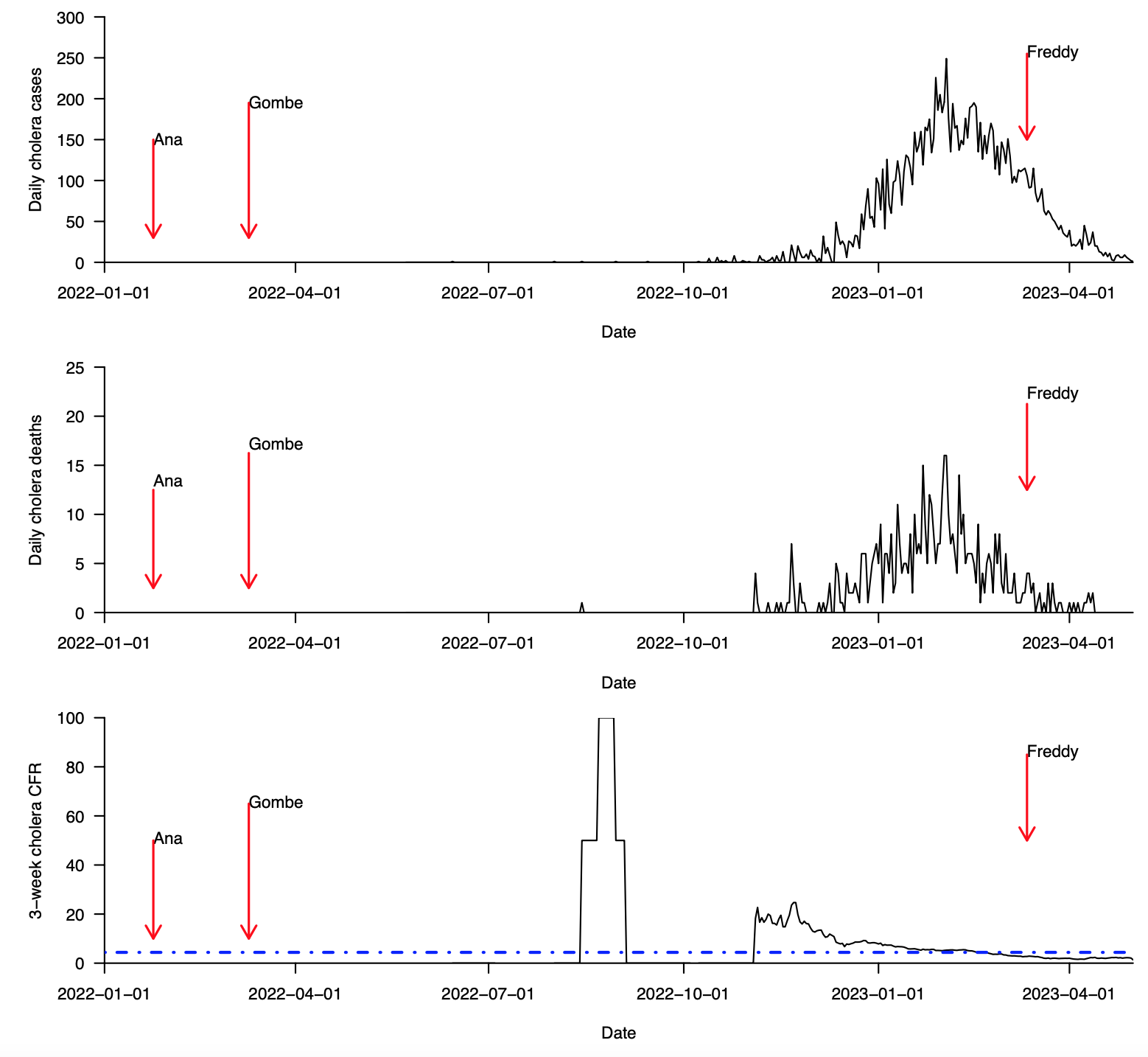

**Supplementary Fig. 2 (continued): Cases, deaths, and case fatality ratio during the 2022–2023 cholera outbreak in Mzimba district in Malawi (data from January, 2022 to May, 2023). (a)** Total daily cholera cases. **(b)** Total daily cholera deaths. **(c)** Overall cholera case fatality ratio (CFR) based on a 21-day sliding window. The 21-day sliding window was chosen to obtain stable estimates of the CFR, especially during weeks and months with few reported cholera cases. Data were obtained from the Public Health Institute of Malawi, Malawi Ministry of Health (MoH) data on May 20, 2023 [(https://cholera.health.gov.mw/surveillance](https://cholera.health.gov.mw/surveillance)).

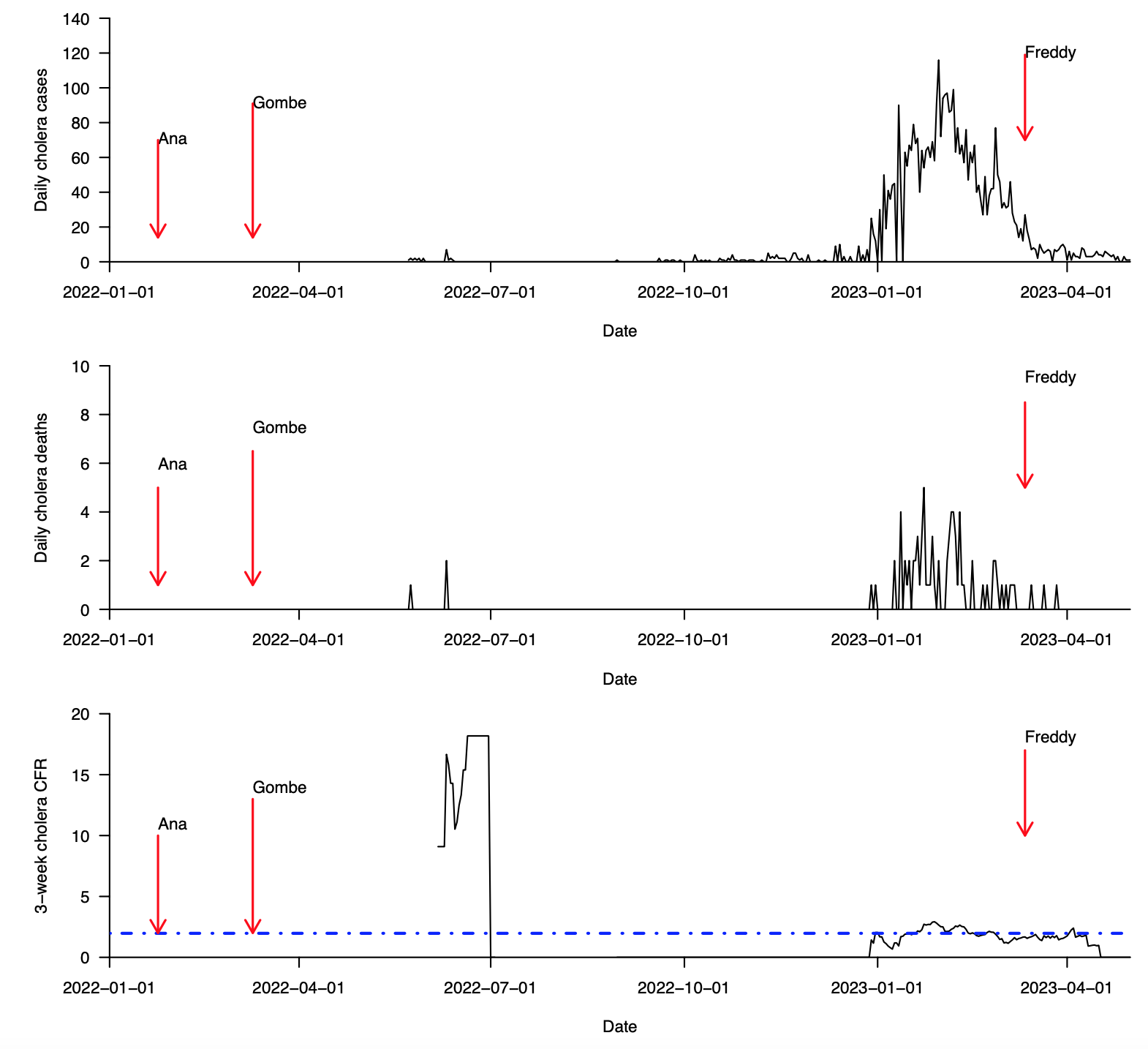

**Supplementary Fig. 2 (continued): Cases, deaths, and case fatality ratio during the 2022–2023 cholera outbreak in Nkhotakota district in Malawi (data from January, 2022 to May, 2023). (a)** Total daily cholera cases. **(b)** Total daily cholera deaths. **(c)** Overall cholera case fatality ratio (CFR) based on a 21-day sliding window. The 21-day sliding window was chosen to obtain stable estimates of the CFR, especially during weeks and months with few reported cholera cases. Data were obtained from the Public Health Institute of Malawi, Malawi Ministry of Health (MoH) data on May 20, 2023 [(https://cholera.health.gov.mw/surveillance](https://cholera.health.gov.mw/surveillance)).

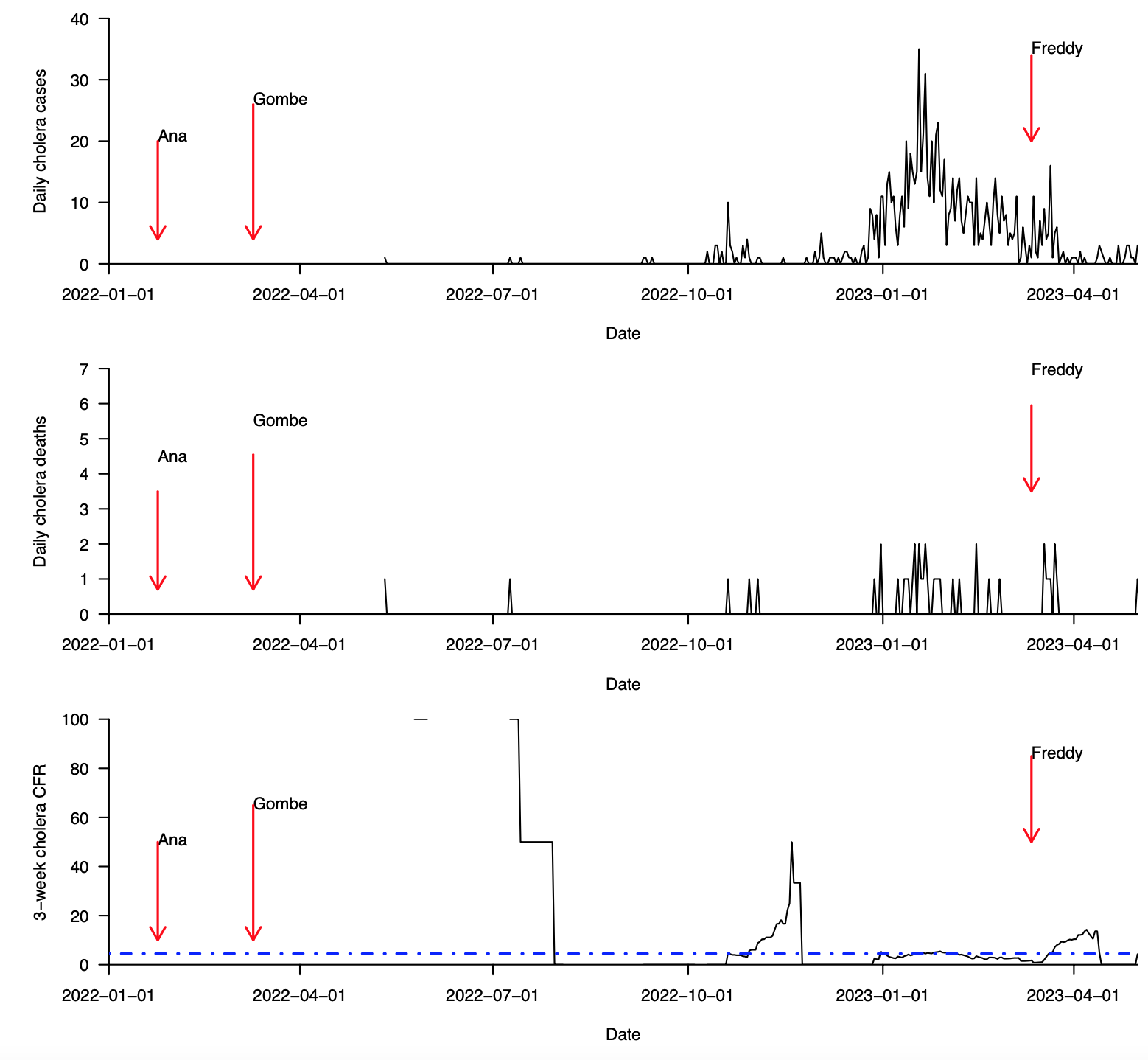

**Supplementary Fig. 2 (continued): Cases, deaths, and case fatality ratio during the 2022–2023 cholera outbreak in Nkhatabay district in Malawi (data from January, 2022 to May, 2023). (a)** Total daily cholera cases. **(b)** Total daily cholera deaths. **(c)** Overall cholera case fatality ratio (CFR) based on a 21-day sliding window. The 21-day sliding window was chosen to obtain stable estimates of the CFR, especially during weeks and months with few reported cholera cases. Data were obtained from the Public Health Institute of Malawi, Malawi Ministry of Health (MoH) data on May 20, 2023 [(https://cholera.health.gov.mw/surveillance](https://cholera.health.gov.mw/surveillance)).

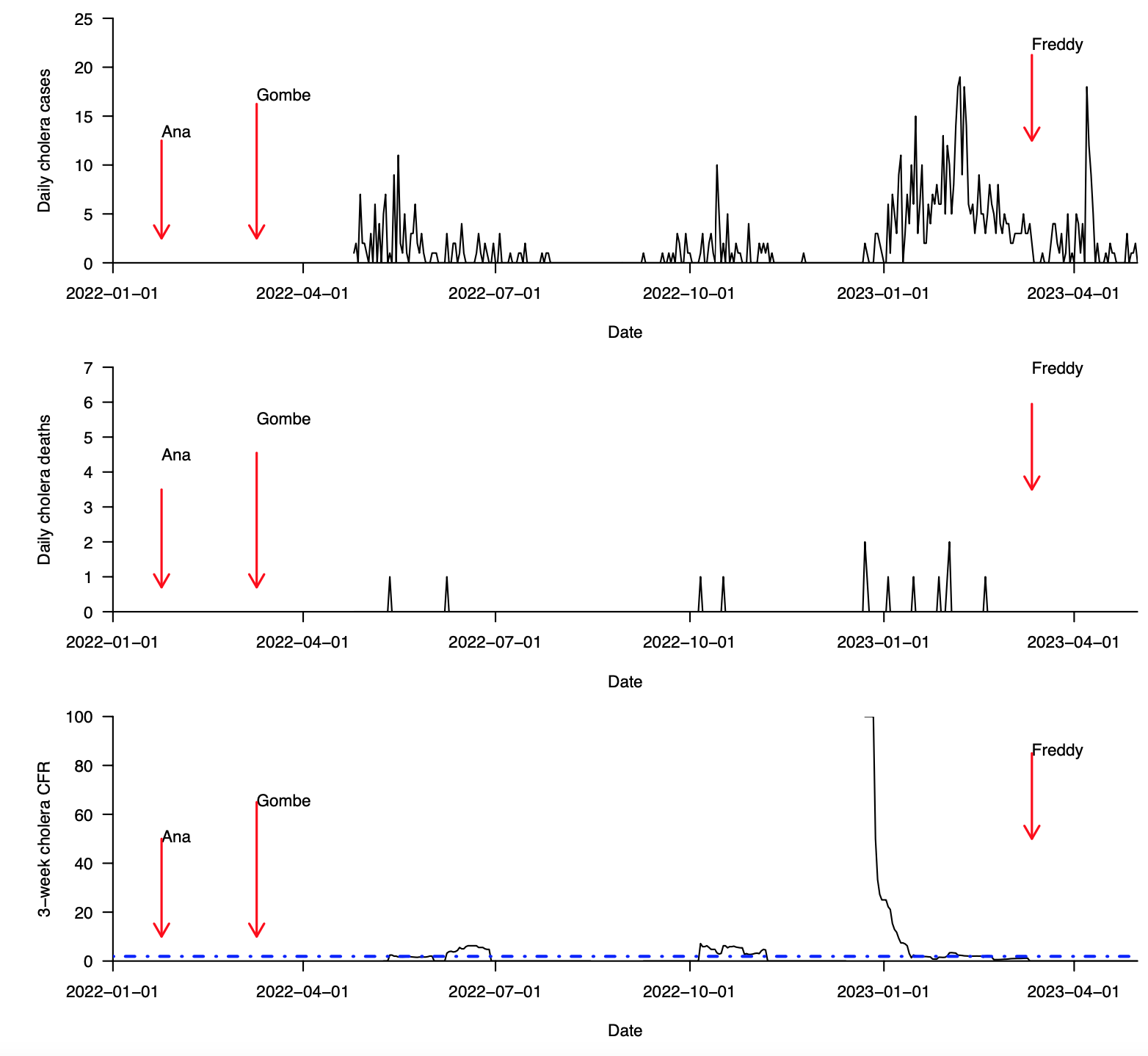

**Supplementary Fig. 2 (continued): Cases, deaths, and case fatality ratio during the 2022–2023 cholera outbreak in Mangochi district in Malawi (data from January, 2022 to May, 2023). (a)** Total daily cholera cases. **(b)** Total daily cholera deaths. **(c)** Overall cholera case fatality ratio (CFR) based on a 21-day sliding window. The 21-day sliding window was chosen to obtain stable estimates of the CFR, especially during weeks and months with few reported cholera cases. Data were obtained from the Public Health Institute of Malawi, Malawi Ministry of Health (MoH) data on May 20, 2023 [(https://cholera.health.gov.mw/surveillance](https://cholera.health.gov.mw/surveillance)).

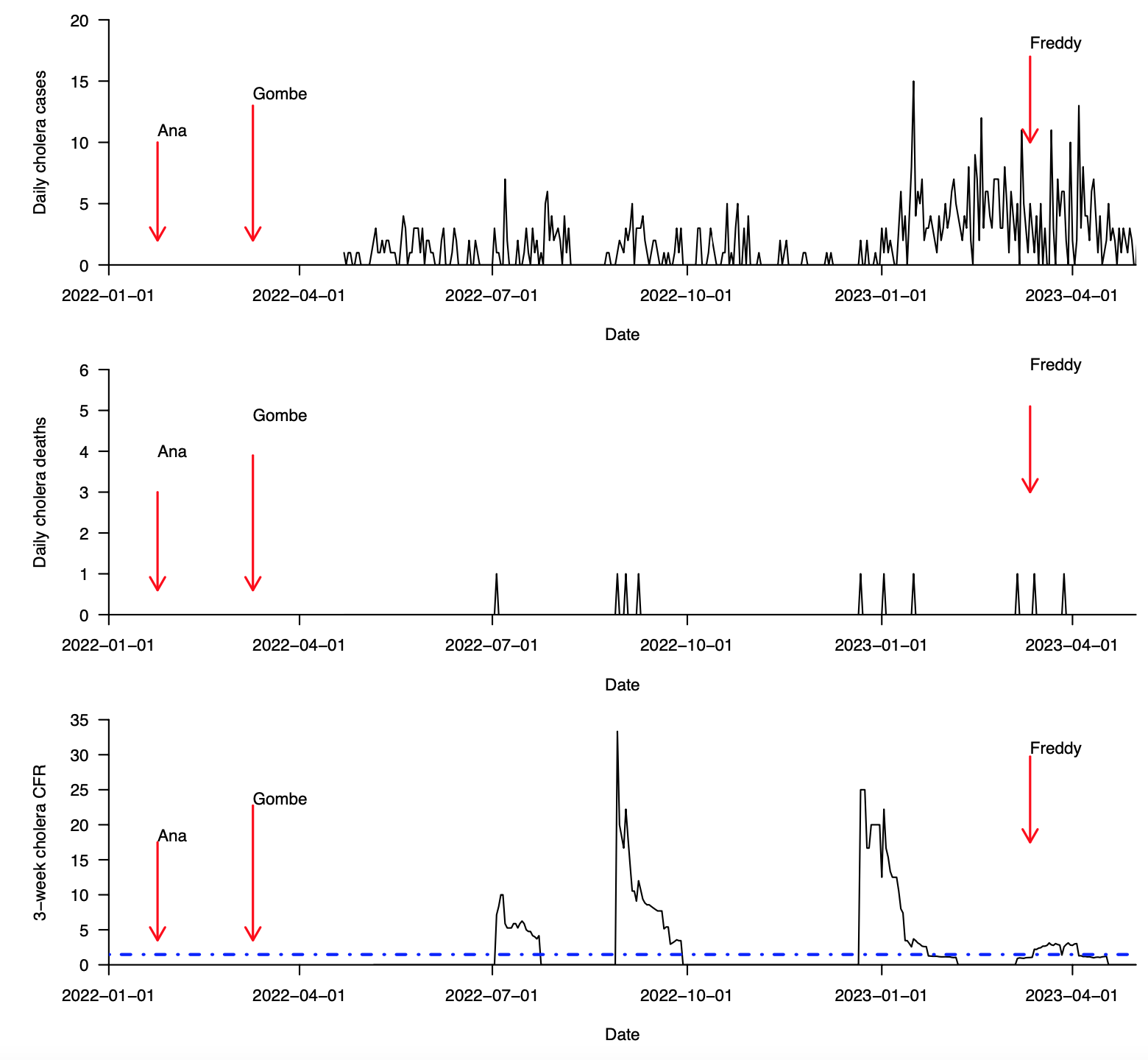

**Supplementary Fig. 2 (continued): Cases, deaths, and case fatality ratio during the 2022–2023 cholera outbreak in Chiradzulu district in Malawi (data from January, 2022 to May, 2023). (a)** Total daily cholera cases. **(b)** Total daily cholera deaths. **(c)** Overall cholera case fatality ratio (CFR) based on a 21-day sliding window. The 21-day sliding window was chosen to obtain stable estimates of the CFR, especially during weeks and months with few reported cholera cases. Data were obtained from the Public Health Institute of Malawi, Malawi Ministry of Health (MoH) data on May 20, 2023 [(https://cholera.health.gov.mw/surveillance](https://cholera.health.gov.mw/surveillance)).

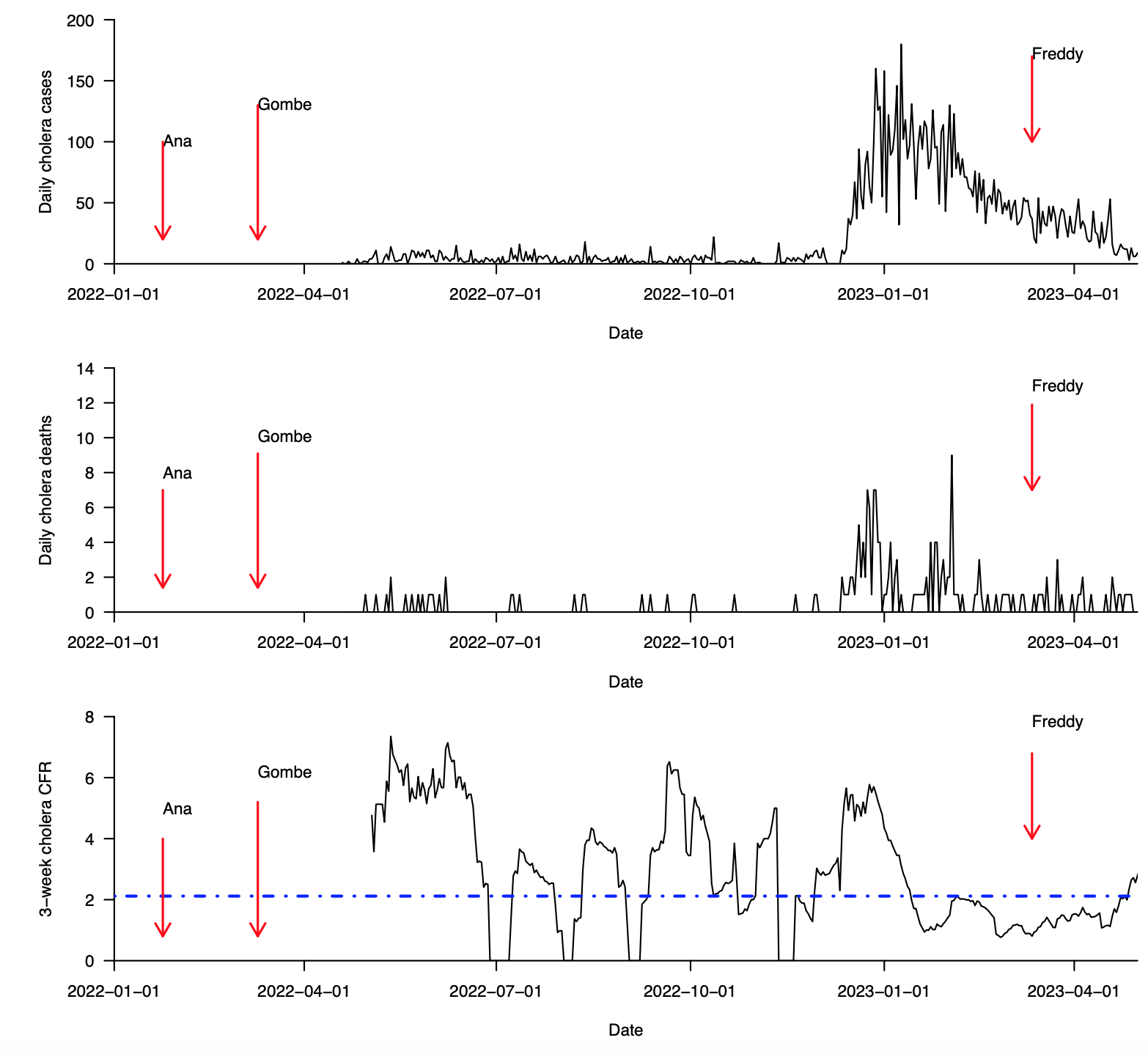

**Supplementary Fig. 2 (continued): Cases, deaths, and case fatality ratio during the 2022–2023 cholera outbreak in Lilongwe district in Malawi (data from January, 2022 to May, 2023). (a)** Total daily cholera cases. **(b)** Total daily cholera deaths. **(c)** Overall cholera case fatality ratio (CFR) based on a 21-day sliding window. The 21-day sliding window was chosen to obtain stable estimates of the CFR, especially during weeks and months with few reported cholera cases. Data were obtained from the Public Health Institute of Malawi, Malawi Ministry of Health (MoH) data on May 20, 2023 [(https://cholera.health.gov.mw/surveillance](https://cholera.health.gov.mw/surveillance)).

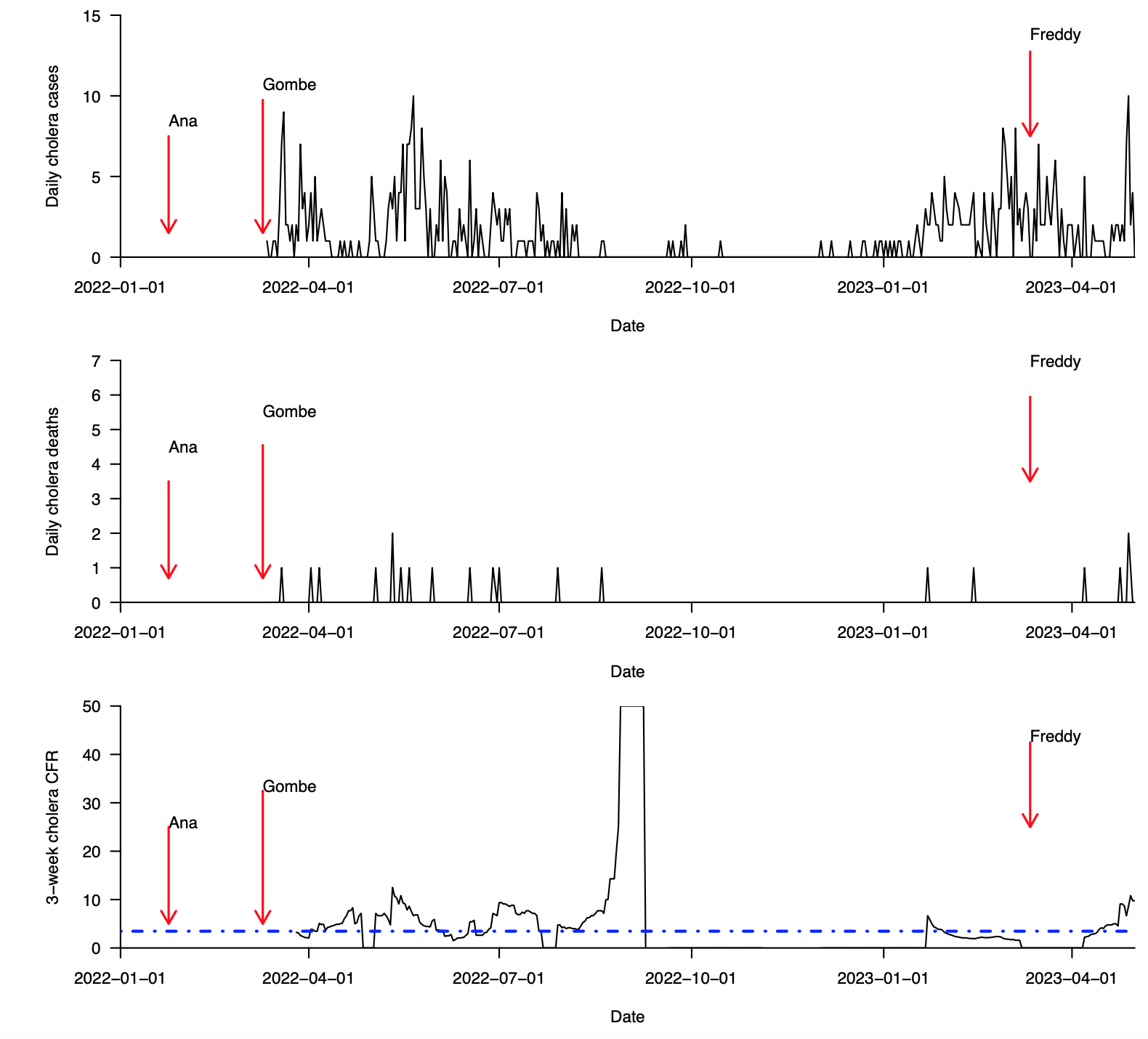

**Supplementary Fig. 2 (continued): Cases, deaths, and case fatality ratio during the 2022–2023 cholera outbreak in Balaka district in Malawi (data from January, 2022 to May, 2023). (a)** Total daily cholera cases. **(b)** Total daily cholera deaths. **(c)** Overall cholera case fatality ratio (CFR) based on a 21-day sliding window. The 21-day sliding window was chosen to obtain stable estimates of the CFR, especially during weeks and months with few reported cholera cases. Data were obtained from the Public Health Institute of Malawi, Malawi Ministry of Health (MoH) data on May 20, 2023 [(https://cholera.health.gov.mw/surveillance](https://cholera.health.gov.mw/surveillance)).

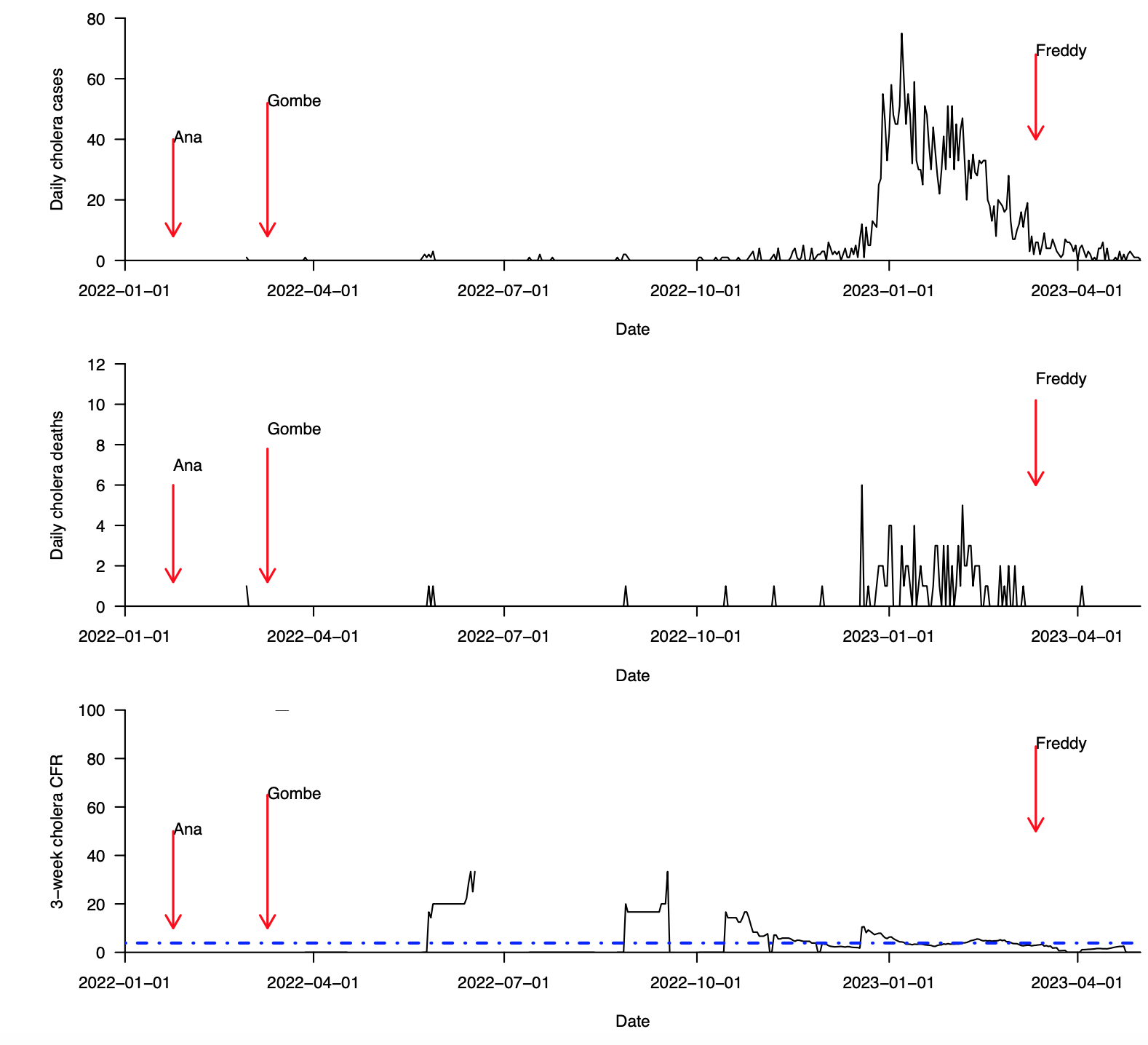

**Supplementary Fig. 2 (continued): Cases, deaths, and case fatality ratio during the 2022–2023 cholera outbreak in Mulanje district in Malawi (data from January, 2022 to May, 2023). (a)** Total daily cholera cases. **(b)** Total daily cholera deaths. **(c)** Overall cholera case fatality ratio (CFR) based on a 21-day sliding window. The 21-day sliding window was chosen to obtain stable estimates of the CFR, especially during weeks and months with few reported cholera cases. Data were obtained from the Public Health Institute of Malawi, Malawi Ministry of Health (MoH) data on May 20, 2023 [(https://cholera.health.gov.mw/surveillance](https://cholera.health.gov.mw/surveillance)).

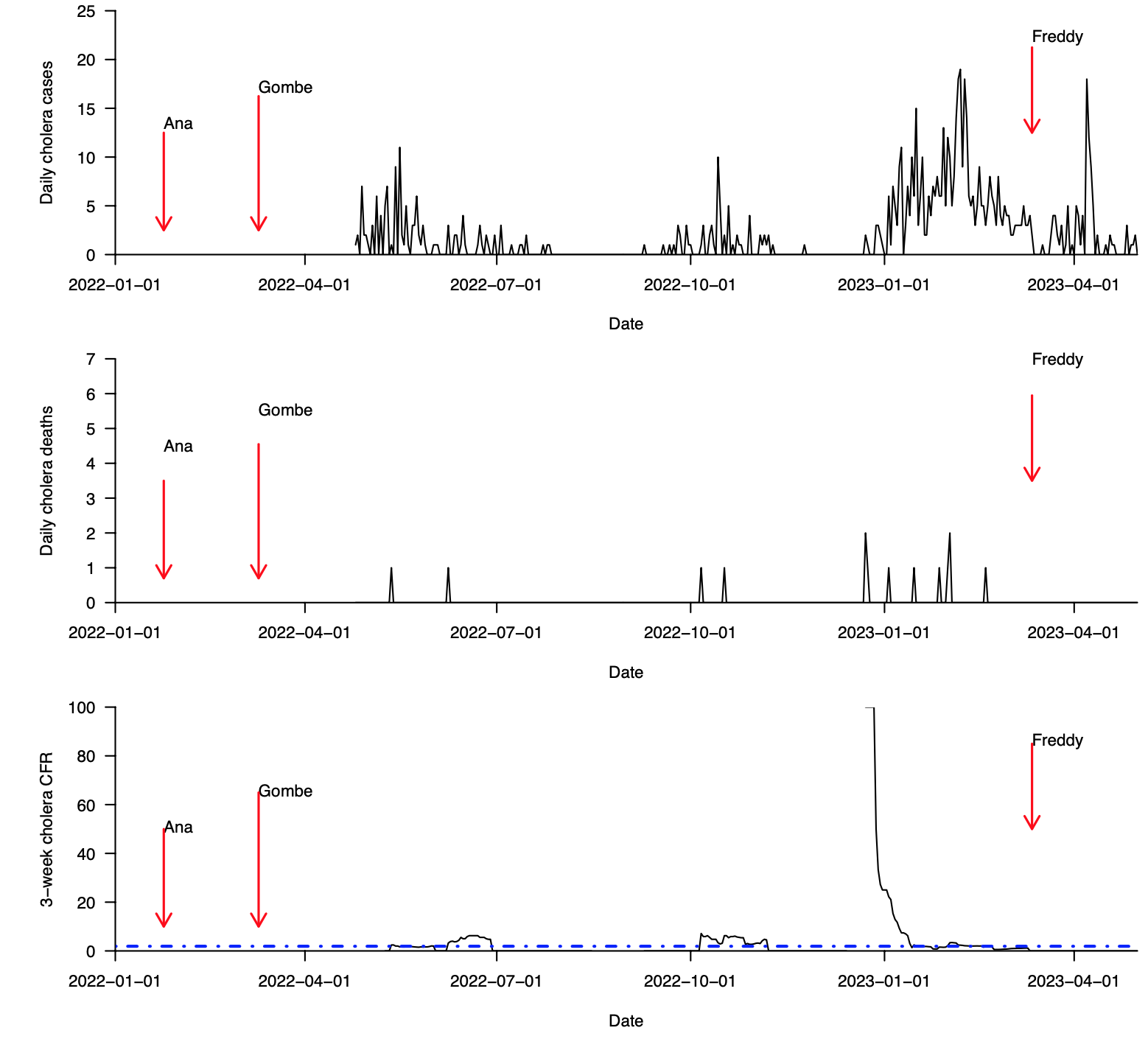

**Supplementary Fig. 2 (continued): Cases, deaths, and case fatality ratio during the 2022–2023 cholera outbreak in Neno district in Malawi (data from January, 2022 to May, 2023). (a)** Total daily cholera cases. **(b)** Total daily cholera deaths. **(c)** Overall cholera case fatality ratio (CFR) based on a 21-day sliding window. The 21-day sliding window was chosen to obtain stable estimates of the CFR, especially during weeks and months with few reported cholera cases. Data were obtained from the Public Health Institute of Malawi, Malawi Ministry of Health (MoH) data on May 20, 2023 [(https://cholera.health.gov.mw/surveillance](https://cholera.health.gov.mw/surveillance)).

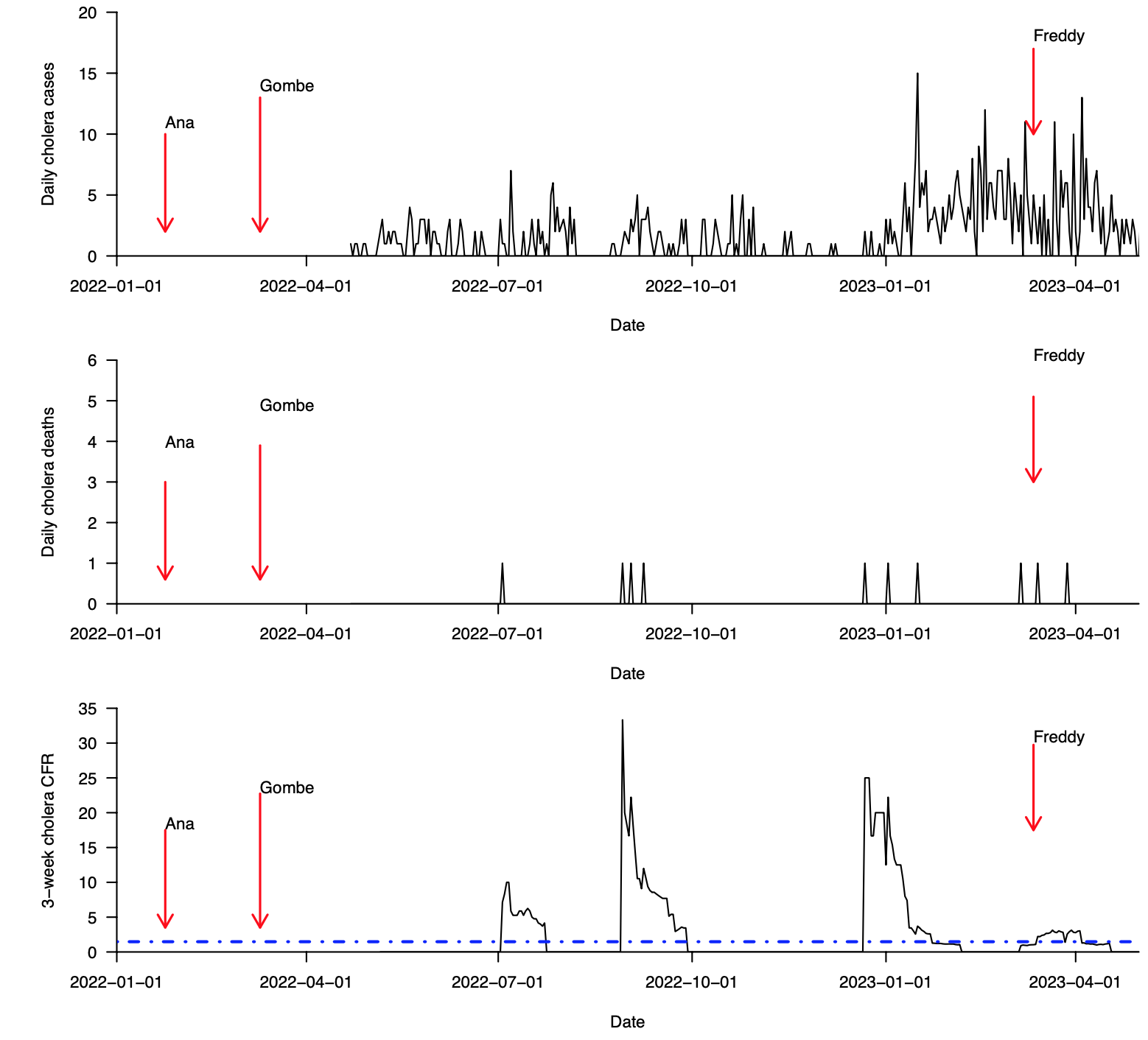

**Supplementary Fig. 2 (continued): Cases, deaths, and case fatality ratio during the 2022–2023 cholera outbreak in Chikwawa district in Malawi (data from January, 2022 to May, 2023). (a)** Total daily cholera cases. **(b)** Total daily cholera deaths. **(c)** Overall cholera case fatality ratio (CFR) based on a 21-day sliding window. The 21-day sliding window was chosen to obtain stable estimates of the CFR, especially during weeks and months with few reported cholera cases. Data were obtained from the Public Health Institute of Malawi, Malawi Ministry of Health (MoH) data on May 20, 2023 [(https://cholera.health.gov.mw/surveillance](https://cholera.health.gov.mw/surveillance)).

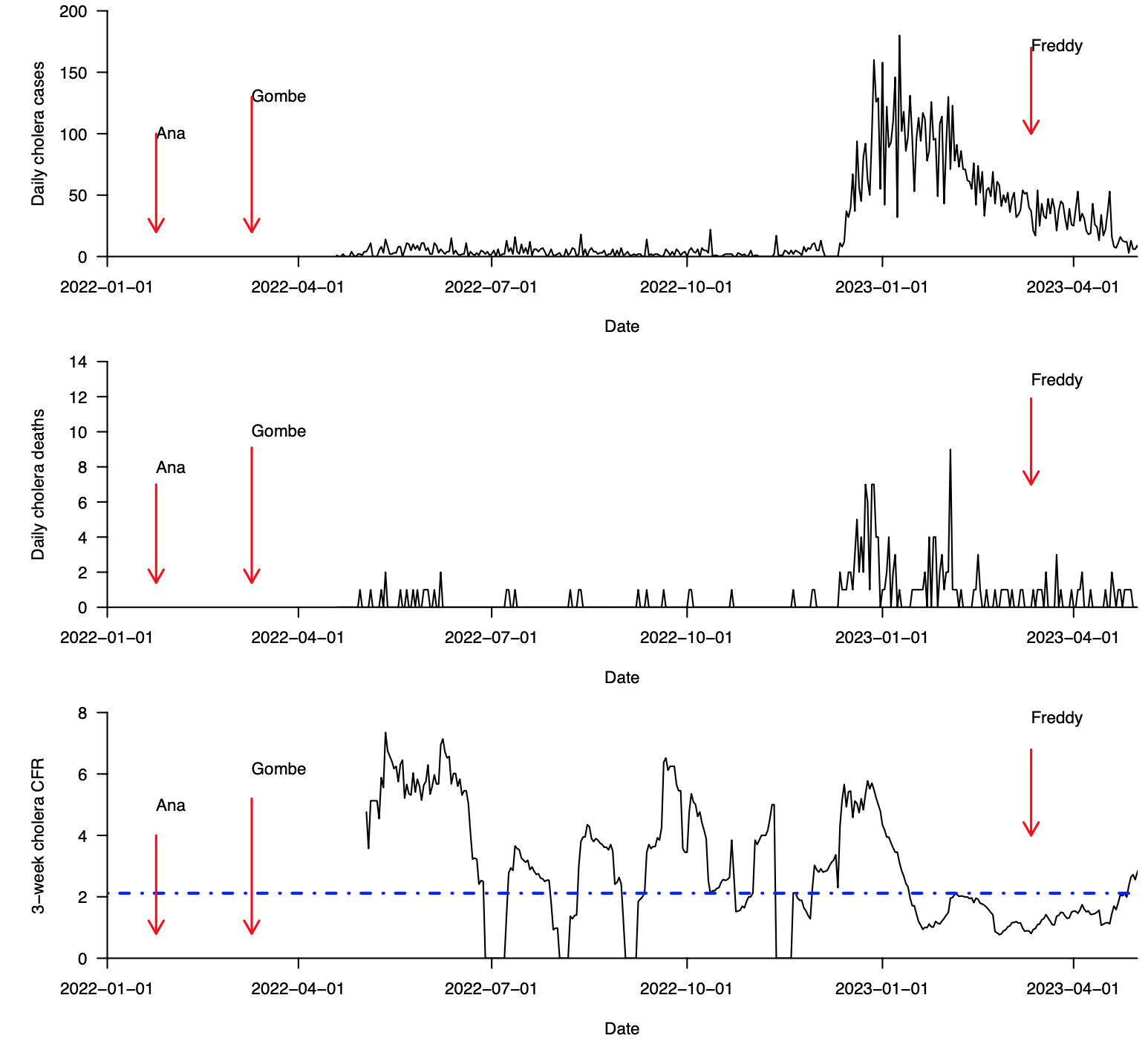

**Supplementary Fig. 2 (continued): Cases, deaths, and case fatality ratio during the 2022–2023 cholera outbreak in Blantyre district in Malawi (data from January, 2022 to May, 2023). (a)** Total daily cholera cases. **(b)** Total daily cholera deaths. **(c)** Overall cholera case fatality ratio (CFR) based on a 21-day sliding window. The 21-day sliding window was chosen to obtain stable estimates of the CFR, especially during weeks and months with few reported cholera cases. Data were obtained from the Public Health Institute of Malawi, Malawi Ministry of Health (MoH) data on May 20, 2023 [(https://cholera.health.gov.mw/surveillance](https://cholera.health.gov.mw/surveillance)).

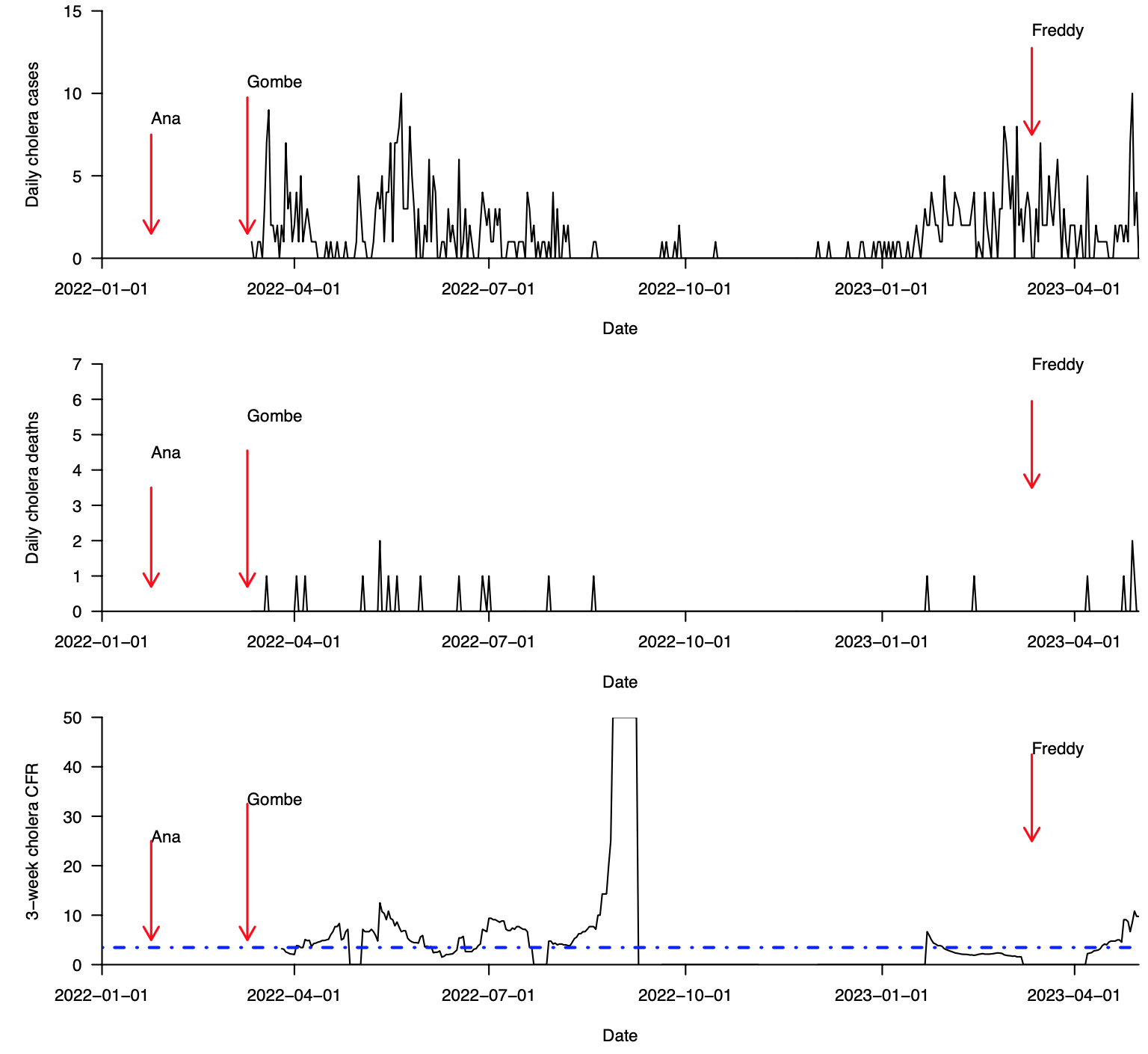

**Supplementary Fig. 2 (continued): Cases, deaths, and case fatality ratio during the 2022–2023 cholera outbreak in Nsanje district in Malawi (data from January, 2022 to May, 2023). (a)** Total daily cholera cases. **(b)** Total daily cholera deaths. **(c)** Overall cholera case fatality ratio (CFR) based on a 21-day sliding window. The 21-day sliding window was chosen to obtain stable estimates of the CFR, especially during weeks and months with few reported cholera cases. Data were obtained from the Public Health Institute of Malawi, Malawi Ministry of Health (MoH) data on May 20, 2023 [(https://cholera.health.gov.mw/surveillance](https://cholera.health.gov.mw/surveillance)).

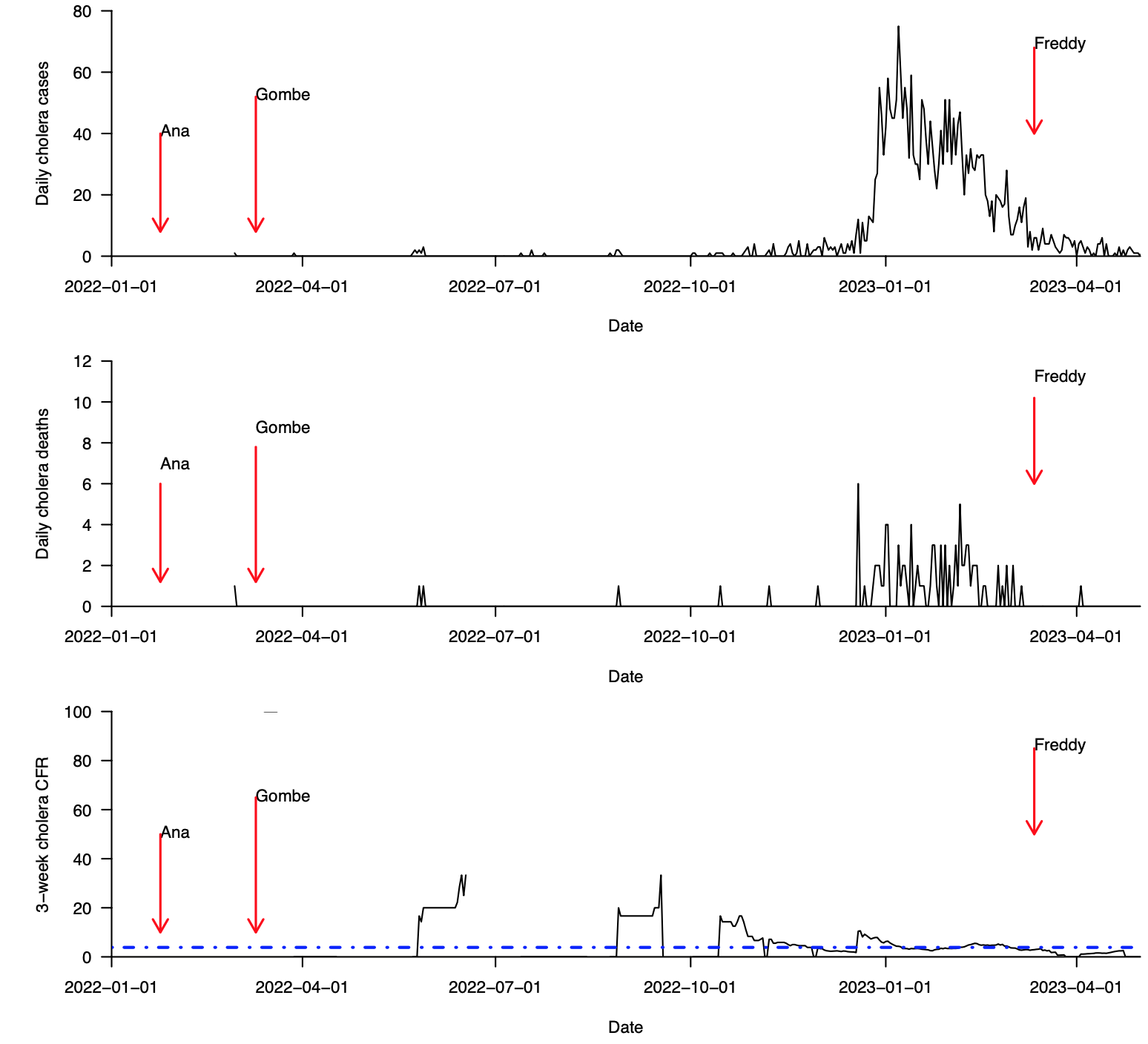

**Supplementary Fig. 2 (continued): Cases, deaths, and case fatality ratio during the 2022–2023 cholera outbreak in Machinga district in Malawi (data from January, 2022 to May, 2023). (a)** Total daily cholera cases. **(b)** Total daily cholera deaths. **(c)** Overall cholera case fatality ratio (CFR) based on a 21-day sliding window. The 21-day sliding window was chosen to obtain stable estimates of the CFR, especially during weeks and months with few reported cholera cases. Data were obtained from the Public Health Institute of Malawi, Malawi Ministry of Health (MoH) data on May 20, 2023 [(https://cholera.health.gov.mw/surveillance](https://cholera.health.gov.mw/surveillance)).
